# Modelling COVID-19 contagion: Risk assessment and targeted mitigation policies

**DOI:** 10.1101/2020.08.26.20182477

**Authors:** Rama Cont, Artur Kotlicki, Renyuan Xu

## Abstract

We use a spatial epidemic model with demographic and geographic heterogeneity to study the regional dynamics of COVID-19 across 133 regions in England.

Our model emphasises the role of variability of regional outcomes and heterogeneity across age groups and geographic locations, and provides a framework for assessing the impact of policies targeted towards sub-populations or regions. We define a concept of efficiency for comparative analysis of epidemic control policies and show targeted mitigation policies based on local monitoring to be more efficient than country-level or non-targeted measures. In particular, our results emphasise the importance of shielding vulnerable sub-populations and show that targeted policies based on local monitoring can considerably lower fatality forecasts and, in many cases, prevent the emergence of second waves which may occur under centralised policies.

## 1 Overview

The novel coronavirus pandemic of 2019-2020 has led to disruption on a global scale, leading to more than 800,000 deaths worldwide at the time of writing, and prompted the implementation of government policies involving a variety of ’non-pharmaceutical interventions’[20] including school closures, workplace restrictions, restrictions on social gatherings, social distancing and, in some cases, general lockdowns for extended periods. This has led to a range of different public health policies across the world, and the efficiency of specific policy choices has been subject to much debate.

While the nature of these restrictions has been justified by the severe threat to public health posed by the virus, their design and implementation necessarily involves a trade-off, often implicit in the decision-making process, between health outcomes and the socioeconomic impact of such social restrictions.

An important feature of the COVID-19 pandemic has been the heterogeneity of epidemic dynamics and the resulting mortality across different regions, age classes and population categories. The importance of these heterogeneities suggests that homogeneous models –often invoked in discussions on reproduction number and herd immunity– may provide misleading insights, and points to the need for more granular modeling to take into account geographic, demographic and social factors which may influence epidemic dynamics.

We propose a flexible modelling framework which can serve as a decision aid to policy makers and public health experts by quantifying this trade-off between health outcomes and social cost. Using a structured population model for epidemic dynamics which accounts for geographic and demographic heterogeneity, we formulate this trade-off as a control problem for a partially observed distributed system and provide a quantitative framework for comparative analysis of various mitigation policies. We illustrate the usefulness of the framework by applying it to the study of COVID-19 dynamics across regions in England and showing how it may be used to reconstruct the latent progression of the epidemic and perform a comparative analysis of various mitigation policies through scenario projections.

Several recent studies have used homogeneous compartmental models [3, 36, 17, 29, 42, 48, 45, 49] or age-stratified versions of such models [1, 15, 16, 34, 44, 52] to analyse the dynamics and impact of the COVID-19 epidemic in various countries. Our framework, while compatible with such homogeneous models at aggregate level, accounts for demographic and spatial heterogeneity in a more detailed manner, leading to regional outcomes which may substantially deviate from such models, as discussed in Section 5.2. Similar, though somewhat less detailed, heterogeneous models have been recently used to study COVID-19 outbreaks by Birge et al. [10] for New York City and Roques et al. [47] for France.

We first present below an overview of the main features of our approach and the key findings, before going into more detail on the methodology and results.

### 1.1 Methodology

We formulate a stochastic compartmental (SEIAR) epidemic model with spatial and demographic heterogeneity (age stratification) for modeling the dynamics of the COVID-19 epidemic and apply this model to the study of COVID-19 dynamics across regions in England.

The model takes into account

- Epidemiological features estimated by previous studies on COVID-19;
- The lack of direct observability of the total number of infectious cases and the presence of a non-negligible fraction of asymptomatic cases;
- The demographic structure of UK regions (age distribution, density);
- Social contact rates across age groups derived from survey data;
- Data on inter-regional mobility; and
- The presence of other random factors, not determined by the above.

We first demonstrate that this model is capable of accurately reproducing the early regional dynamics of the disease, both pre-lockdown and a month into lockdown, using a detailed calibration procedure that accounts for demographic heterogeneity across regions, low testing rates, and existence of asymptomatic carriers. The calibration reveals interesting regional patterns in social contact rates before and during lockdown.

Underlying any public health policy is a trade-off between a health outcome –which may relate to mortality or hospitalizations– and the socio-economic impact of measures taken to mitigate the magnitude of the impact on public health. We present an explicit formulation of this trade-off and use it to perform a comparative analysis of various ‘social distancing’ policies, based on two criteria:

- the benefit, in terms of reduction in projected mortality; and
- the cost, in terms of restrictions on social contacts.

The goal of our analysis is to make explicit the policy outcomes for decision-makers, without resorting to (questionable) concepts such as the ’economic value of human life’ used in some actuarial and economic models [1, 42, 49].

In our comparative analysis, we consider a broad range of policies and pay particular attention to population-wide versus *targeted* mitigation policies, feedback control based on the number of observed cases and the benefits of broader testing. We introduce a concept of *efficient* policy, and show how this concept allows to identify decision parameters which lead to the most efficient outcomes for each type of mitigation policy. The granular nature of our model, together with validation based on epidemiological data, provide a more detailed picture of the relative merits of various public health policies.

### 1.2 Summary of findings

Our first set of results concerns the reconstruction of the progression of the pandemic in England, in particular its latent spread through asymptomatic carriers.

- Using a baseline epidemic model consistent with epidemiological data and observations on fatalities and cases reported in England up to June 2020, we estimate the current number of infected individuals in England on August 1, 2020 to be more than 1 million, including 233,000 (or 22%) symptomatic and 858,000 (or 78%) asymptomatic carriers. We also estimate more than 17.8 million persons in England (31.7% of the population) to been exposed to COVID-19 and recovered by August 1, 2020. These estimates are much higher than numbers discussed in media reports, often based on the number of *reported* cases, suggesting that England is closer to ‘herd immunity’ than previously thought.
- Based on a comparison of fatality counts and reported cases, we infer that less than 5% of cases in England had been detected prior to June 2020. This low reporting probability has important consequences: in particular, it implies that one may observe a streak of many consecutive days without any new reported cases whilst the epidemic is in fact silently progressing.
- We observe significant differences in epidemic dynamics across regions in England, with higher rates of contagion in northern regions compared to southern regions, both before and during the lockdown period.
- We estimate that, in absence of social distancing and confinement measures, the number of fatalities in England may have exceeded 216,000 by August 1, 2020, indicating that the lockdown has saved more than 174,000 lives.
- Comparison with a homogeneous model with compartments defined at national level reveals large differences in regional forecasts of case numbers and fatalities, pointing to the importance of demographic and geographic heterogeneity for modeling the impact of COVID-19.

Once the model has been calibrated to replicate the regional progression of COVID-19 in England for the period March 1–May 31, 2020, we use it for scenario projections under various mitigation policies. Comparative analysis of mitigation policies reveals that measures targeted towards subpopulations –vulnerable age groups or regions with outbreaks– are more efficient than measures applied at the population as a whole. More specifically:

- Shielding of elderly populations is by far the most effective measure for reducing the number of fatalities.
- By contrast, school closures and workplace restrictions are seen to be less effective than social distancing measures outside of school and work environments.
- Adaptive policies (‘feedback control’) which trigger measures when the number of daily observed cases exceed a threshold, are shown to be more effective than preplanned policies, leading to a substantial improvement in health outcomes. As such policies are based on monitoring of new cases, broader testing significantly improves their outcome.
- Decentralised policies based on regional monitoring of daily reported cases are found to more efficient than centralised policies based on national indicators, resulting on average in an overall reduction of 20,000 in fatalities and, in many cases, the prevention of a ‘second wave’ which may occur under centralised policies.
- Comparative analysis of policies (Table 8) shows a wide range of health outcomes, ranging from 53,000 to 165,000 fatalities. The most effective policy in terms of reducing fatalities involves triggering of regional confinement measures decentralised based on monitoring of new cases, together with shielding of elderly populations.

### 1.3 Outline

The modeling framework is described in Section 2. Data sources and parameter estimations are detailed in Section 3. Section 4 highlights the implications of partial observability of state variables and the associated model uncertainty. Section 5 discusses the counterfactual scenario of no intervention, which serves as a benchmark to evaluate the impact of social distancing policies.

The outcomes of various epidemic control policies are then discussed in Sections 6 and 7. Pre-planned policies are discussed in Sections 6.1 and 6.2, while Section 7 discusses *adaptive* (‘feedback’) control policies, in which measures are triggered when the daily number of new reported cases exceeds a threshold.

## 2 Modeling framework

We now describe the framework used to model the dynamics of the epidemic. To take into account the role of geographic and demographic heterogeneity, we use a stochastic compartmental (SEIAR) model with age stratification, mobility across sites, social contact across age stratification, and the impact of asymptomatic infected individuals. For general concepts on deterministic and stochastic compartmental models we refer to Anderson and May [5], Andersson and Britton [6], Brauer and Castillo-Chavez [11], Britton et al. [12], Grassly and Fraser [23], Lloyd and Jansen [35].

### 2.1 State variables

We consider a regional meta-population model with *K* regions labeled *r* = 1,…, *K*. Each region *r* has a population *N*(*r*) which is further subdivided into 4 age classes, labeled *a* ∈ {1, 2, 3, 4} representing respectively children (below 20 years), adults (19–60) and two senior groups (60–70 years and above 70 years). We denote *N*(*r*, *a*) the population in region *r* in age category *a*, with *N*(*r*, 1) + *N*(*r*, 2) + *N*(*r*, 3) + *N*(*r*, 4) = *N*(*r*).

Individuals in each region and age group are categorized into six compartments:

- Susceptible (*S*) individuals who have not yet been exposed to the virus;
- Exposed (*E*) individuals who have *contracted* the virus but are not yet infectious. Exposed individuals may then become *infectious* after a certain incubation period;
- Infectious (*I*) individuals who manifest symptoms;
- *Asymptomatic* (*A*) infectious individuals;
- Recovered (*R*) individuals. In line with current experimental and clinical observations on COVID-19, we shall assume that individuals who have recovered have temporary immunity, at least for the horizon of the scenarios considered, and cannot be reinfected [8]; and
- Deceased (*D*) individuals.

The progression of the disease in the population is monitored by keeping track of the respective number

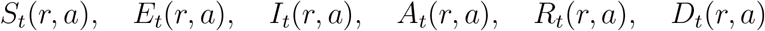

of individuals in each compartment. As the model focuses on the dynamics of the epidemic over a short period (1000 days), we neglect demographic changes over this period and assume that the population size *N*(*r, a*) in each location and age group is approximately constant, that is

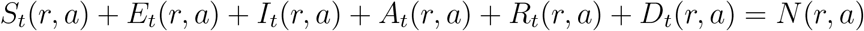

is constant.

### 2.2 A metapopulation SEIAR model

When each subpopulation (*r, a*) is large and homogeneous, the dynamics of state variables may be described through the following system of equations, represented in Figure 1:

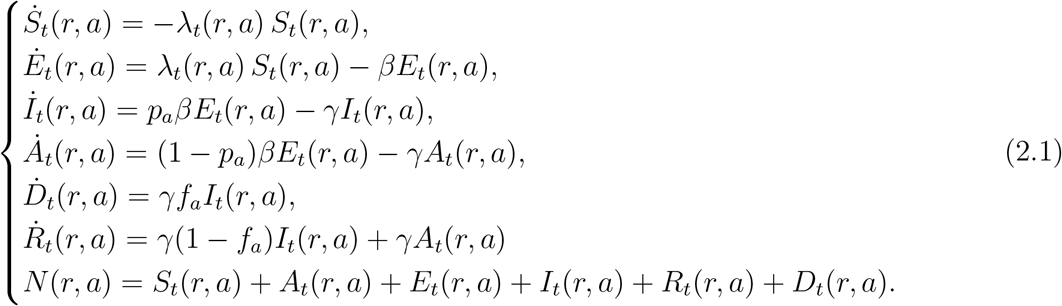

where

- 0 *< α <* 1 is the infection rate per contact, that is the probability of infection conditional on contact;
- *β* is the incubation rate, and 1*/β* is the average incubation period;
- γ is the rate at which infectious individuals recover;
- 0 < *p_a_ <* 1 is the probability for an infected individual in age group *a* to develop symptoms;
- *f_a_* is the infection fatality rate for age group *a*, representing the probability that an infected individual in age group *a* dies from the disease; and
- The *force of infection λ_t_*(*r, a*), which measures the rate of exposure at location *r* for age group *a*, is given by

**Figure 1:**
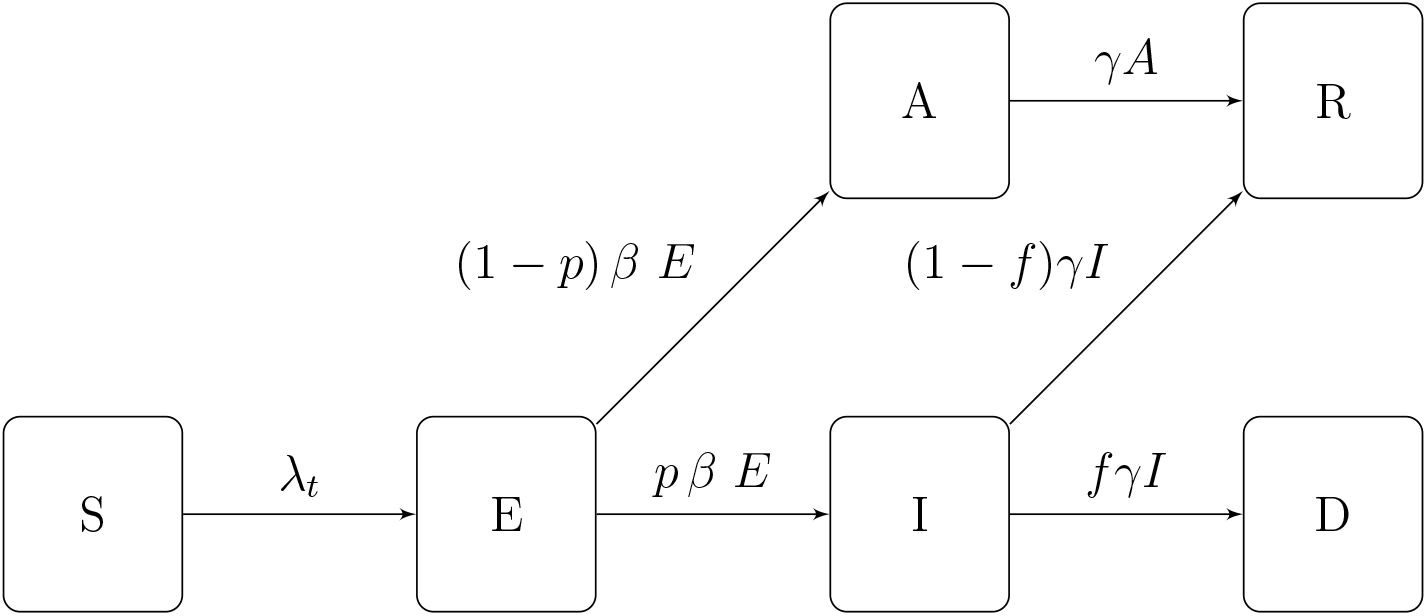
Epidemic dynamics.

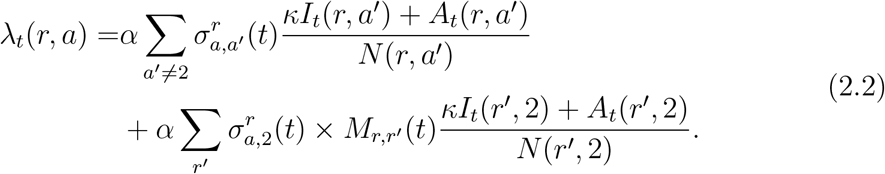

The force of infection in each subpopulation depends on the rate of contact with (infected) individuals in other subpopulations, leading to interactions across subpopulations which differentiate this model from a homogeneous model. These interactions occur through:

- *Contacts across age groups:* the term 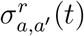 represents the average number of persons from age class *a*′ encountered per day by a person from age class *a* in region *r* on a day *t*. For infectious individuals with symptoms, we assume a lower contact rate *κσ < σ* due to (partial) self-isolation; and
- *Inter-regional mobility: M_r,r′_* (*t*) represents the proportion of individuals from region *r′* among the population of adults at a location *r* on a given day *t*.

### 2.3 Stochastic dynamics

The deterministic model described in Section 2.2 ignores the variability of outcomes [25], as random factors are not taken into account in the model. To take into account the variability of outcomes and assess their respective probabilities, we model the variables (*S*(*t*), *E*(*t*), *I*(*t*), *A*(*t*)) as a continuous-time Markov point process [4, 6] defined through its transition rates conditional on the history 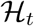 up to date *t*:

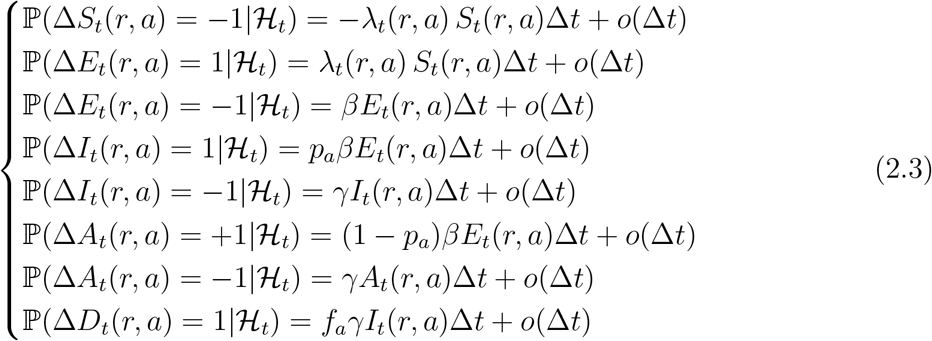

The stochastic dynamics in (2.3) are consistent with the deterministic dynamics of (2.1) for large populations, in the sense that the population fractions represented by each compartment converge to those represented by the solution of (2.1) as min*_r_ N*(*r*) increases. However, even when the overall population is large, the stochastic dynamics (2.3) can substantially deviate from the deterministic model (2.1), especially in small subpopulations and in the early phases of the epidemic when the number of infected individuals in each region may be small.

In the sequel we use the stochastic model (2.3) for the state variables and occasionally compare the outcomes with (2.1) to assess the variability of outcomes and the role of randomness (see Section 5.3).

### 2.4 Policies for epidemic control

Social distancing policies (and lockdowns) affect epidemic dynamics by influencing (lowering) the social contact rates 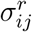 and the inter-regional mobility *M_r,r_*_′_. To discuss targeted policies which may influence differently social contact rates at different locations, we decompose the baseline social contact matrix *σ^r^* as

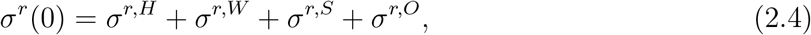

where the components correspond respectively to contacts at home (*σ^r,H^*), at work (*σ^W^*), school (*σ^S^*) and other locations (*σ^O^*). Social distancing policies are then parameterised in terms of their impact on various components of the social contact matrix:

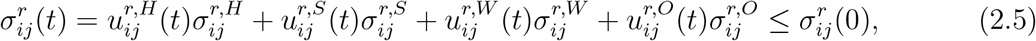

where 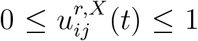 are modulating factors which measure the impact of the policy on social contacts between age groups *i* and *j* at a location *X* in region *r*. In absence of social distancing or confinement measures, we have 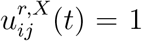; the value of 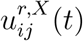 reflects the fraction of social contacts when between age groups *i* and *j* at location *X* in region *r* when the policy is applied.

This parameterisation allows us to consider policies targeted towards sub-population or specific regions. For example, school closure in region *r* during time period [*t*_1_, *t*_2_] corresponds to setting 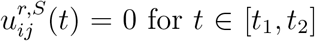 for *t* ∈ [*t*_1_, *t*_2_], while 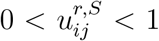 corresponds to social distancing in schools, with lower values of 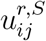 corresponding to stricter enforcement of measures.

Regarding the inter-regional mobility matrix *M*, following the interpretation discussed in Section 3.2, we modulate its value according to the fraction 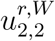 of the population who continue to commute, that is

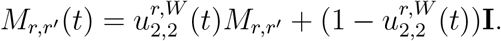

Here *M_r,r′_* is the fraction of population in region *r* whose habitual residence is in region *r′* before lockdown. The modulating factors 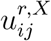 may be chosen in advance or expressed as a function of the state of the system. We distinguish:

- *Pre-planned* (also called ‘open-loop’) policies, in which target values of modulating factors 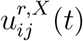 are decided in advance; and
- *Adaptive* policies (also called ‘closed loop’ or feedback control), in which actions are decided and updated as a function of observed quantities such as number of daily reported cases or number of daily fatalities.

#### Comparative analysis of mitigation policies

To perform comparative analysis across different policies, we need to evaluate policy outcomes across two dimensions: health outcome and socio-economic impact.

We quantify the health outcome of each policy by the total number of fatalities during a reference period, taken to be *t*_max_ = 1000 days after the reference date of March 1, 2020. The length of this reference period is chosen such that it takes into account an eventual ‘second wave’ of fatalities. We denote this outcome by 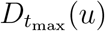, which represent the total fatalities at date *t*_max_ associated with policy *u*.

To quantify the socio-economic impact of a policy, we use as metric the reduction in social contact resulting from the policy over the horizon [0,*t*_max_], that is

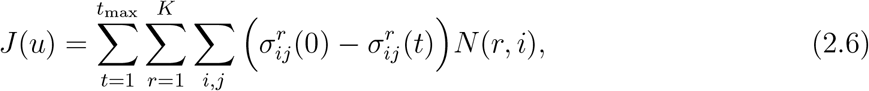

defined in terms of man × day units.

The range of policies examined below lead to different outcomes in terms of fatalities 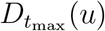 and social cost *J*(*u*). A policy *υ dominates* (or improves upon) a policy *u* if it leads to a similar or better health outcome at an equal or lower cost:

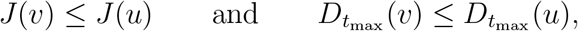

with at least one inequality being strict. A policy *u* is *efficient* among a class of policies *U* if it cannot be improved upon by any policy in this class. Given a set of policies *U*, the subset of efficient policies forms the *efficient frontier* of *U*.

Some recent economic models [1, 42, 49] formulate the trade-off in different terms, by introducing a concept of monetary *value of human life* in order to build a (monetary) welfare function combining both terms. Aside from ethical issues linked to the very concept of monetisation of human life, there is no consensus on its actual value, which is a key determinant of the trade-off in this approach. Our approach avoids specifying such a value and aims at identifying the range of efficient policies, leaving the final choice of the trade-off to policymakers.

In what follows, the goal is to determine the efficient frontier and describe the characteristics and outcomes of such efficient policies. Pre-planned policies are discussed in Sections 6.1 and 6.2, while adaptive policies are discussed in Section 7.

## 3 Data sources and parameter estimation

### 3.1 Data sources

The basic inputs of the model are panel data on number of cases and fatalities reported at the level of Upper Tier Local Authorities (UTLA) level in England^1^. This defines the granularity of the model: we partition the population of England into 133 regions as defined by the Nomenclature of Territorial Units for Statistics at level 3 (NUTS-3)^2^. Demographic data for NUTS-3 regions, is available from Eurostat^3^. For the purpose of our study we distinguish four age groups, as shown in Table 1. We perform mapping from UTLA to NUTS-3 regions to ensure consistency across data sources^4^.

**Table 1:** Age group distribution for England, 2019.

| Age group | [0,20) | [20,60) | [60,70) | [70, 100) |
| --- | --- | --- | --- | --- |
| Size | 13.21 M<br>23.6% | 29.43 M<br>52.6% | 5.86 M<br>10.5% | 7.40 M<br>13.3% |

Appendix A provides the list of UK regions used in this study. The size *N*(*r, a*) of age group *a* in region *r* is obtained using the 2018 dataset from the Office for National Statistics^5^.

### 3.2 Modeling of inter-regional mobility

For our baseline estimate of inter-regional mobility we use the data on location of usual residence and place of work, provided by the UK Office for National Statistics 2011 Census data^6^. The dataset classifies people aged 16 and over in employment during March 2011 and shows the movement between their area of residence and workplace, defined in Local Administrative Units at level 1 (LAU-1) terms. We then map this data onto NUTS-3 regions using the lookup table between LAU-1 and NUTS-3 areas provided by the Office for National Statistics^7^.

The data is then represented in the model through the inter-regional mobility matrix *M*, whose elements *M_r,j_* represent the fraction of population in region *r* whose habitual residence is in region *j*. Denote by Π(*r,j*) the population with residence registered in region *j* and workplace registered in region *r* for *r* ≠ *j*. In addition, we denote Π(*r, r*) = *N*(2, *r*) the population at location *i* in the age category 20 *–* 60 years. Then we estimate the coefficients *M_r,j_* by

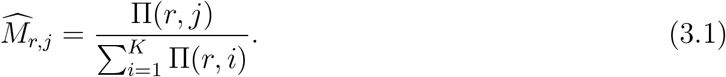

### 3.3 Epidemiological dynamics

Epidemiological parameters were either estimated from publicly available sources [2, 39] or set to values consistent with recent clinical and epidemiological studies in COVID-19 [18, 27, 53].

#### Social contact rates

Contact rates across age classes have been estimated in studies by Mossong et al. [38, 39] and Béraud et al. [9]. We follow the approach of Mossong et al. [38] to estimate the following baseline social contact rates across our four age groups (see Table 1):

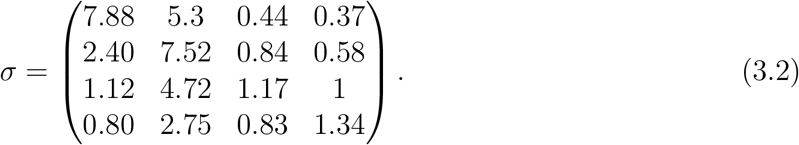

Using the PyRoss methodology of Adhikari et al. [2], we further decompose the contact matrix, as in (2.4), into four components representing contacts at home (*σ^H^*), work (*σ^W^*), school (*σ^S^*) and other locations (*σ^O^*). Estimation methods and parameter values for these matrices are discussed in Appendix B.

However, contact rates vary across different regions due to the heterogeneity in socioeconomic composition structure and specific regional characteristics, such as population density and level of urbanisation. To account for this heterogeneity, we parameterise the (pre-lockdown) contact matrix in region *r* as *σ^r^* (0) = *d_r_σ* where the regional adjustment factors {d_r_, *r* = 1.. 133} are estimated to reproduce the regional growth rate of reported cases before the lockdown period. The results are displayed in Figure 2.

**Figure 2:**
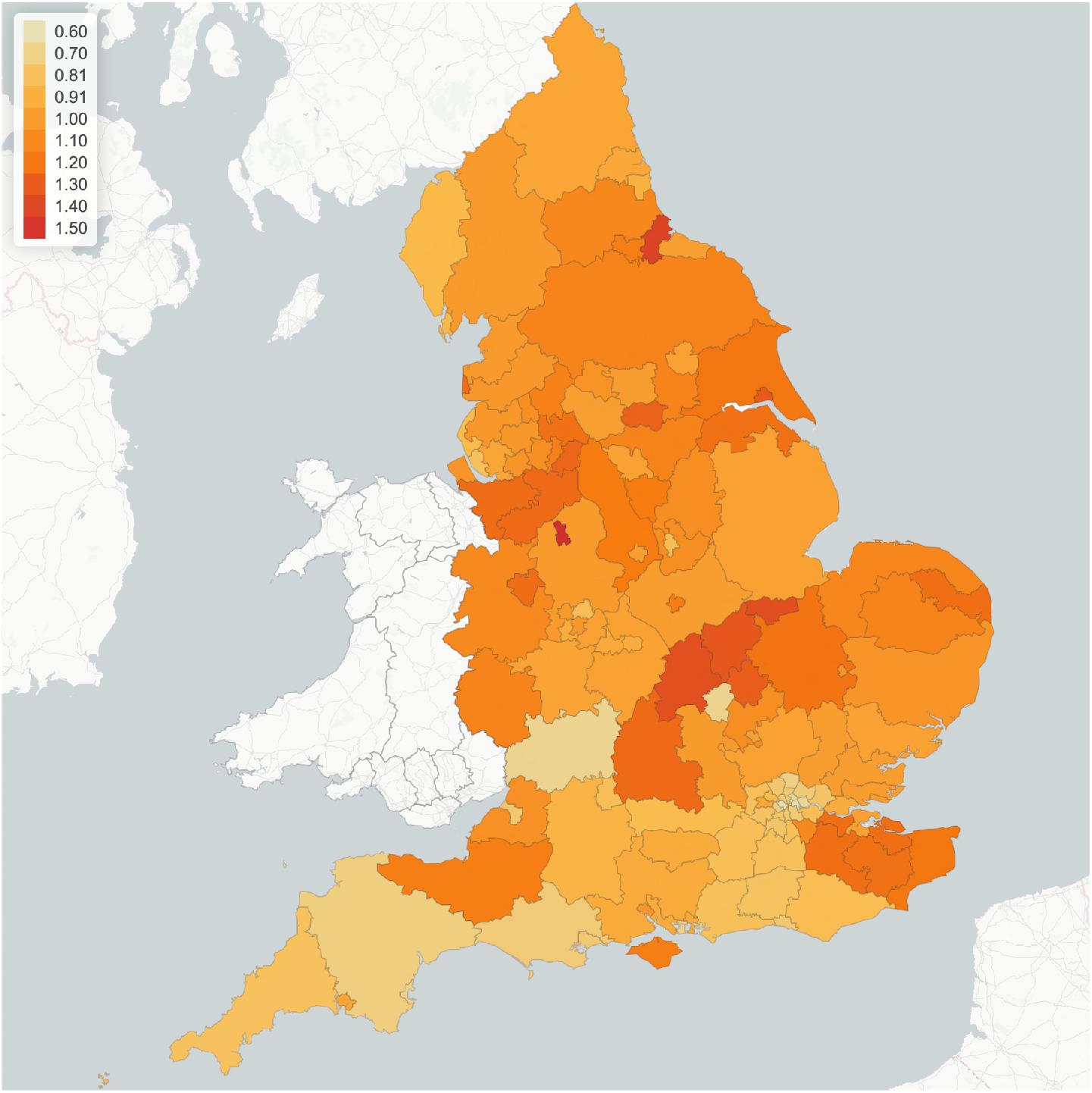
Regional multiplier *d_r_* for social contact matrix, implied by epidemic dynamics pre-lockdown (before March 23, 2020).

As seen in Figure 2, our findings imply substantial heterogeneity of social contact rates across regions. Average contact rates in some regions (in particular in southern regions) are much lower than the national average, while in other regions (in particular the North-East and North-West) they are 20% to 30% higher. As we will observe below, these differences have a considerable impact on regional epidemic dynamics.

**Incubation rate**, *β* Following the study of Ferguson et al. [20], we use the incubation rate *β* = 0.2, which corresponds to an incubation period of approximately 5 days. This is further supported by several empirical studies on diagnosed cases in China outside Hubei province. An early study of Backer et al. [7] based on 88 confirmed cases, which uses data on known travel to and from Wuhan to estimate the exposure interval, indicates a mean incubation period of 6.4 days with a 95% confidence interval (CI) of 5.6-7.7 days. Linton et al. [32], based on 158 confirmed cases, estimate a median incubation period of 5.0 days with 95% CI of 4.4-5.6 days and estimate the incubation period to have a mean of around 5 days with 95% CI of 4.2-6.0 days. Lauer et al. [30] estimates a median of incubation period to be 5.1 days with 95% CI of 4.5-5.8 days, based on 181 cases over the period of 4 January 2020 to 24 February 2020.

#### Proportion of symptomatic and asymptomatic infections

The probability *p* that an infected individual develops symptoms is an important parameter for epidemic dynamics, yet subject to a high degree of uncertainty: studies on various data sets [13, 16, 37, 21, 41] are based on small samples and yield a wide range of estimates. In particular, an early estimates from the Diamond Princess cruise ship [37] and Japanese evacuation flights from Wuhan yielded estimates as high as *p* ≃ 0.7 − 0.8 [40], while a July 2020 study by the ONS [41], based on a much larger sample, showed that p can be as low as 0.23. However, clinical studies [16] indicate that this probability may strongly depend on the age group considered.

In this study, we use a range of values for the age-dependent probability *p_a_* whose upper bound is consistent with Davies et al. [16] and whose lower bound is consistent with the estimates provided by the ONS [41]. These values are displayed in Table 2. Given the much larger sample size used in the study of the ONS [41], we use the corresponding estimates (‘low values’ in Table 2) as benchmark unless stated otherwise.

**Table 2:** Age-dependent symptomatic ratios. Source: the ONS [41].

| Age group | 0-19 | 20-59 | 60-69 | $\geq 70$ |
| --- | --- | --- | --- | --- |
| | $p_1$ | $p_2$ | $p_3$ | $p_4$ |
| **Low estimate** | 0.055 | 0.25 | 0.375 | 0.375 |
| **High estimate** | 0.11 | 0.5 | 0.75 | 0.75 |

#### Recovery rate

γ In line with Cao et al. [14], Li et al. [31] and Rocklöv et al. [46], we use a recovery rate γ = 0.1, which corresponds to an average infectious period of 10 days.

#### Infection fatality rates

We denote by *f_a_* the (infection) fatality rate for age group *a*. In practice, these parameters are difficult to estimate during outbreaks and estimates may be subject to various biases [33]. Note that the *infection* fatality rate (IFR) is different from (and generally much smaller than) the *case* fatality rate.

Fatality rates for COVID-19 have been observed to be highly variable across age groups [27, 50, 53]. Based on the infection fatality rates provided in Verity et al. [53] for different age groups and the UK population distribution, we derive the following aggregated IFR for the respective four age groups of interest: *f*_1_ = 0.01% (CI: 0.0008 − 0.037%), *f*_2_ = 0.25% (CI: 0.120 − 0.475%), *f*_3_ = 2.1% (CI: 1.11 − 3.89%), and f_4_ = 6.5% (CI: 2.96 − 10.25%). These estimates are consistent with data obtained from other countries; see for example Salje et al. [50].

### 3.4 Estimation of infection rate *α*

We use a simulation-based indirect inference method of Gourieroux et al. [22] for estimating the parameter *α*. We simulate the stochastic model (2.3) for a range of values 0.03 ≤ *α* ≤ 0.15 and compare the logarithmic growth rates of the simulated reported cases 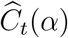 with the one estimated from reported cases *C_t_* in England.

For the simulation we use parameters specified in Table 3 and the following initial conditions for *t*_0_ = March 10, 2020: 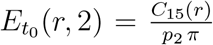, 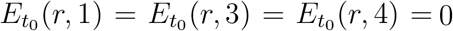, 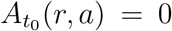, 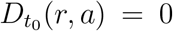, 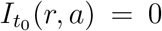. These initial conditions ensure that the simulations agree on average with regional case numbers on March 15, 2020, for all values of *α*.

**Table 3:** Clinical parameters for COVID-19 model.

| Parameter | Values | Source |
| --- | --- | --- |
| Infection rate | $\alpha = 0.055$ | Consistent with [17, 18] |
| Incubation rate | $\beta = 0.2$ | [20, 30, 32] |
| Recovery rate | $\gamma = 0.1$ | [14, 31, 46] |
| Fatality rate | $f_1 = 0.01, f_2 = 0.25, f_3 = 2.1, f_4 = 6.5$ | [53] |

This procedure yields an estimated value of 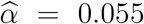 and a confidence interval [0.051, 0.062] (see Figure 3a). As shown in Figure 3b, this value of 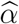, together with the model parameters in Table 3, yields a good fit of the pre-lockdown evolution of case numbers.

**Figure 3:**
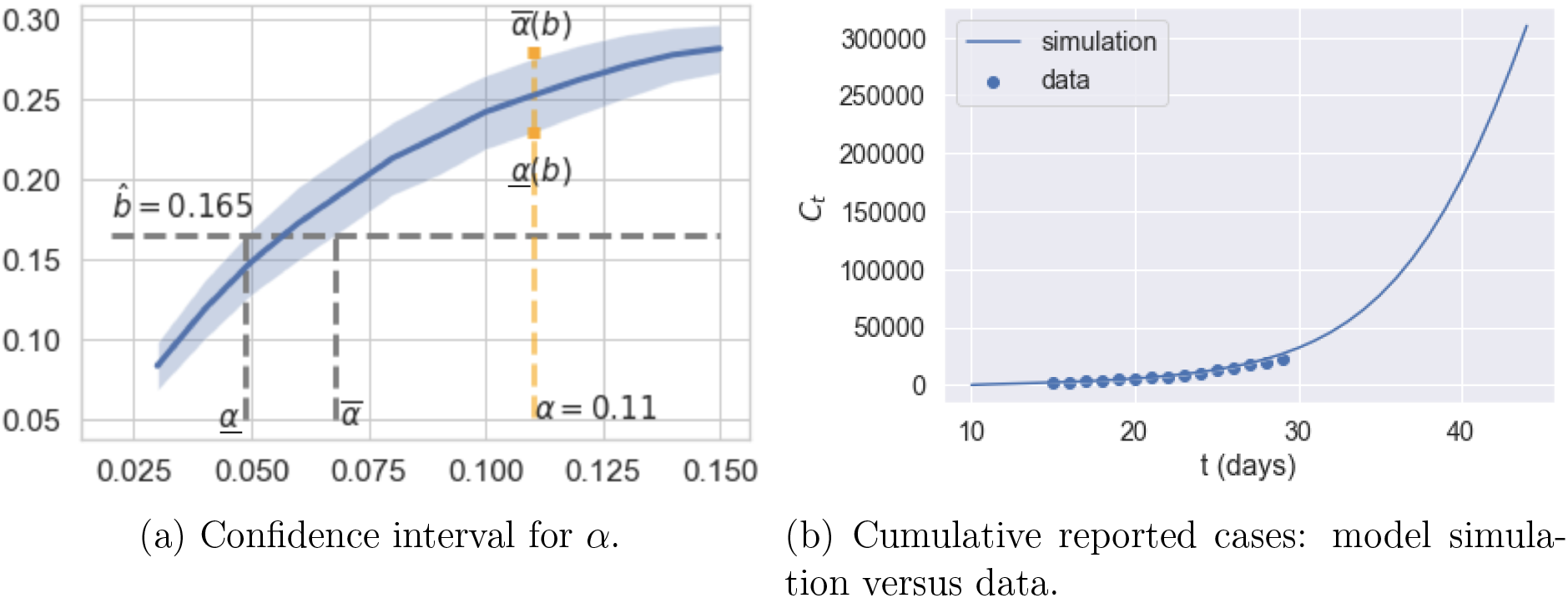
Estimation of infection rate *α* using pre-lockdown growth rate of reported cases.

These results are also consistent with estimates obtained in Donnat and Holmes [17] and Dorigatti et al. [18] using data from other countries.

### 3.5 Inter-regional mobility and social contact during confinement

Confinement measures were implemented across the United Kingdom starting March 23, 2020 via the Coronavirus Act^8^. During this ‘lockdown’ period schools and workplaces were closed and social contact was reduced, as evidenced by mobility data^9^. However mobility data also reveal regional differences in the impact of the lockdown.

We model the reduction in inter-regional mobility through an adjusted mobility matrix

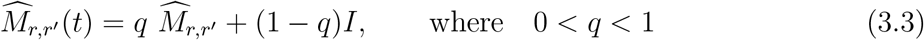

and 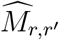 is the inter-regional mobility matrix defined in (3.1). According to the Labor Force Survey data from 2018/19 [19], 7.1 million adults across the UK are considered as ‘key workers’. We set *q* = 20% to take into account the fact that these key workers continued to access their workplace during the lockdown period. This is also consistent with the methodology in Rawson et al. [45] and empirical studies of Santana et al. [51] on mobility changes before and after lockdown in the UK. Figure 4 shows the submatrix corresponding to daily mobility across London boroughs, and illustrates the observed dramatic drop in commute patterns.

**Figure 4:**
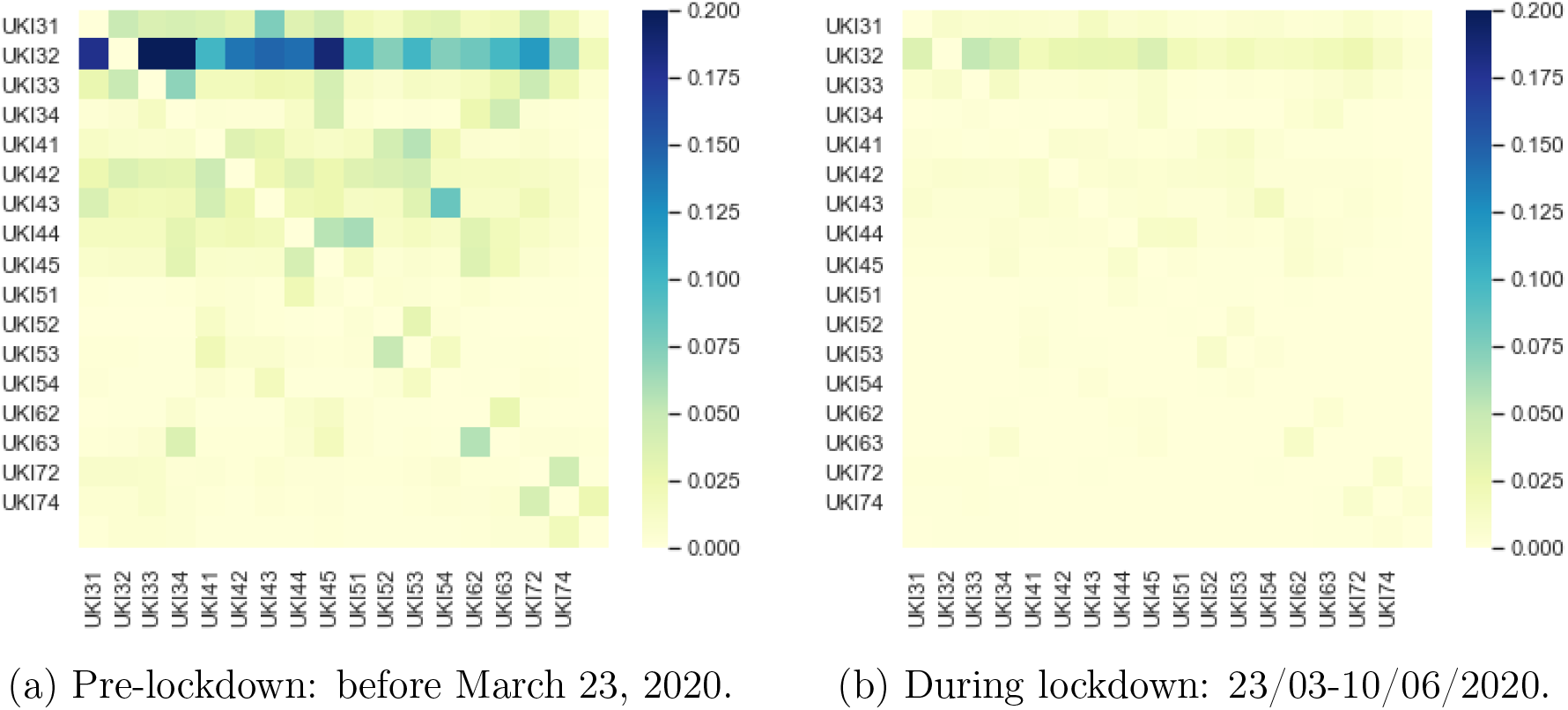
Inter-regional mobility across London boroughs.

We model the impact of confinement on the social contact matrix through a regional multiplier *l_r_*:

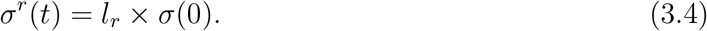

where *l_r_* ≤ *d_r_* represents the reduction in social contacts during the lockdown period; *l_r_* = *d_r_* corresponds to the pre-lockdown level of social contact. The value of *l_r_* is estimated from panel data on regional epidemic dynamics during the period from March 23 to June 1, 2020, using a least-squares logarithmic regression on the number of observed regional cases.

The average value of this reduction factor is found to be

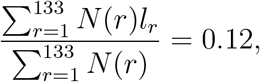

that is an average reduction of 88% in social contacts, an order magnitude corroborated by mobility data [51], showing that lockdown was very effective in reducing social contact. As shown in Figure 5 and Table 4, the estimated value of *l_r_* shows some variation across regions, ranging from 7% to 18%, implying varying levels of compliance with confinement measures across regions in England.

**Figure 5:**
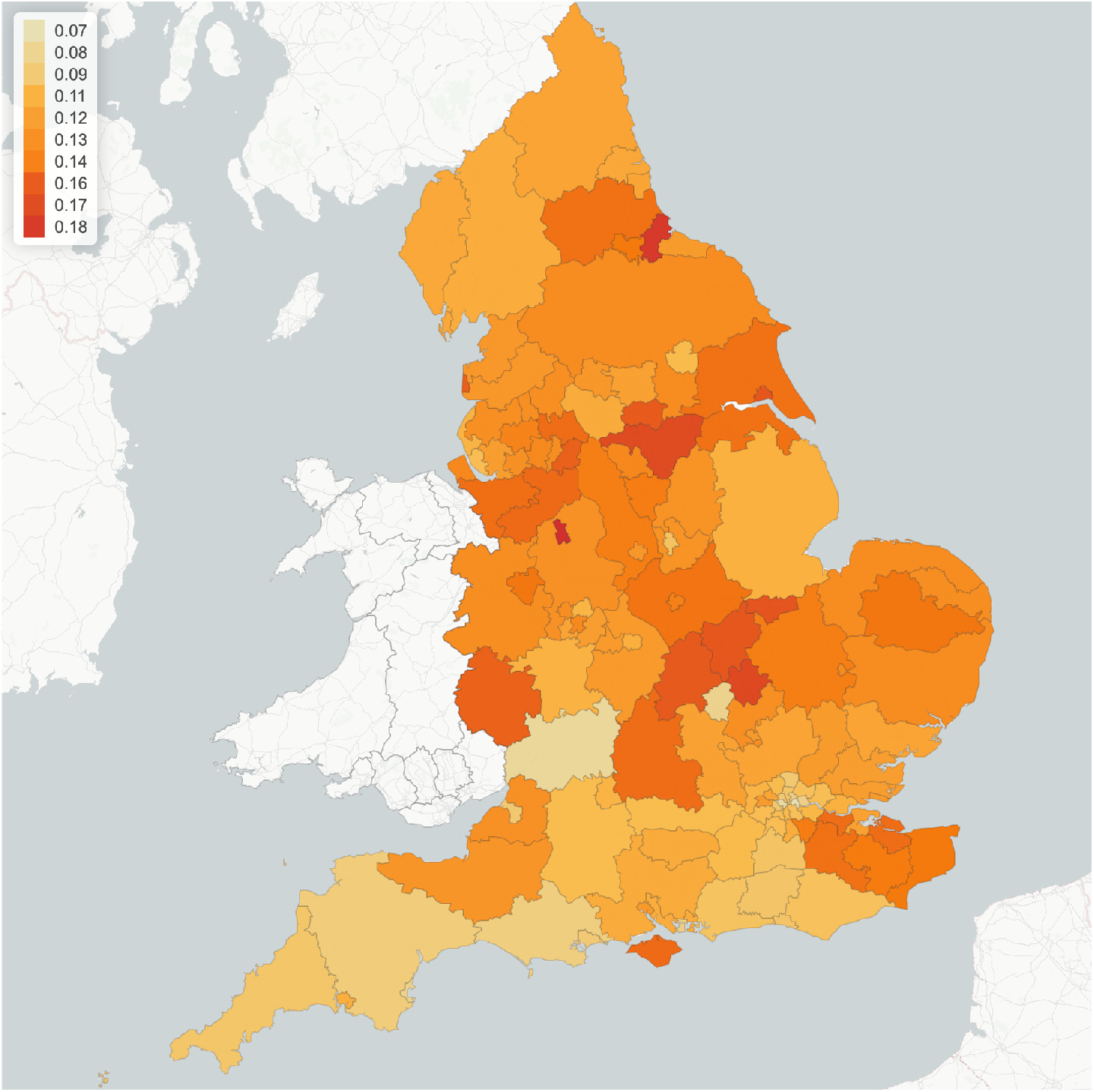
Reduction in social contact during lockdown across different regions. Values correspond to factor *l_r_* in Eq. (3.4).

**Table 4:** Estimated values for regional adjustments *d_r_* and *l_r_* in NUTS1 regions.

| NUTS1 Region | Pre-lockdown ($d_r$) | Lockdown ($l_r$) |
| --- | --- | --- |
| South West (UKK) | 0.742 | 0.099 |
| East Midlands (UKF) | 0.972 | 0.134 |
| London (UKI) | 1.166 | 0.100 |
| West Midlands (UKG) | 1.044 | 0.126 |
| Yorkshire and Humber (UKE) | 1.106 | 0.137 |
| South East (UKJ) | 0.923 | 0.115 |
| North East (UKC) | 1.284 | 0.131 |
| North West (UKD) | 1.154 | 0.133 |
| East of England (UKH) | 1.018 | 0.128 |

**Table 5:** Outcomes for policies represented in Figure 26.

| Policy | **Blue dotted:**<br>$m = 0.5, T = 335$ | **Orange dash:**<br>$m = 0.5, T = 105$ | **Green solid**<br>$m = 0.4, T = 105$ |
| --- | --- | --- | --- |
| Social cost ($\times 10^{11}$) | 3.6 | 3.0 | 3.5 |
| Projected fatalities | 164,300 | 165,500 | 143,400 |

### 3.6 Goodness-of-fit

Having estimated the model parameters using data on reported cases between March 10 and May 20, 2020 we assess the goodness-of-fit and out-of-sample performance using reported cases and fatalities between May 21 and June 22, 2020. The results, shown in Figures 6 and 7, show that the model is able to reproduce well both the in-sample and out-of-sample evolution of number of cases and fatalities, at national level as well as regional level (see Figure 8).

**Figure 6:**
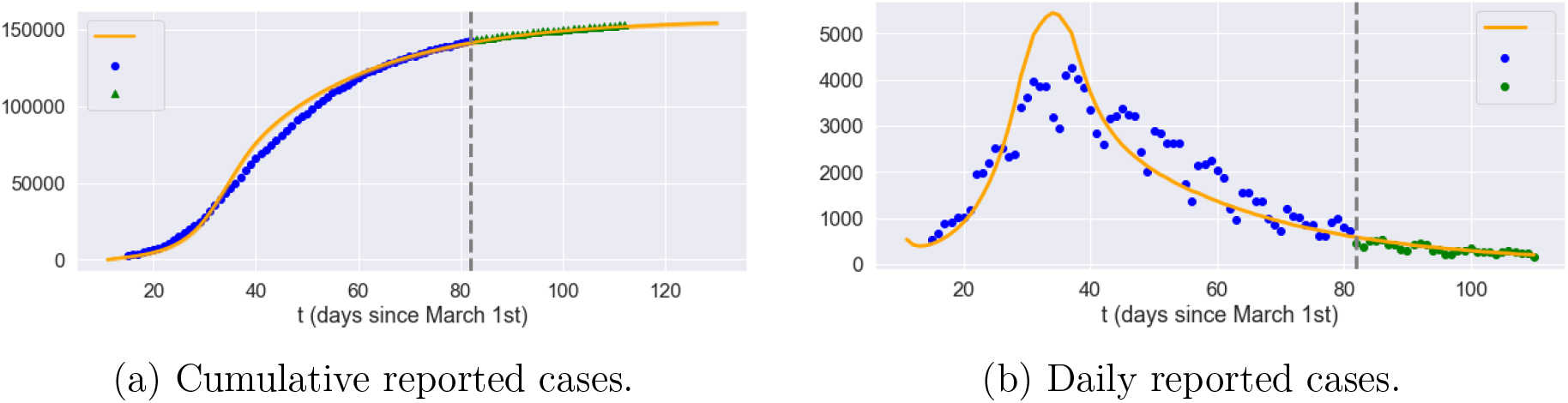
Reported cases in England. Grey dashed line: separation between estimation sample and test data; orange line: model simulation; blue dot: in-sample data; green triangle: out-of-sample data.

**Figure 7:**
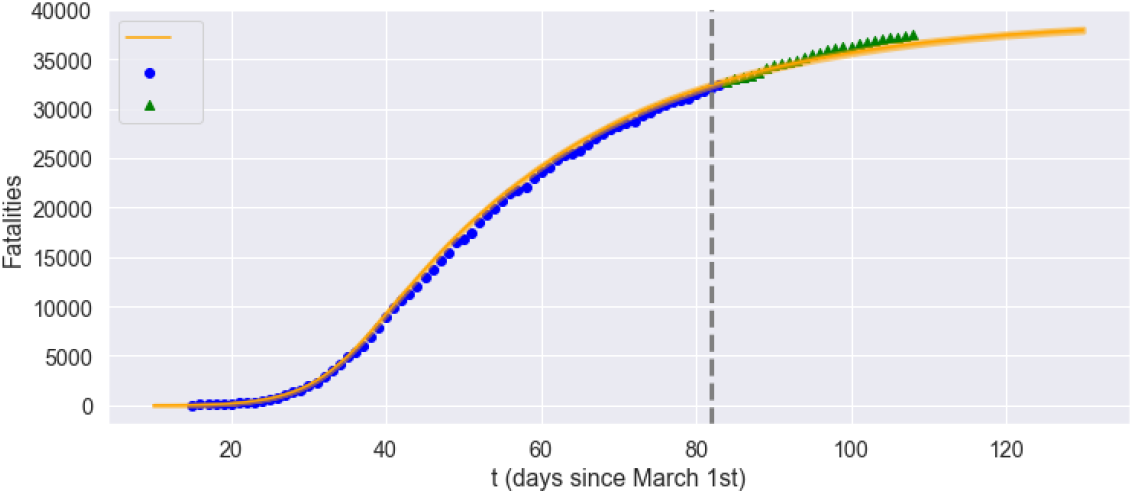
Fatalities in England: comparison of model with data. Grey dashed line: separation between estimation sample and test data; orange line: model simulation; blue dot: in-sample data; green triangle: out-of-sample data.

**Figure 8:**
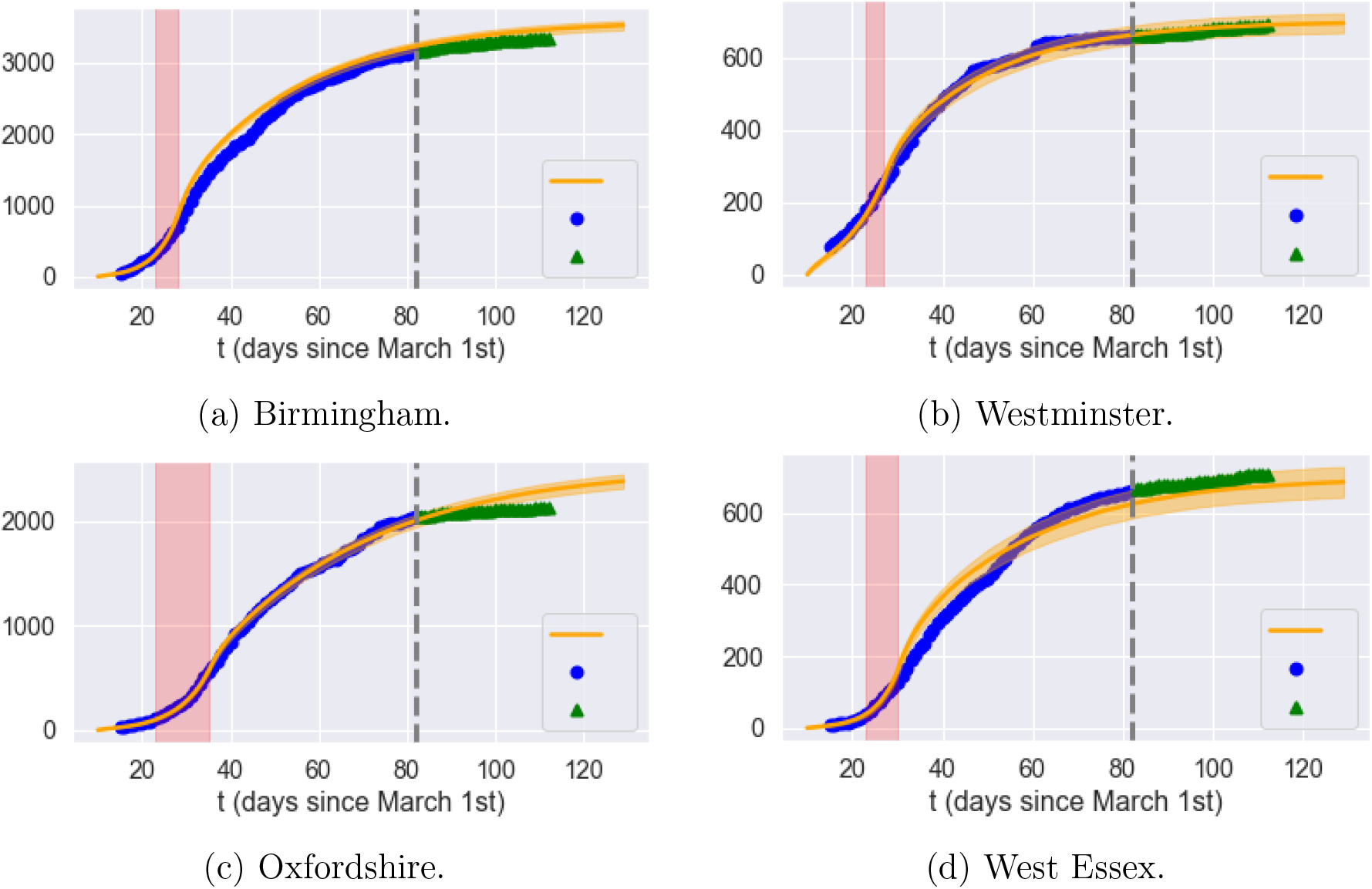
Cumulative reported cases in selected regions. Grey dashed line: separation between estimation sample and test data; orange line: average of 50 simulated scenarios; blue dot: in-sample data; green triangle: out-of-sample data.

## 4 Observable quantities and uncertainty

When applying such models to epidemic data, a key point is to realize that the state variables *S, E, I, A, R* are not directly observed (and certainly not in real time) but need to be inferred from other observable quantities.

### 4.1 Observable quantities

The two main observables in COVID-19 data are

- the cumulative number of *reported* cases, and
- the cumulative number of COVID-19 fatalities *D_t_*;

broken down by region and age group.

Of the two, fatalities are generally considered more reliable, as deaths are nearly always reported, while identification of cases requires testing or self-reporting. We thus identify the observed number of fatalities with the state variable *D_t_*.

In absence of widespread testing, as has been the case with COVID-19, only a *fraction π* of cases are reported. This fraction may change with time due to testing campaigns^10^.

We therefore cannot assume the number of infectious cases to be directly observed: rather, we estimate it from the death count *D_t_* (see also Jombart et al. [26]).

Let *C_t_* be the cumulative number of (symptomatic) infectious cases. Assuming that

- the daily number *r*(*t*) of reported cases is a fraction *π*(*t*) of new cases:

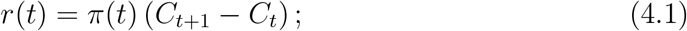
- deaths occur on average *T* days after detection;

we obtain that the daily death count is proportional to the lagged number of new infectious cases:

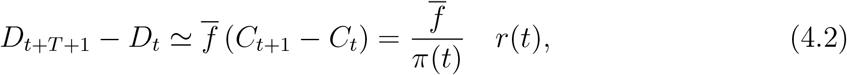

where 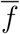 is the (average) infection fatality rate. We use these relations to obtain an estimate for the cumulative number *C_t_* of symptomatic infections and the reporting ratio *π*(*t*).

First, using (4.2) we estimate the average delay *T* between case reporting and death by identifying the lag *T* which maximizes the correlation between the *D_t+T+_*_1_ − *D_t_* and *r*(*t*). As shown in Figure 9, using aggregate fatality counts for England, a maximum correlation *ρ_max_=*95% is observed for a lag of *T* = 8 days between reporting and death. Note that this delay is shorter than the typical recovery period, implying that reporting typically does not occur at the onset of the infection but after a delay of a few days, which is consistent with other studies (for example, Harris [24]). Using an average fatality rate of 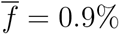 for the UK as in Ferguson et al. [20] (see discussion in Section 3.3), we estimate the reporting probability to be

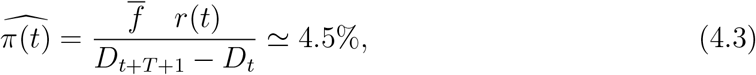

**Figure 9:**
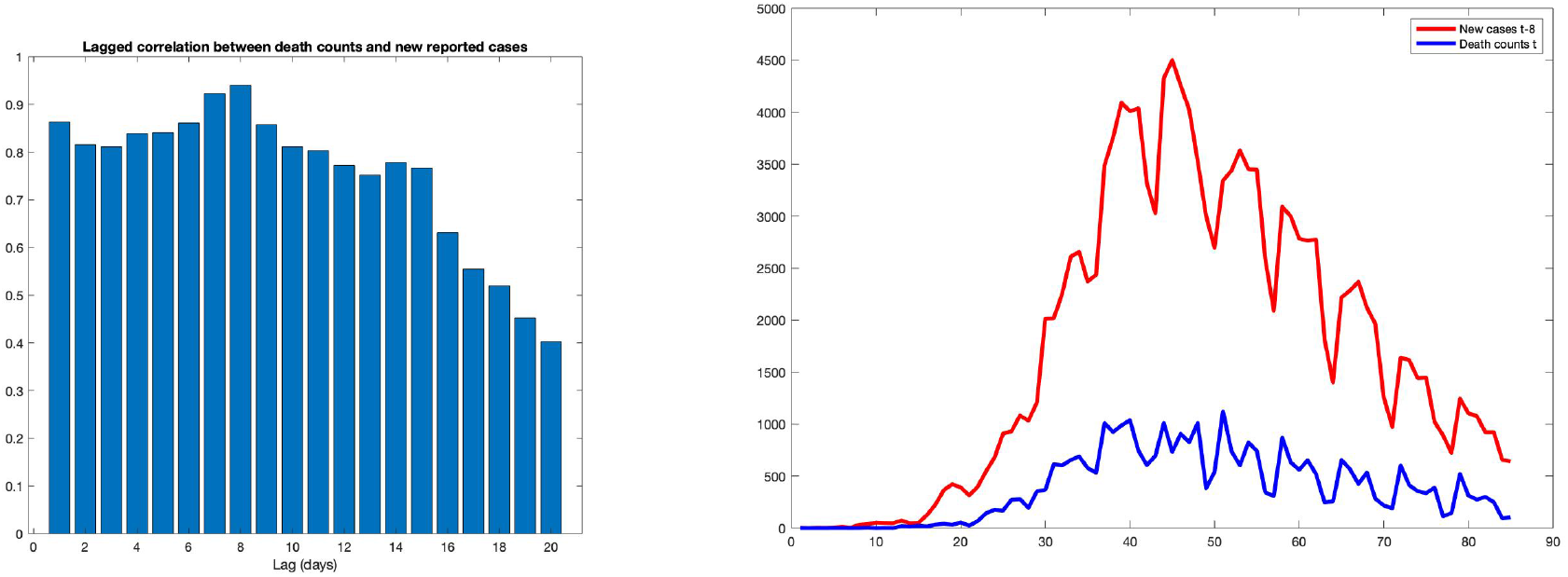
Left: correlation between daily death counts and lagged number of new reported cases (England). A maximum correlation *ρ*_max_ = 95% is observed for a lag of *T* = 8 days between reporting and death. Right: daily death count vs lagged number of new cases (with 8 day lag) for England.

which implies that the total number of cases in England is more than 20 times the reported number. This ratio fluctuates between 0.04 and 0.06 during the observation period, as shown in Figure 10.

**Figure 10:**
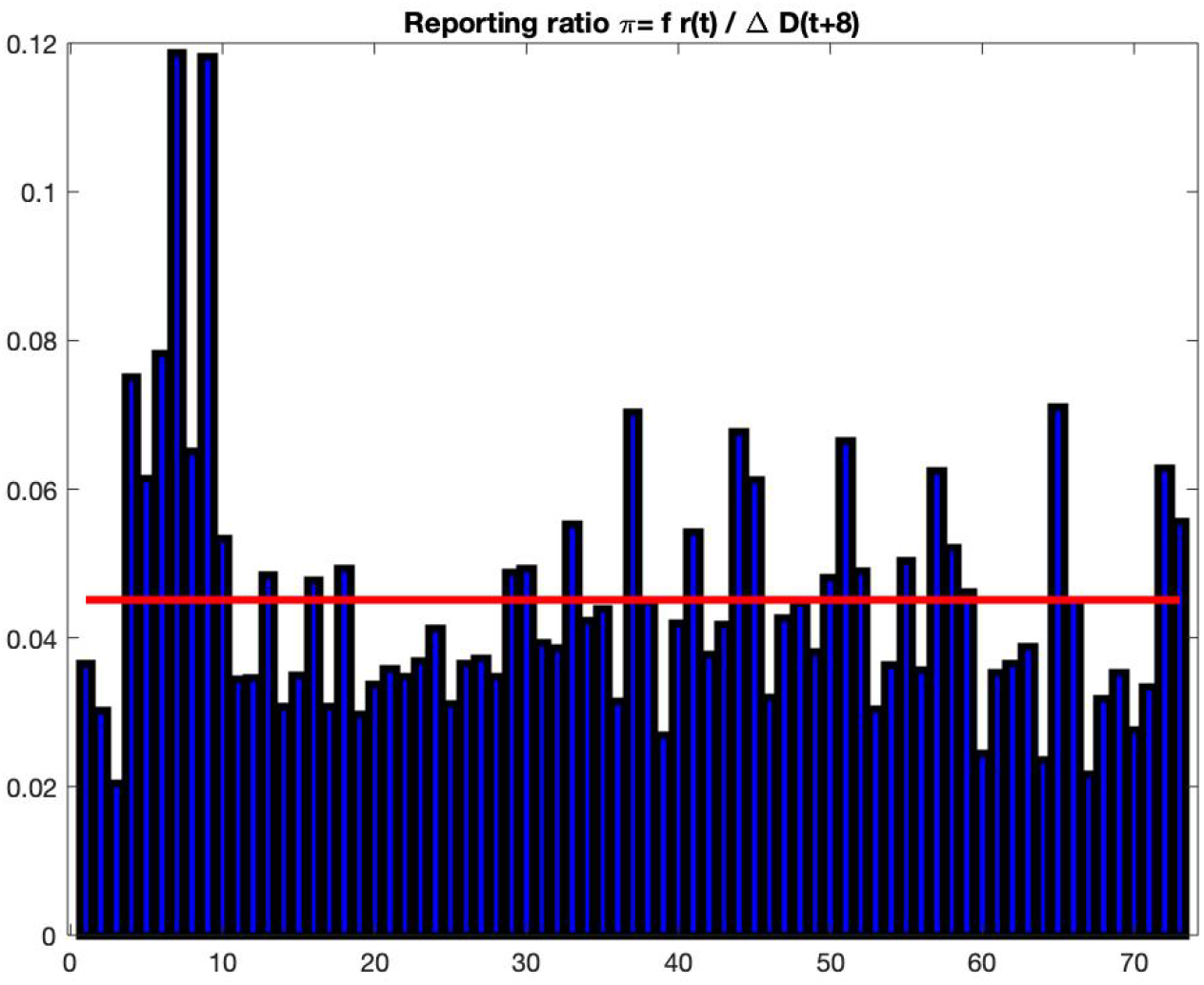
Estimate of the case reporting ratio *π*(*t*) based on fatalities and reported cases. Sample average is 4.5%.

### 4.2 Reliability of reported cases as indicator

A key issue in epidemic control is the availability of reliable indicators for the intensity of an ongoing epidemic. Public health authorities have communicated the daily number of reported cases and fatalities, and these have served as the main inputs for policy planning.

An important corollary of the above discussion is that, given the combination of random factors affecting dynamics and the considerable uncertainty on the actual number of new infections, it is perfectly possible to observe a run of many consecutive days without new reported cases while in fact the *actual* number of infections is on the rise.

Figure 11 shows an example of scenario in our model where, for 100 consecutive days, although a small number of (symptomatic and asymptomatic) cases appear, due to the low detection probability (*π* = 4.5%), none of them is reported. Nevertheless, after a run of 100 days without any reported cases (green shaded area in Fig 11), which would have prompted most public health authorities to lower their guard, the epidemic takes off again. This scenario is not unlike what occurred in South Korea and several other locations in 2020.

**Figure 11:**
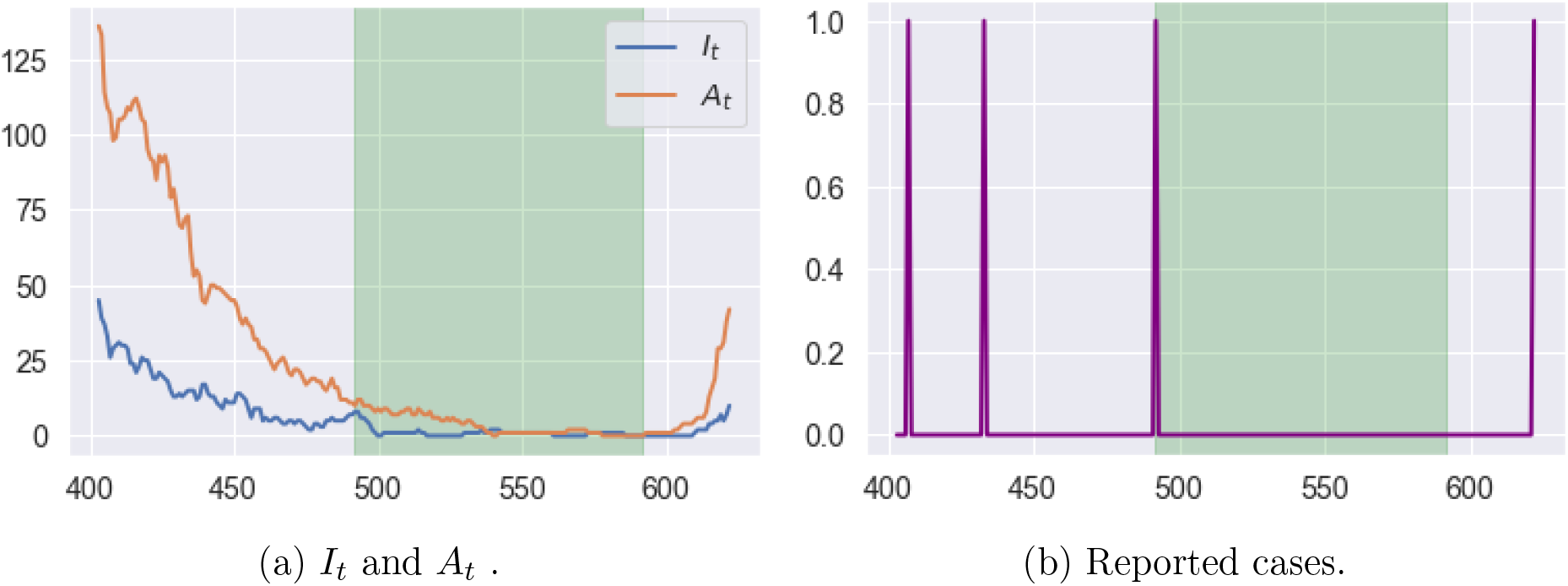
Example of latent progression of the epidemic with zero reported case for 100 consecutive days (green shaded area). On the last day of this period, we have *E_t_* = 0 *I_t_* = 0 and *A_t_* = 1. Reporting probability is *π* = 4:5%.)

Figure 12 shows the probability of observing a subsequent (second) peak in infections if social distancing measures are lifted after no reported cases for *L* consecutive days. This probability is estimated using 500 simulated paths from the model (2.3). It is striking to observe that, even after 100 days with no reported cases, the probability of observing a resurgence of the epidemic is around 40%. Figure 12 (**blue dashed line**) shows the same probability conditional on observing no fatalities for *L* consecutive days.

**Figure 12:**
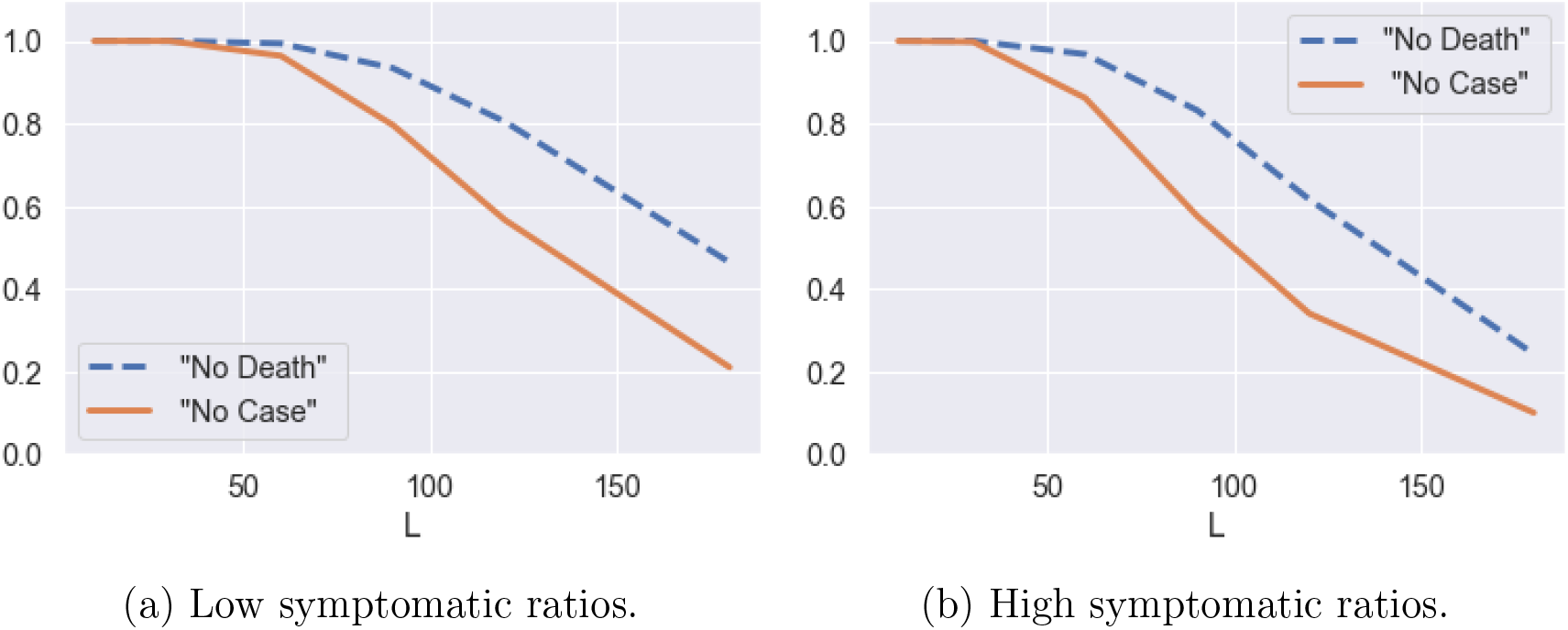
Probability of having a second peak in infections after no reported cases (solid line) and no fatalities (dashed line) for *L* consecutive days. Estimates based on 500 simulated scenarios.

These observations point to the importance of broader testing: as shown in Figure 13, an increase in the probability *π* of detecting new cases leads to a strong decrease in the probability of misdiagnosing the end of the epidemic, as in the scenario described above. In absence of widespread testing, policymakers are faced with the problem of controlling a system under partial observation.

**Figure 13:**
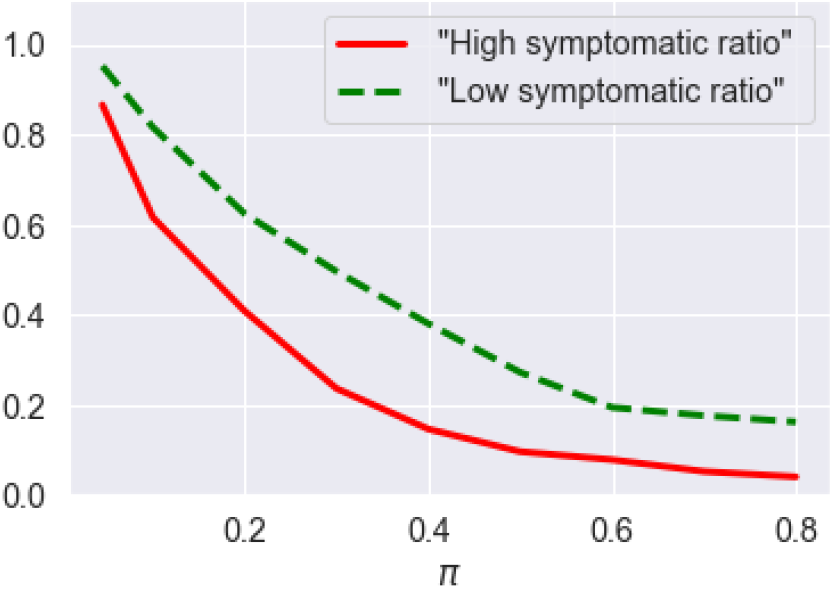
Probability of having a second peak in infections following 60 consecutive days with no reported cases, as a function of reporting probability *π* (after July 1st) (solid line for high symptomatic ratios and dashed line for low symptomatic ratios). Estimates based on average of 500 simulated scenarios.

## 5 Counterfactual scenario: no intervention

A much debated issue has been whether the lockdown and subsequent social distancing restrictions were necessary or whether health outcomes would have been comparable in absence of any restrictions, eventually leading to herd immunity. To examine this question we consider the counterfactual scenario of no intervention and estimate the fatalities and peak number of infections under such a scenario.

### 5.1 Magnitude and heterogeneity of outcomes

Our counterfactual simulations show that, in absence of social distancing and confinement measures, the number of fatalities in England may have exceeded 216,000 by August 1, 2020. This is 174,000 more than the outcome actually observed on this date following the lockdown.

Figure 14 displays the evolution of number of symptomatic infections and fatalities in absence of restrictions, under two different assumptions on symptomatic ratios (see Table 2). Under the assumption of low symptomatic ratios, the total fatalities in absence of any mitigation policy are estimated to be 216, 000 on average across 100 scenarios, with 12.8 million (more than 20% of England’s population) being infected and symptomatic. A peak number of 3, 720,000 symptomatic individuals in England would have been reached on April 30th.

**Figure 14:**
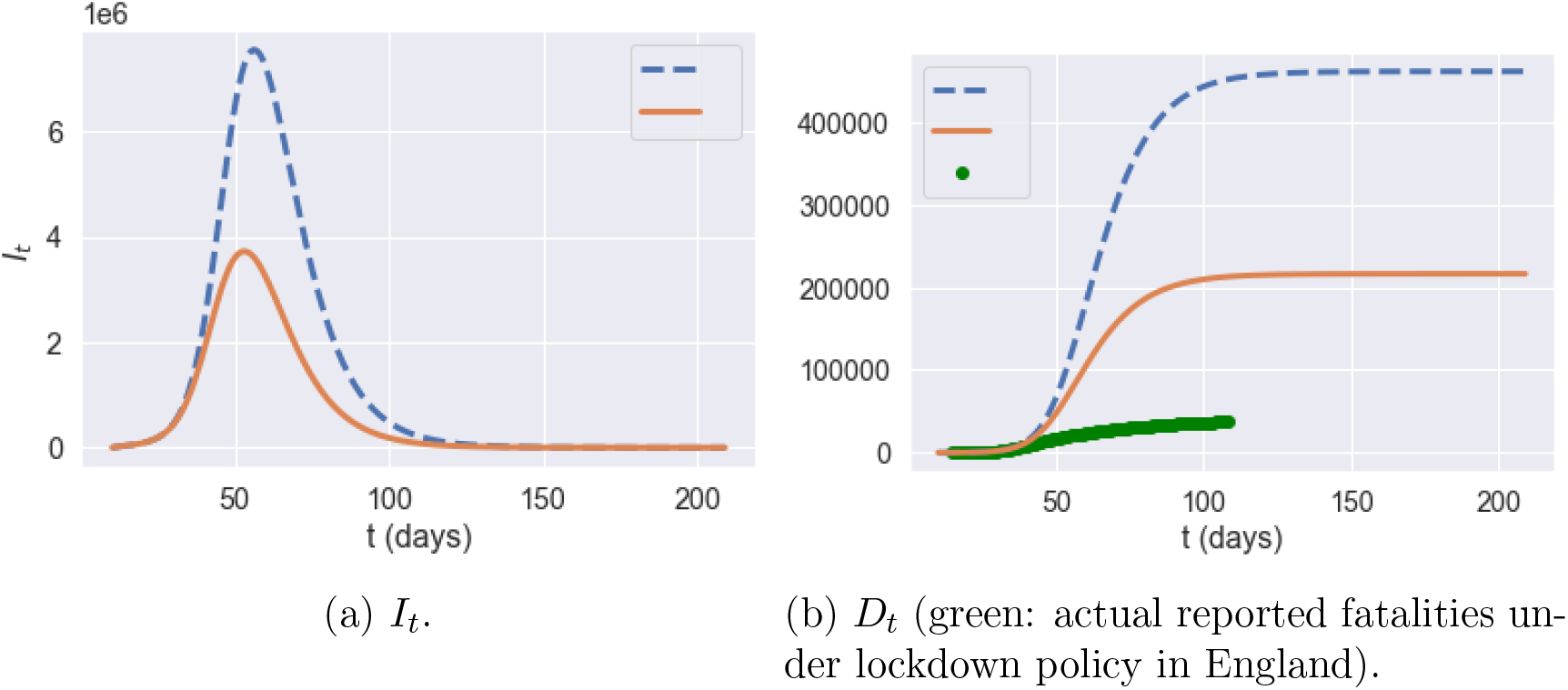
Comparison of different quantities in England with no intervention: high symptomatic ratios (**blue** dashed line) versus low symptomatic ratios (**orange** solid line), averaged across 100 simulated scenarios.

Figure 15 decompose these results across different age groups.

**Figure 15:**
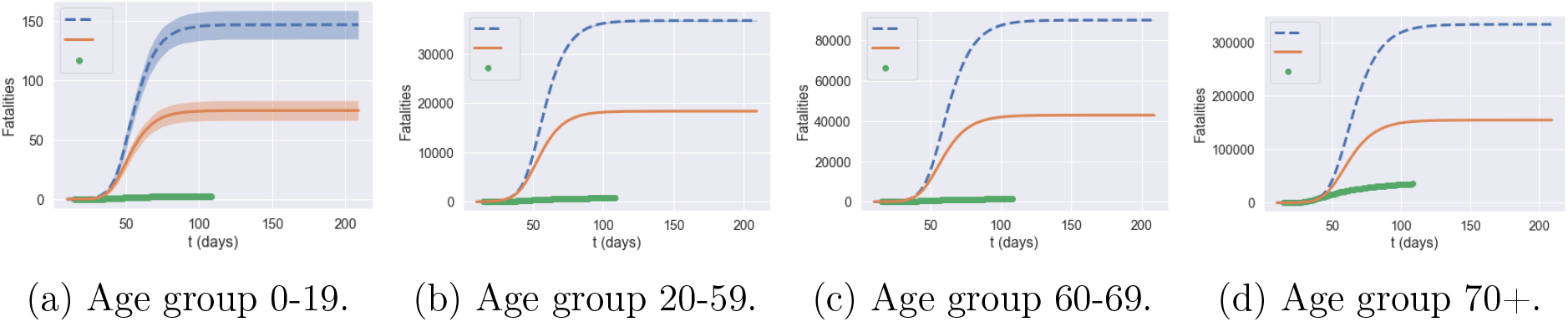
Fatalities by age group under no interventionm averaged across 100 scenarios: high symptomatic ratios (**blue** dashed line) versus low symptomatic ratios (**orange** solid line). Green dots: actual reported fatalities in England under lockdown policy.)

The low and high estimates for symptomatic ratios lead to very different simulation outcomes in terms of peak *I_t_* values and total death numbers, which illustrates the huge impact of parameter uncertainty on model projections.

#### Heterogeneity of regional outcomes

Regions exhibit heterogeneous outcomes in terms of peak time, peak value, and fatalities (per 100,000 inhabitants).

### 5.2 Impact of demographic and spatial heterogeneity

Homogeneous SIR models [3, 36, 17, 29, 42, 48, 45, 49] or age-stratified versions of such models [1, 15, 16, 44, 52] have been used in many recent studies on COVID-19 in the UK and other countries.

The heterogeneity of outcomes observed in our simulation suggests that homogeneous epidemic models may fail to capture some important features of COVID-19 dynamics which are relevant for public health policy. We investigate this point further by comparing our simulations with two homogeneous benchmark: a country-level SEIAR compartmental model and an age-stratified version of it.

#### Homogeneous SEIAR model

The homogeneous SEIAR model corresponds to the case where the country is considered as a single region, assuming away geographic heterogeneity:

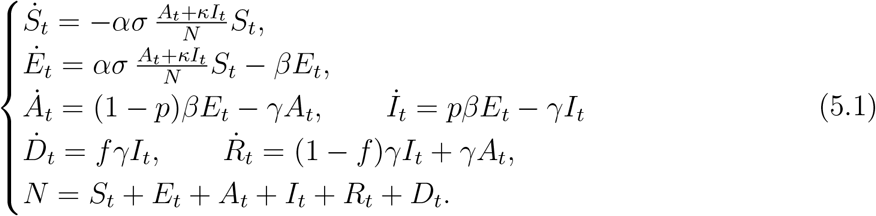

where *σ* is the average contact rate in the population and *N* is the total population.

We use the values of *α, β* and γ as specified in Table 3 and population-averaged versions of low symptomatic ratios in Table 2. We aggregate the POLYMOD age-stratified contact matrix (3.2) from POLYMOD, the probability of developing symptom, and fatality rate using the England population age distribution (Table 1) leading to an aggregate fatality rate of 1.1%, probability of developing symptoms of 45.4%, and contact number of 4.157. As in Sections 3.3 and 3.5, we use an indirect inference method [22] to estimate the implied scaling parameters for social contact rates by matching daily case dynamics before and during lockdown, leading to *d* = 2.909 and *l* = 0.115.

We also consider an age-stratified (**A**) version of this model with 4 age groups. The rate of exposure for age group *a* is given by

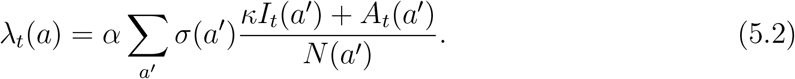

For the age-stratified model, we use *α, β* and γ as in Table 3. We estimate the scaling parameters for social contact rates before and during lockdown as above, yielding *d* = 1.07 and *l* = 0.112.

As shown in Figure 17, these country-level models actually yield reasonable fits to aggregate dynamics of cases and fatalities.

**Figure 16:**
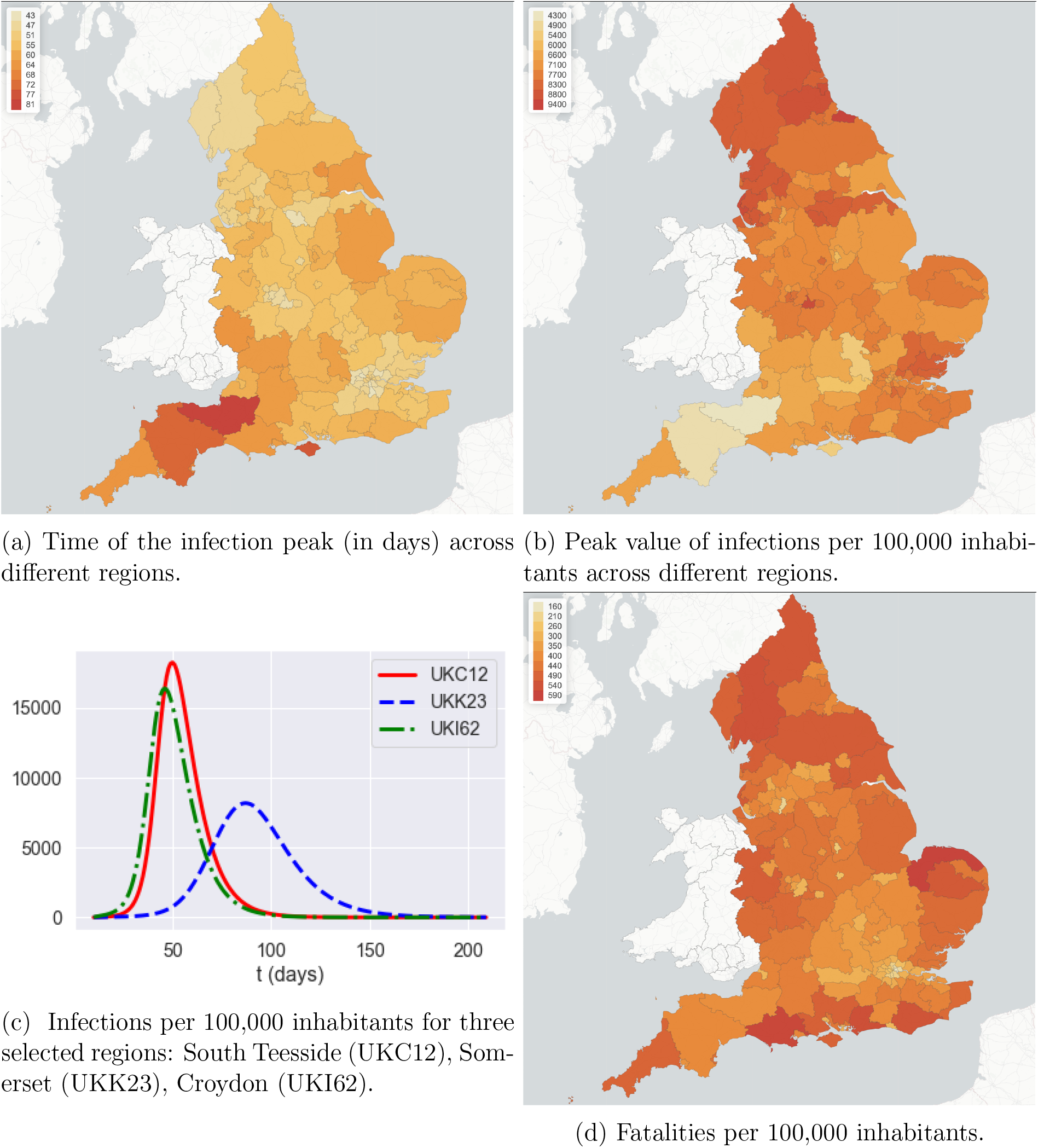
Heterogeneity of outcomes across different regions, in absence of intervention. Outcomes are averaged over 100 simulated scenarios.

**Figure 17:**
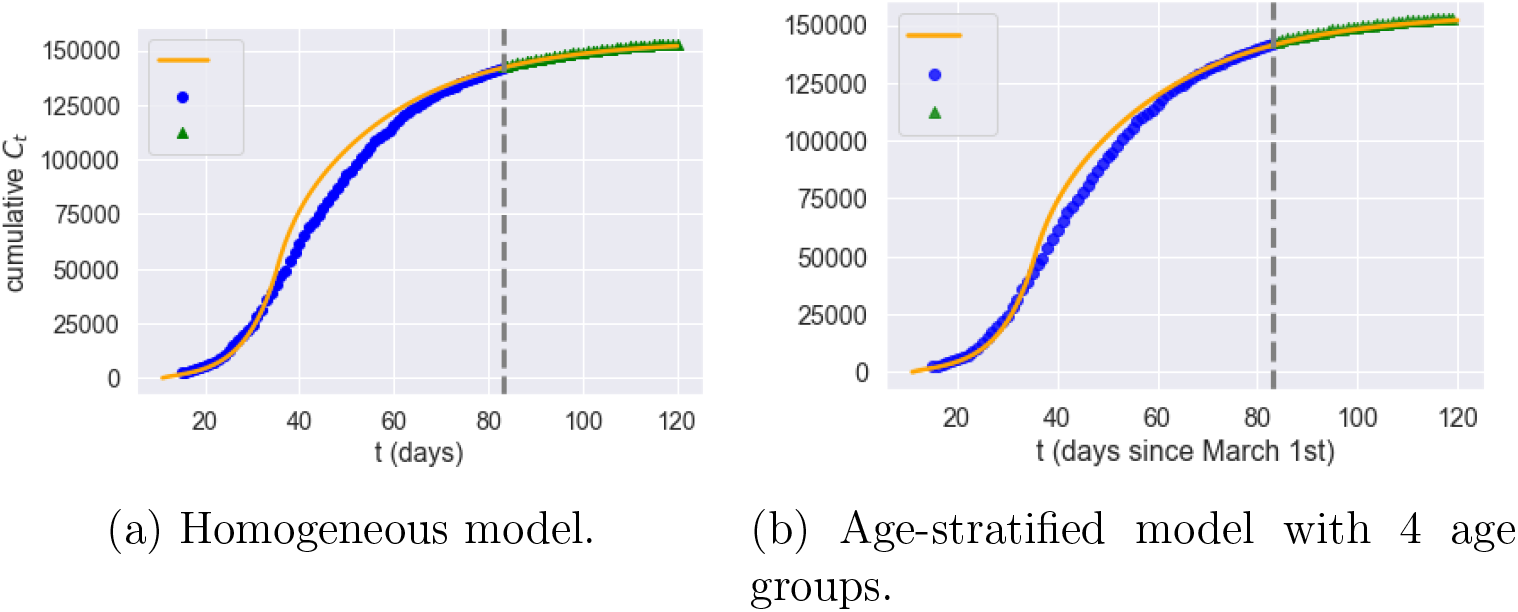
Homogeneous and age-stratified country-level models: goodness of fit (Grey dashed line: separation of estimation sample from test data; orange line: model; blue dot: data; green triangle: out-of-sample data).

**Figure 18:**
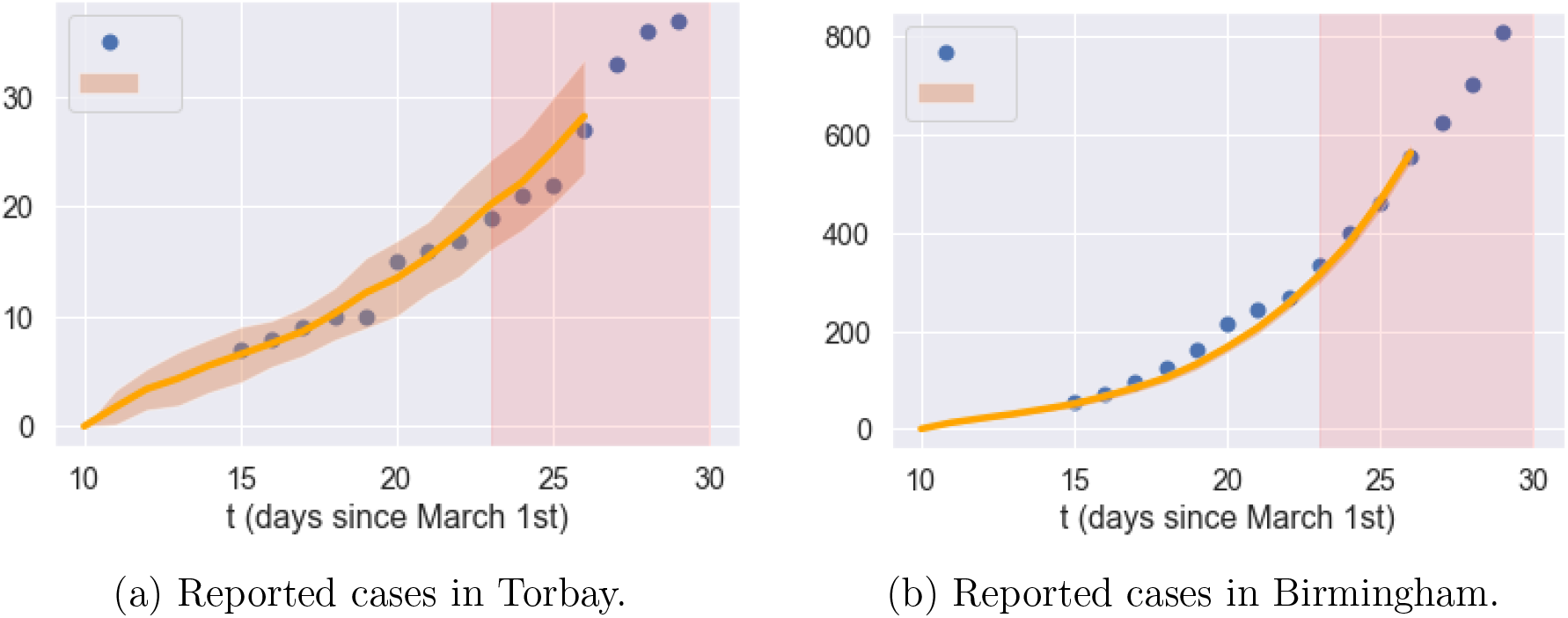
Dynamics of reported cases before lockdown in Torbay and Birmingham: data compared with heterogeneous model (**F**). Orange line: average of 1000 simulated scenarios; pink shade: lockdown period.

However, the inability of these country-level models to capture regional heterogeneity and inter-regional exchanges leads to regional outcomes which are very different from our model with spatial heterogeneity (**F**). Figures 19-22 compare simulation results for the homogeneous model (**H**), the age-stratified model (**A**) and our spatial heterogeneous model (**F**) in two regions: Torbay (UKK42) and Birmingham (UKG31). The homogeneous model is observed to over-estimate fatalities in age groups 1 and 2 and underestimates fatalities in age groups 3 and 4. The age-stratified model and the heterogeneous model agree on fatalities across age groups but, as shown in Figures 19-22, neither the age-stratified model nor the homogeneous model can capture the regional features of epidemic dynamics, such as the difference in peak time and peak value of infections across regions.

**Figure 19:**
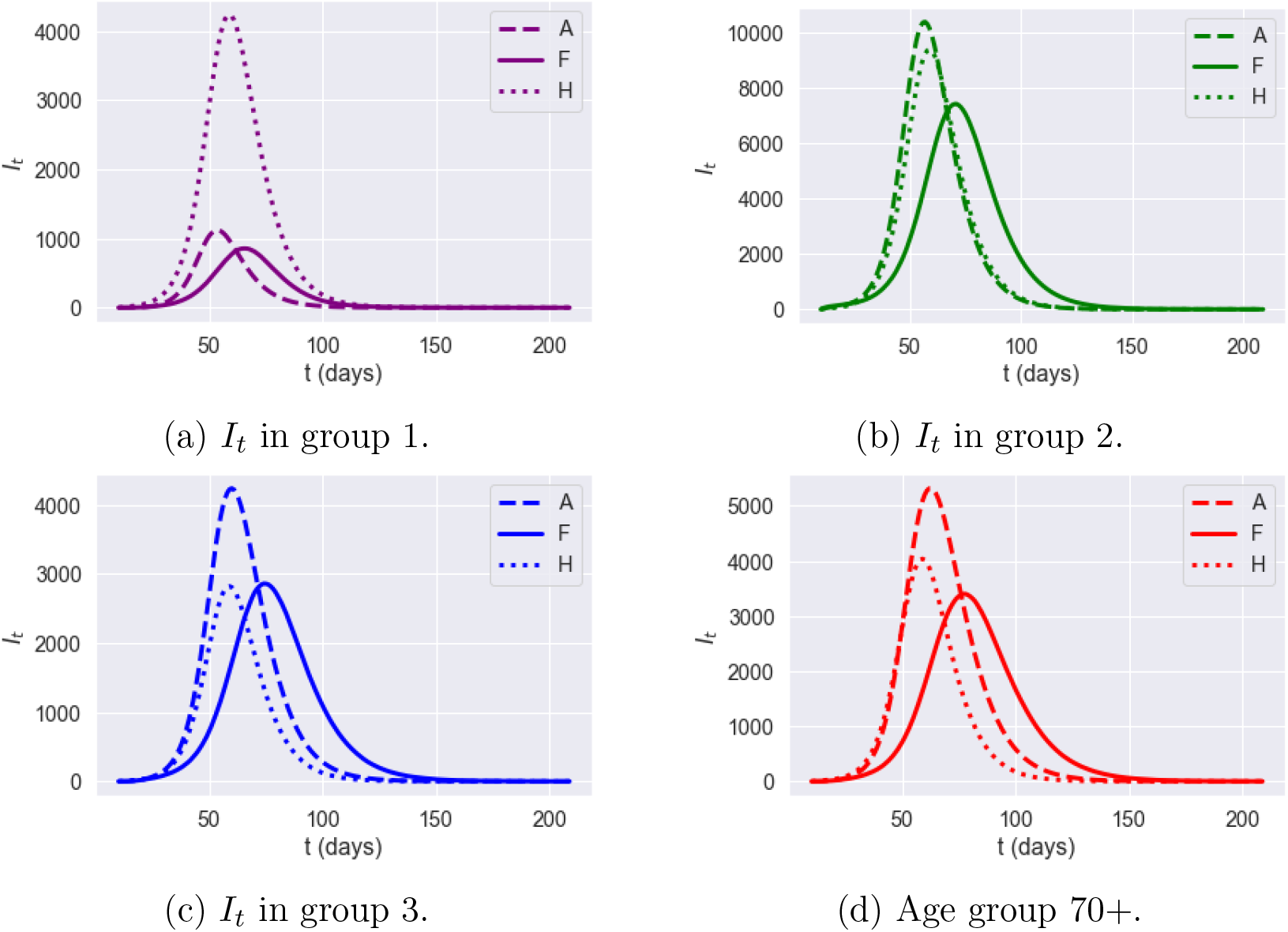
Dynamics of infections *I_t_* in Torbay (UKK42): impact of model granularity.

**Figure 20:**
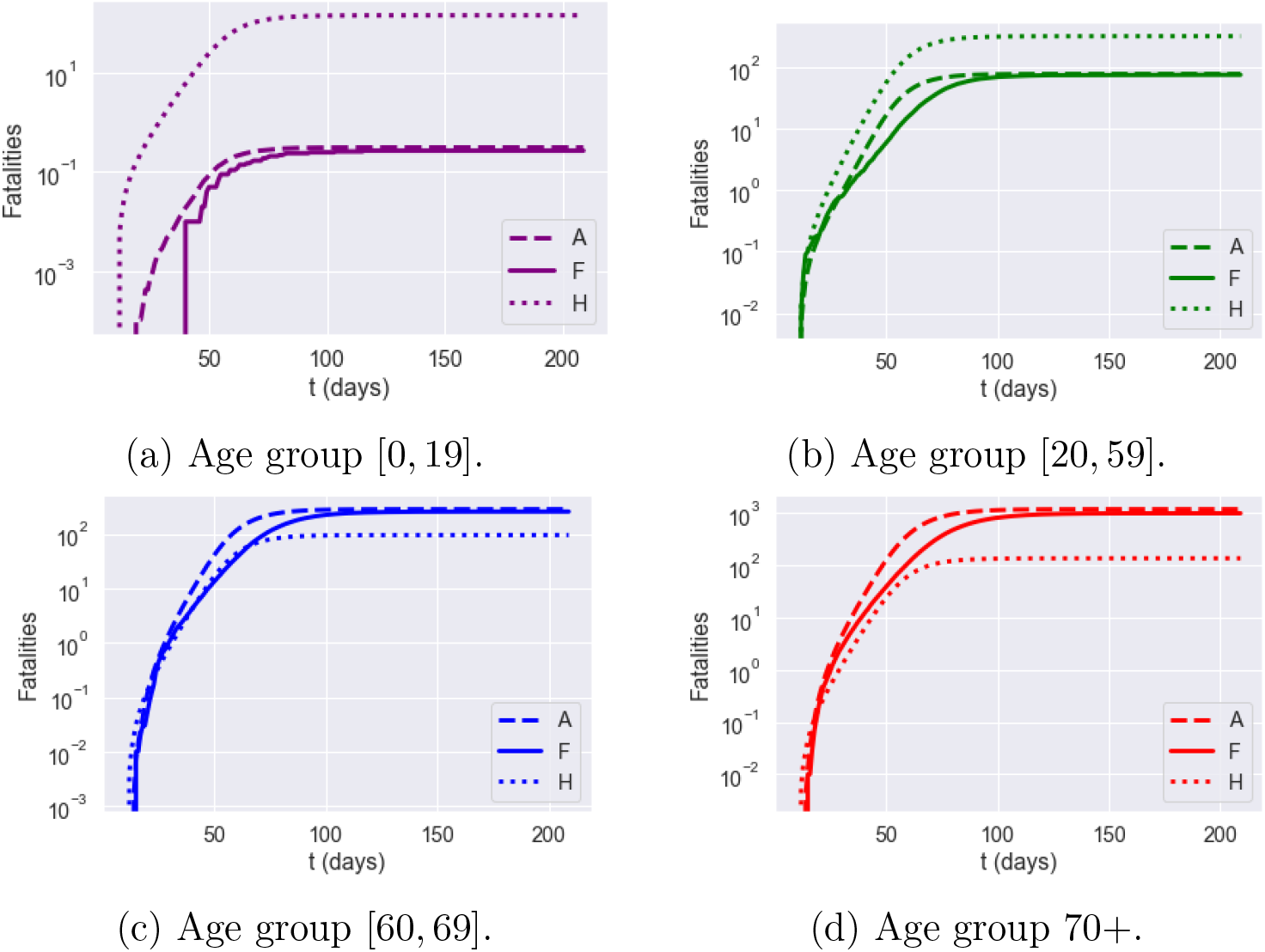
Dynamics of fatalities *I_t_* in Torbay (UKK42): impact of model granularity.

**Figure 21:**
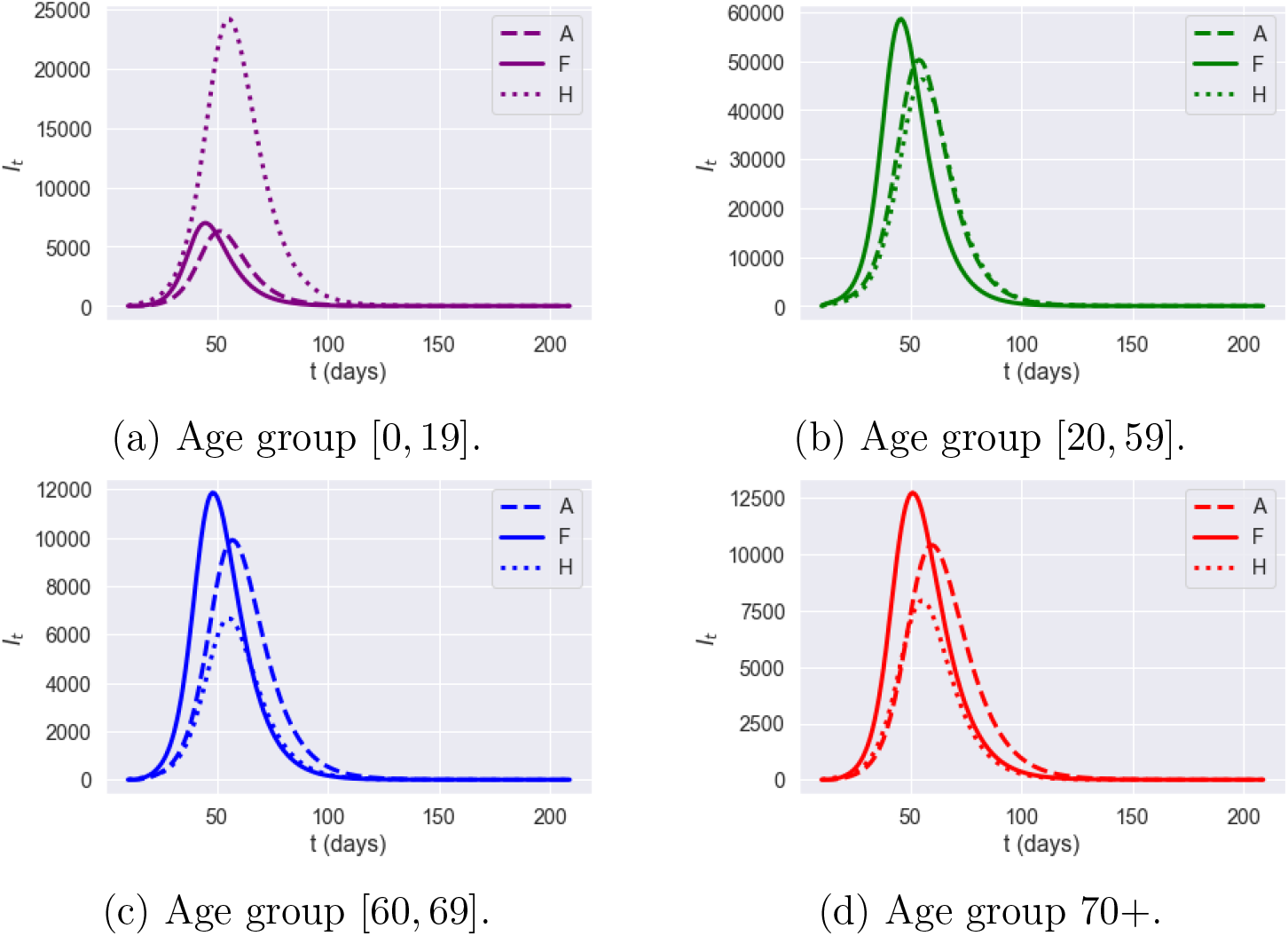
Dynamics of symptomatic infections (*I_t_*) in Birmingham (UKG31): impact of model granularity.

**Figure 22:**
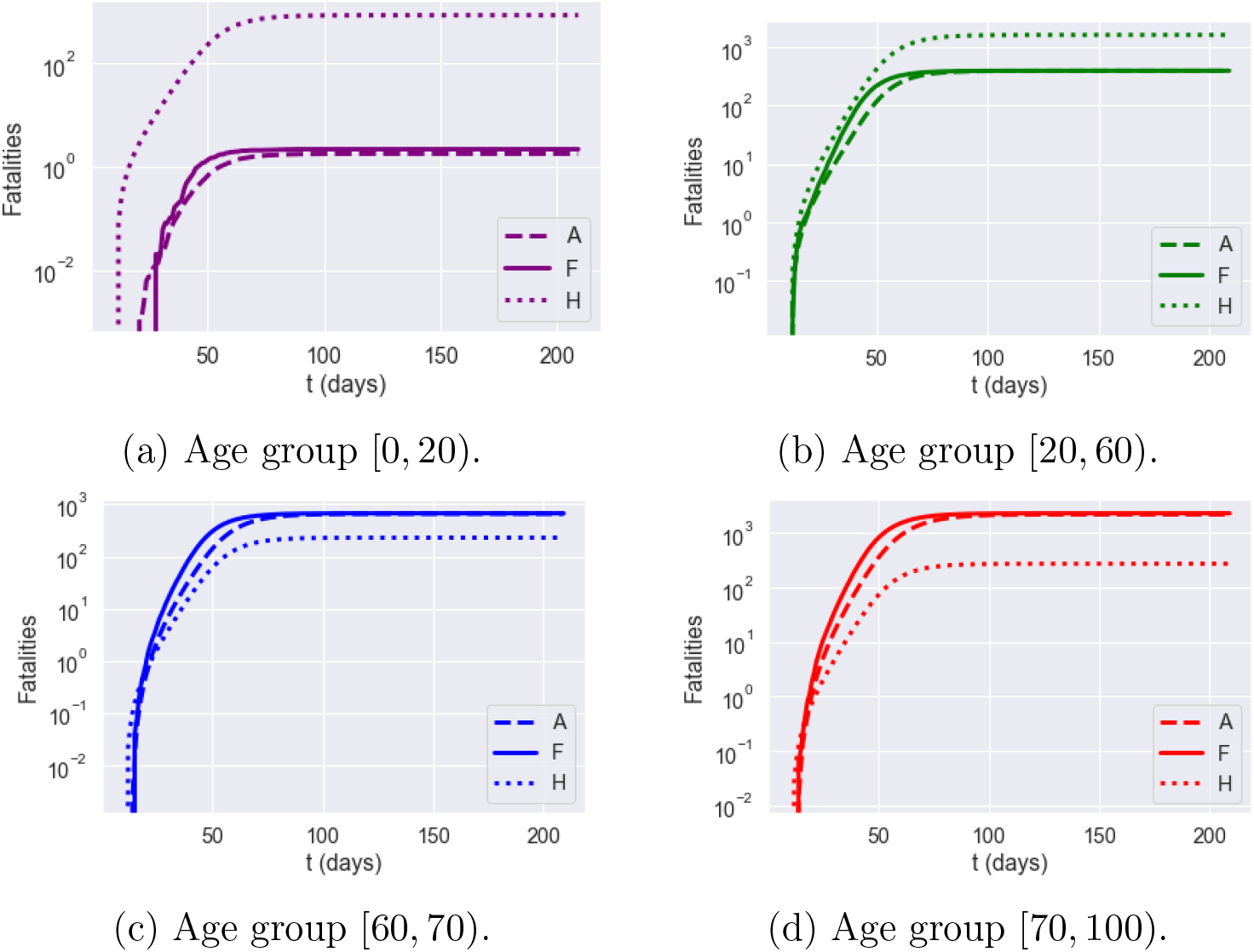
Cumulative fatalities for Birmingham (UKG31): impact of model granularity.

We therefore conclude such country-level models should not be used in the context of policy discussions which target regional measures or regional outcomes.

### 5.3 Variability of outcomes

Given a set of initial conditions, the stochastic model (2.3) may lead to a range of outcomes due to the randomness present in the dynamics. Figures 23-24 show an example of variability of outcomes across different scenarios. As expected from asymptotic analysis of large population limits [12], the relative variability across scenarios is of order 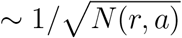 for a sub-population of size *N*(*r,a*). Although not negligible, especially at the onset of the epidemic, this variability remains small at the regional level given the granularity used in our model. In the sequel we have thus reported the average outcomes across 100 or 1000 simulated scenarios for each case examined.

**Figure 23:**
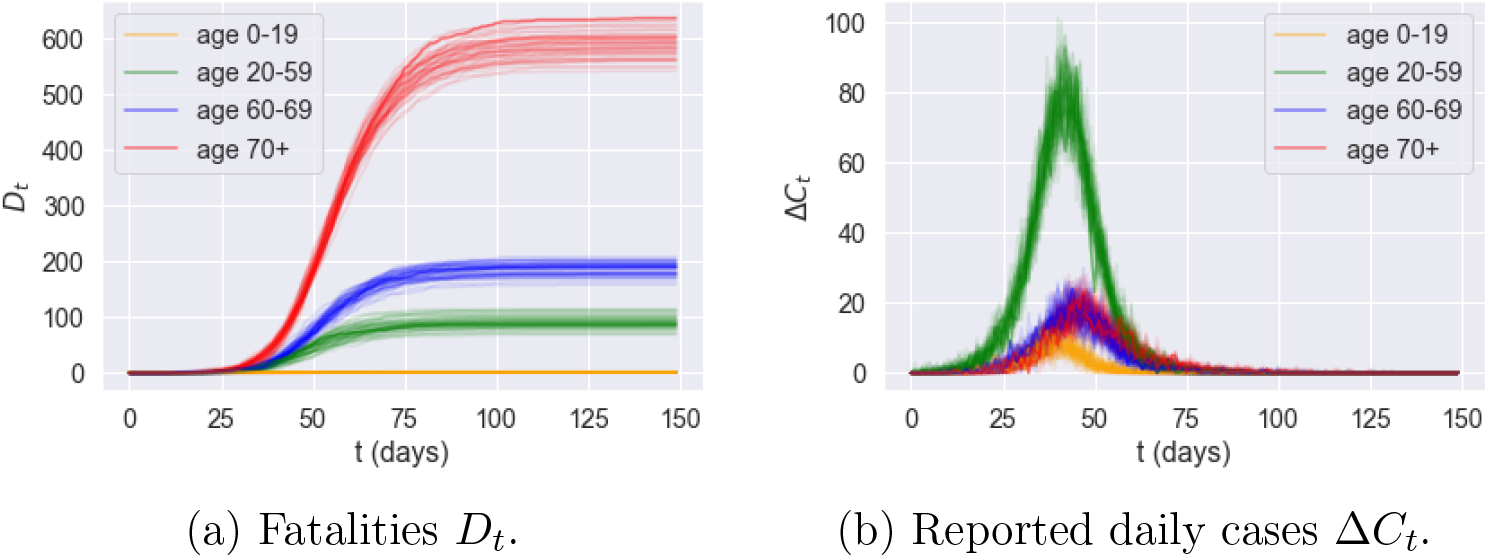
Effect of randomness: variability of outcomes across 50 sample paths for Kingston upon Hull (UKE11). (Low symptomatic ratios.)

**Figure 24:**
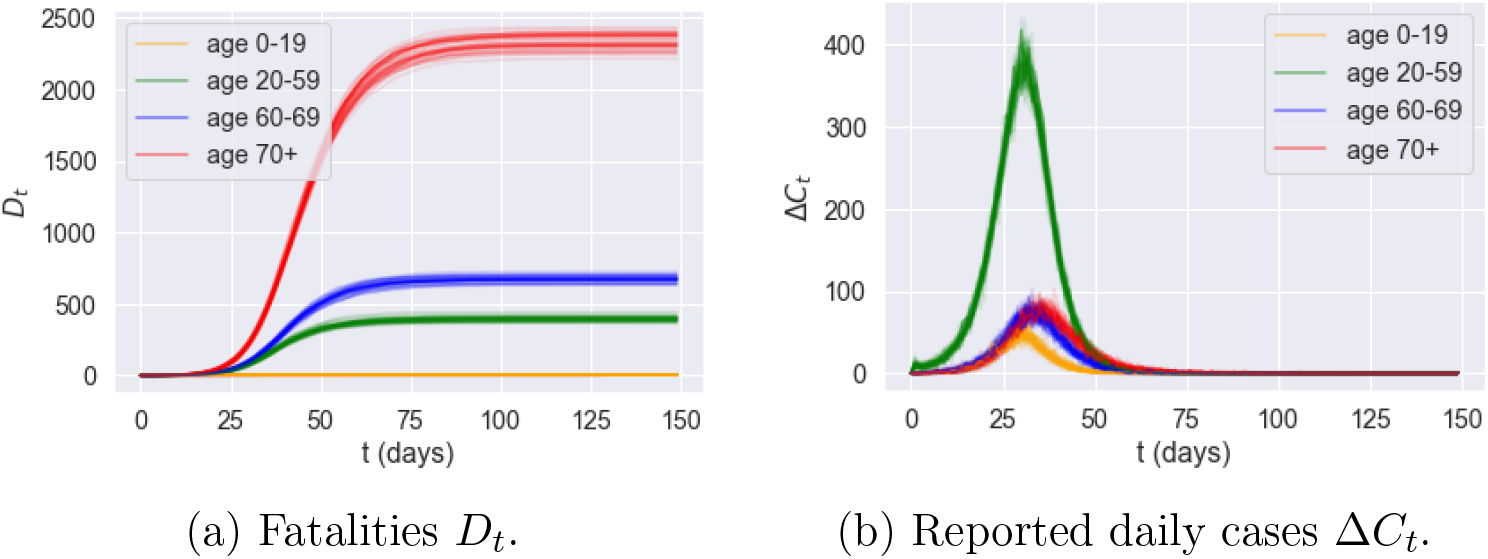
Effect of randomness: variability of outcomes across 50 sample paths for Birmingham (UKG31). (Low symptomatic ratios.)

## 6 Comparative analysis of epidemic control policies

### 6.1 Confinement followed by social distancing

We first consider the impact of a ‘lockdown’ followed by social distancing, which reflects the situation in the UK since end March 2020. We examine in particular the impact of a lockdown duration *T* and the level of social distancing after lockdown on the number of fatalities and the associated social cost. To do so, we parameterize the contact matrix as

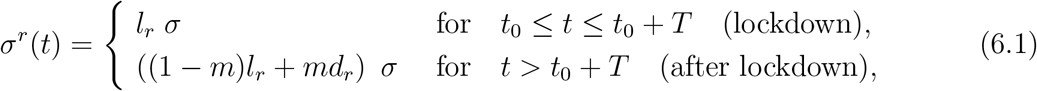

where *l_r_* measures the level of social distancing under lockdown, as estimated from observations for the period from Mar 23 to May 31, and the parameter *m* ∈ [0,1] measures the level of compliance with social distancing measures. A value of *m* close to zero indicates a level of social contact similar to lockdown, while *m* = 1 corresponds to normal levels of social contact.

*t* = 0 corresponds to March 1, 2020. All scenario simulations correspond to a lockdown starting at *t*_0_ = March 23, 2020. We consider a range 105 ≤ *T* ≤ 335 for the lockdown duration and 0.2 ≤ *m* ≤ 1 for post-lockdown compliance levels. Note that the actual lockdown duration in England corresponds to *T* = 105.

As shown in Figure 25a, the level of social distancing *after* the confinement period is observed to be more effective in controlling the epidemic (Figure 25b), than extending the period (Figure 25a). This is consistent with the findings in Lipton and Lopez de Prado [34]. Smaller values of *m*, associated with stricter social distancing, lead to a lower of fatalities but for at an increased social cost (Figure 25b). On the other hand, the lengthening of the lockdown duration *T*, while significantly increasing the associated social cost, does not result in a significant reduction in the number of fatalities, especially if social distancing after lockdown is relaxed.

**Figure 25:**
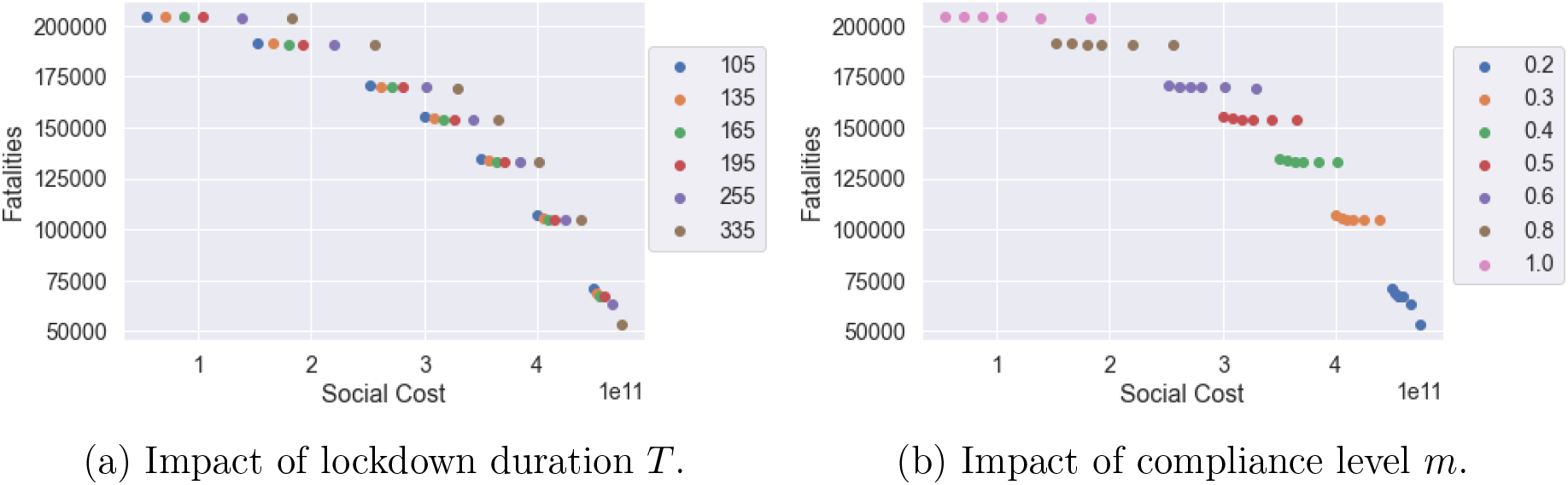
Fatalities against social cost for different T and *m* values (results for low symptomatic ratios).

Figure 25 also shows that some of these policies are inefficient, in the sense that we can reduce fatalities *and* the social cost simultaneously by shortening the lockdown period or by relaxing social distancing constraints, as shown in Figure 26.

**Figure 26:**
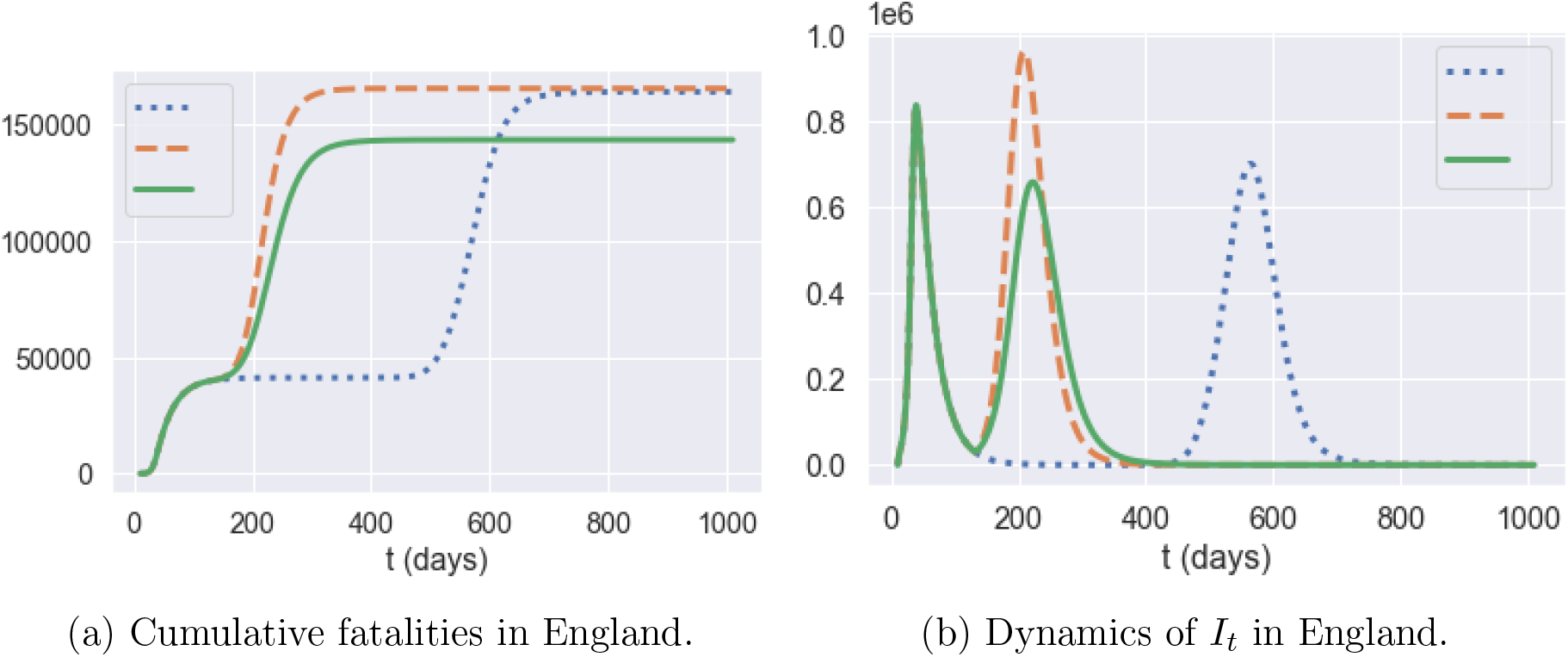
Comparison of three policies: **blue dotted** line: *m* = 0:5 and *T* = 335; **orange dashed** line: *m* = 0:5 and *T* = 105; **green solid** line: *m* = 0:4 and *T*_2_ = 105. (Results for 50 simulated scenarios.)

By comparing the **orange** and **blue** plots, which represent the same post-lockdown compliance level (*m* = 0.5), we observe that extending the lockdown duration increases social cost without reducing the total number of fatalities. On the other hand, comparing the **orange** and **green** plots, which correspond to the same lockdown duration of *T* = 105 days, shows that moving the compliance level from *m* = 0.5 to *m* = 0.4 reduces the second peak amplitude by 35% and fatalities by 12.5%.

#### Impact of parameter uncertainty

The above results are highly sensitive to the value of the symptomatic ratios which, as noted in Section 3, are highly uncertain (see Table 2). Figure 27 shows the policy outcomes for low versus high symptomatic ratios across different compliance levels and lockdown duration. As observed in this figure, while the overall pattern of the efficiency diagram is similar, the projected fatality levels shift considerably depending on the assumption on the symptomatic ratio: from 70,000-200,000 for low symptomatic ratios to 170,000-430,000 for high symptomatic ratios.

**Figure 27:**
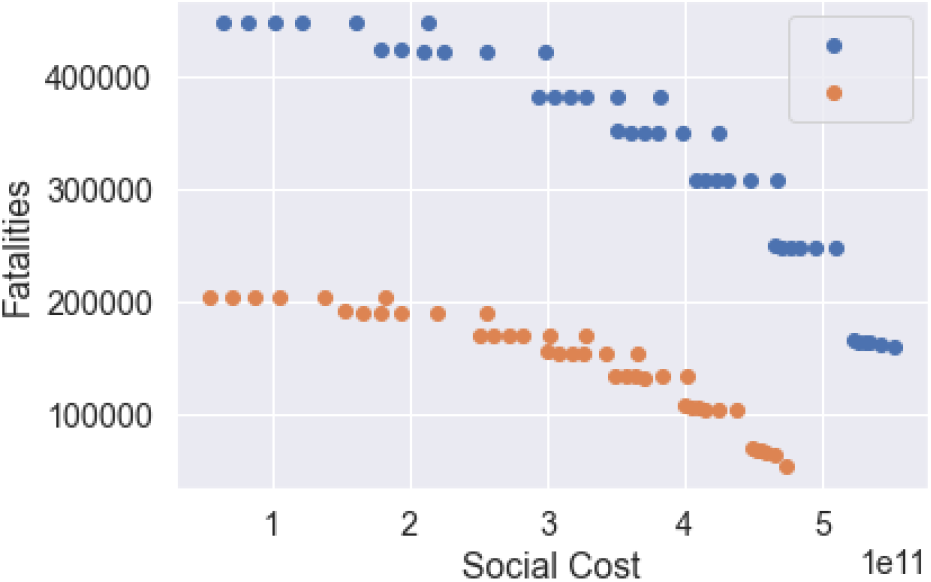
trade-off between fatalities and social cost for T-day lockdown followed by social distancing (0:2 ≤ *m* ≤ 1, 105 ≤ *T* ≤ 335): low symptomatic ratio (**orange**) and high symptomatic ratio (**blue**).

#### Regional heterogeneity

While the policies discussed here are applied uniformly across all regions, we observe a significant heterogeneity in mortality levels across regions, as well in terms of the timing and amplitude of a second peak tin infections. As shown in 28, some regions exhibit mortality levels up to 4 times higher than others. This huge disparity in mortality rates cannot be explained by demographic differences alone, which are much less pronounced: more important seem to be the differences in social contact patterns, as illustrated in Figure 2. Indeed, as shown in Figure 29a, there is a positive correlation (> 40%) between regional COVID-19 mortality and the intensity of social contact as measured by the parameter *d_r_* defined in Sec. 3.3.

**Figure 28:**
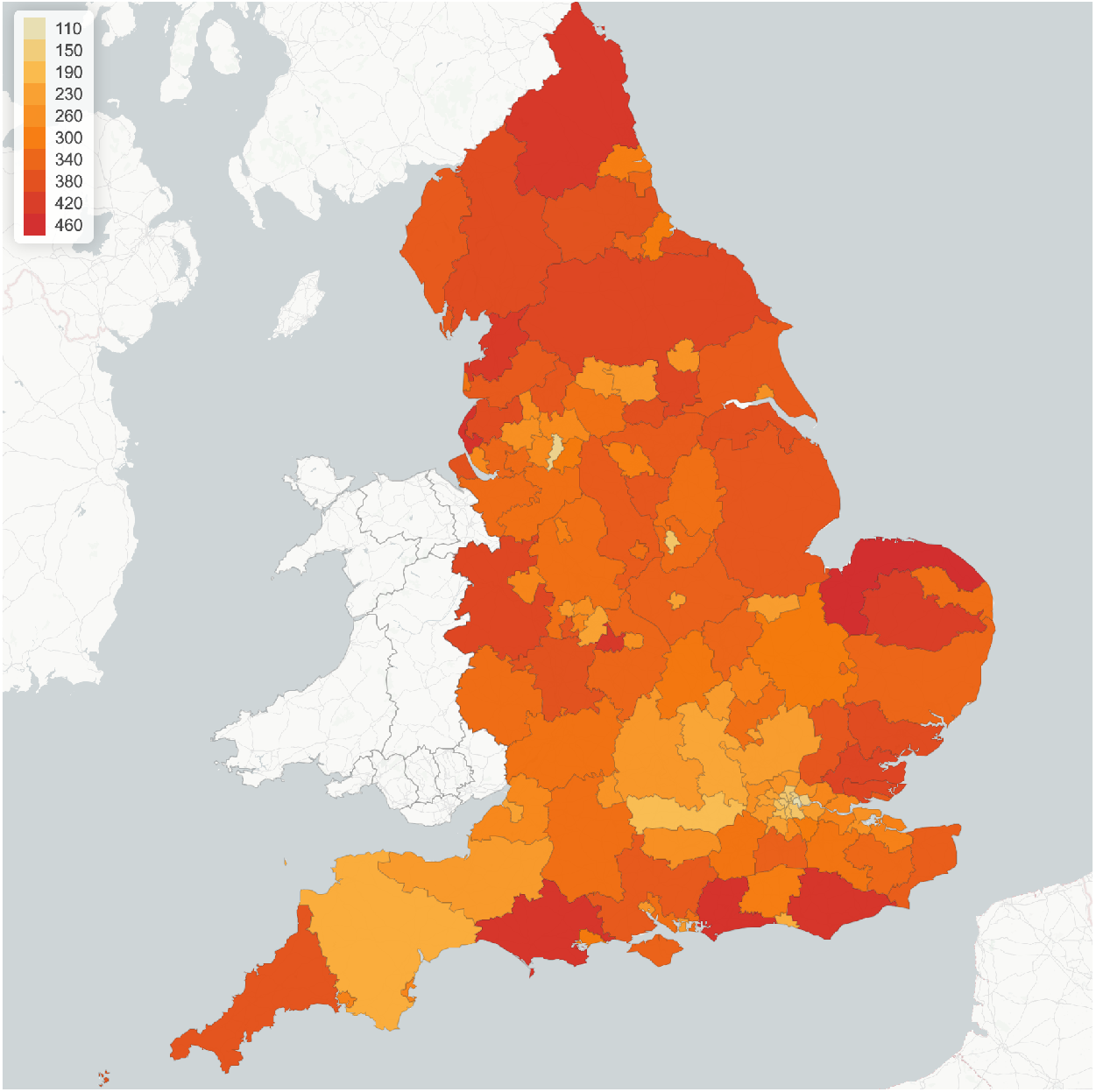
Lockdown of 105 days followed by social distancing (*m* = 0:5): regional mortality per 100,00 inhabitants.

**Figure 29:**
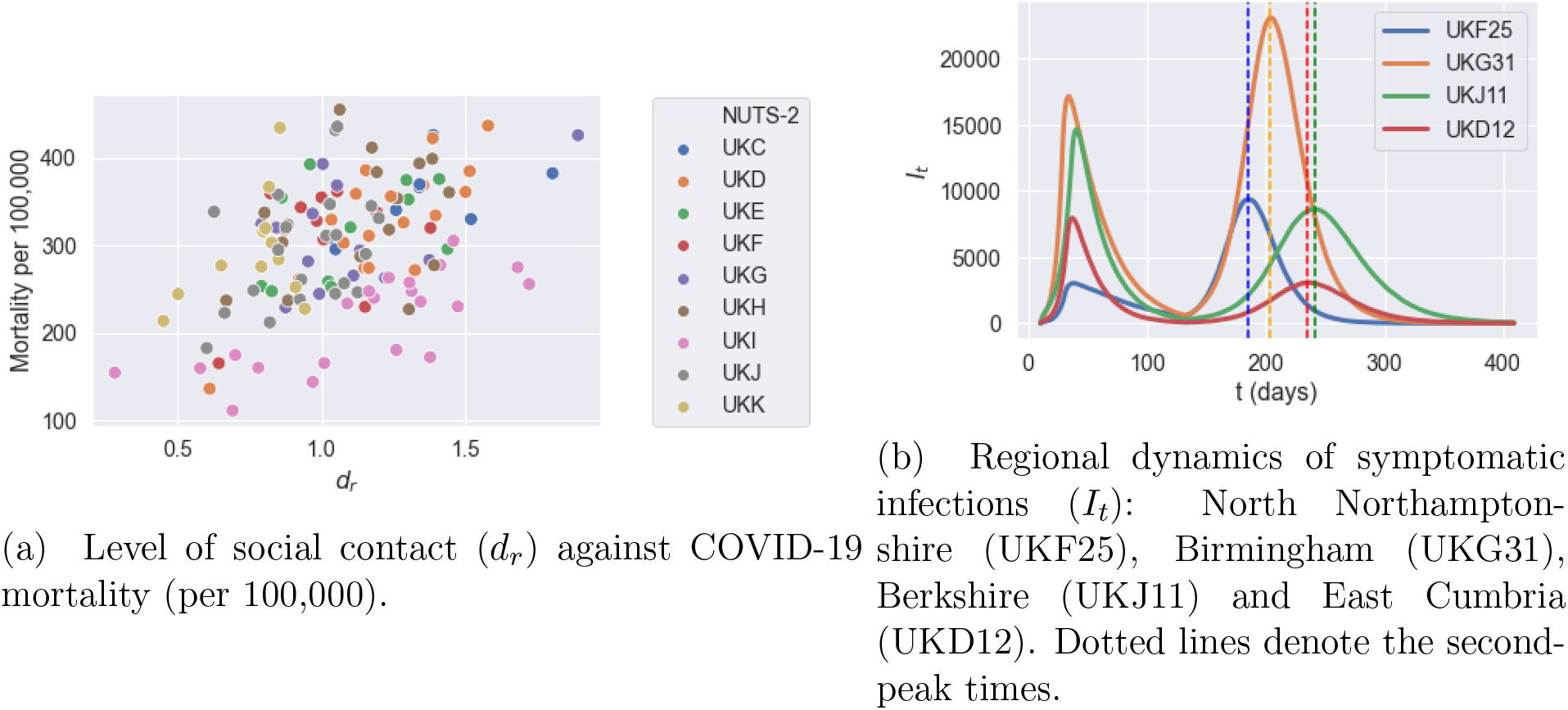
Regional outcomes for lockdown of 105 days followed by social distancing (m = 0:5).

Figure 29b shows that this heterogeneity is also reflected in the timing and amplitude of second peaks. We observe that East Cumbria has a small second peak compared to its first one, Birmingham and Berkshire experience a second peak with a similar size compared to the first one, while North Northamptonshire suffers from a much more severe second peak. The second peak in North Northamptonshire and Berkshire occur around *t* = 175 and *t* = 231, respectively.

### 6.2 Targeted policies

We now consider the impact of social distancing measures targeting particular age groups or environments (school, work, etc.) following a lockdown of duration *T*, by setting

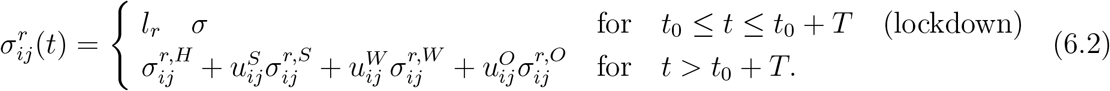

We consider different targeted measures after a lockdown period of *T* = 105 days (the actual duration of the lockdown in England): school closure, shielding of elderly populations and workplace restrictions, restrictions on social gatherings and combinations thereof. Note that there is no control over the social contacts at home.

#### Closure of schools

Although most of the infected population in age group 1 is asymptomatic, they may in turn infect the population in age groups 3 and 4 who are more likely to develop symptoms. School closure corresponds to *u^S^* = 0, school reopening with social distancing correspond to *u^S^* = 0.5, and school reopening without social distancing correspond to *u^S^* = 1.

#### Shielding

The high infection fatality rates among elderly populations (age groups 3 and 4) have naturally lead to consider shielding policies for these populations. We model this as a reduction in social contacts of age groups 3 and 4 to the level observed under lockdown:

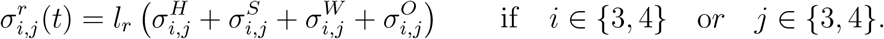

#### Workplace restrictions

We model the impact of a restricted return to work after confinement by assuming different proportion of workforce return after the lockdown period by choosing

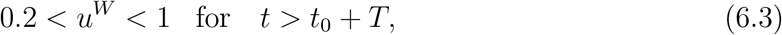

the lower bound *u^W^* = 0:2 corresponding to restricting workplace return to ‘essential workers’, as discussed in Section 3.5. Since workplace restrictions have an effect on commuting, such measures also have an impact on the inter-regional mobility matrix

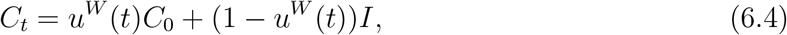

where *C*_0_(*r; r′*) is the baseline mobility matrix defined in (3.1).

#### Restrictions on social gatherings

Although social activities, such as gatherings at pubs or sports events, may aggravate the contagion of COVID-19, keeping certain levels of social activities is important to the economic recovery and the well-being of individuals. The parameter *u^O^* measures the fraction of social gatherings: during the lockdown this fraction was estimated to be as low as 20% (see Section 3.5). In what follows, we consider *u^O^* ∈ [0.3; 1.0] after the period of lockdown.

and various values of *u^S^, u^W^*, and *u^O^* (*u^H^* = 1 and *T* = 105).

#### 6.2.1 Pubs and schools

Table 6 show the impact of school closures and social distancing at schools on projected fatalities and social contacts. Reopening of schools, while reducing significantly the social cost, does not seem to lead to a significant increase in fatalities.

**Table 6:** Impact of school closures and social distancing at schools: outcomes averaged across 50 simulated scenarios, *u^H^* = *u*^W^ = 1, *u*^O^ = 0.5.

| | School closure<br>$u^S = 0$ | Social distancing at school<br>$u^S = 0.5$ | Normal school regime<br>$u^S = 1.0$ |
| --- | --- | --- | --- |
| Social cost ($10^{11}$) | 2.3 | 2.0 | 1.6 |
| Projected fatalities | 163849.0 | 166319.3 | 168380.2 |

We now compare two post-confinement policies, one (labeled ‘Schools’) consisting in leaving schools open while social gatherings (‘pubs’) are restricted (*u^S^* = 1, *u^O^* = 0.2), and the other (labeled ‘Pubs’) consisting in closing schools while not restricting social gatherings (*u^S^* = 0, *u^O^* = 1). The social cost for the ‘Pubs’ policy is 2.4, while the cost for the ‘Schools’ policy is 3.1. However, as shown in Figure 31, the ‘open school’ policy leads to 35% fewer fatalities 35% compared to the ‘open pubs’ policy.

**Figure 30:**
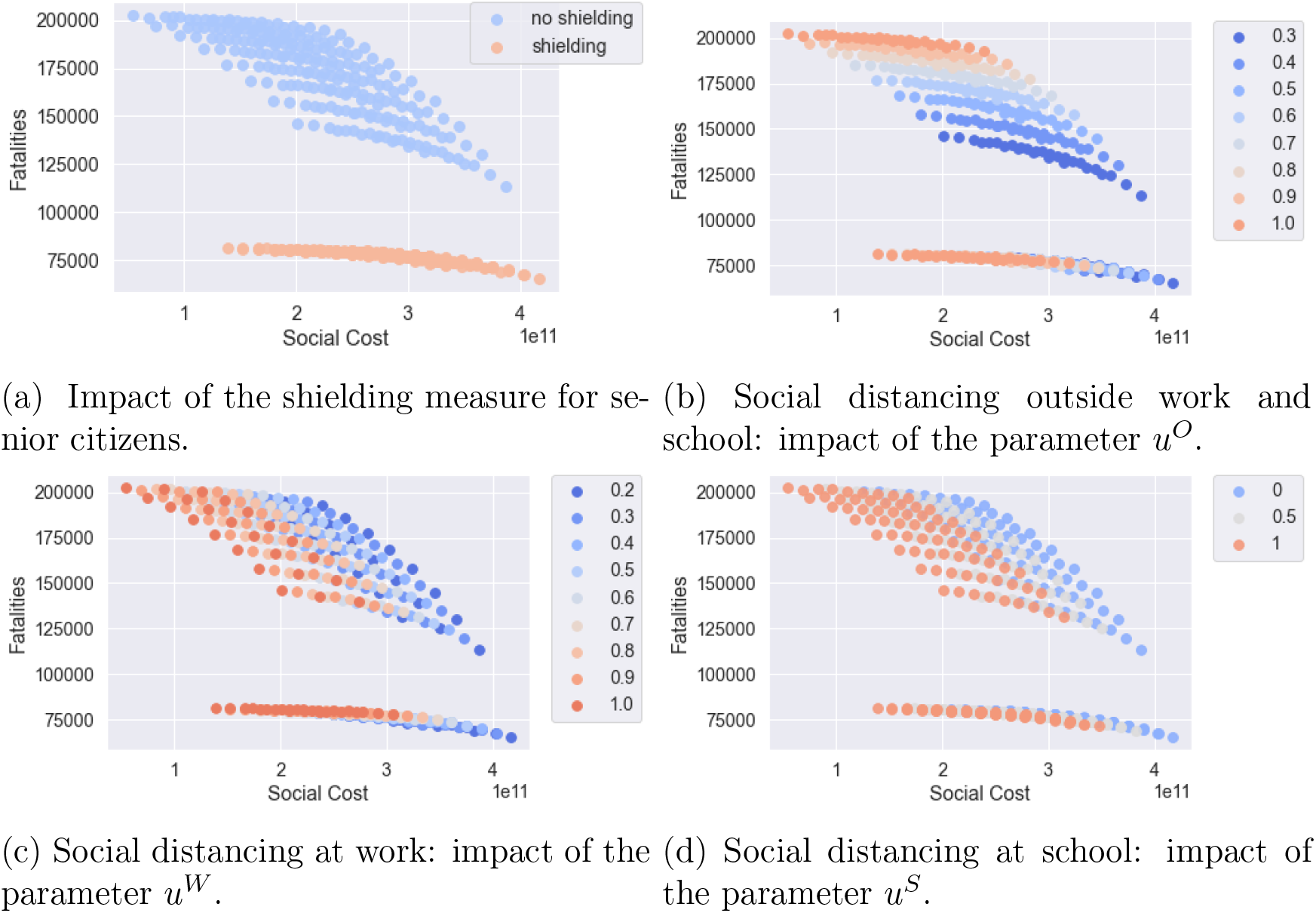
Efficiency plot of social cost against projected fatalities for the shielding measure

**Figure 31:**
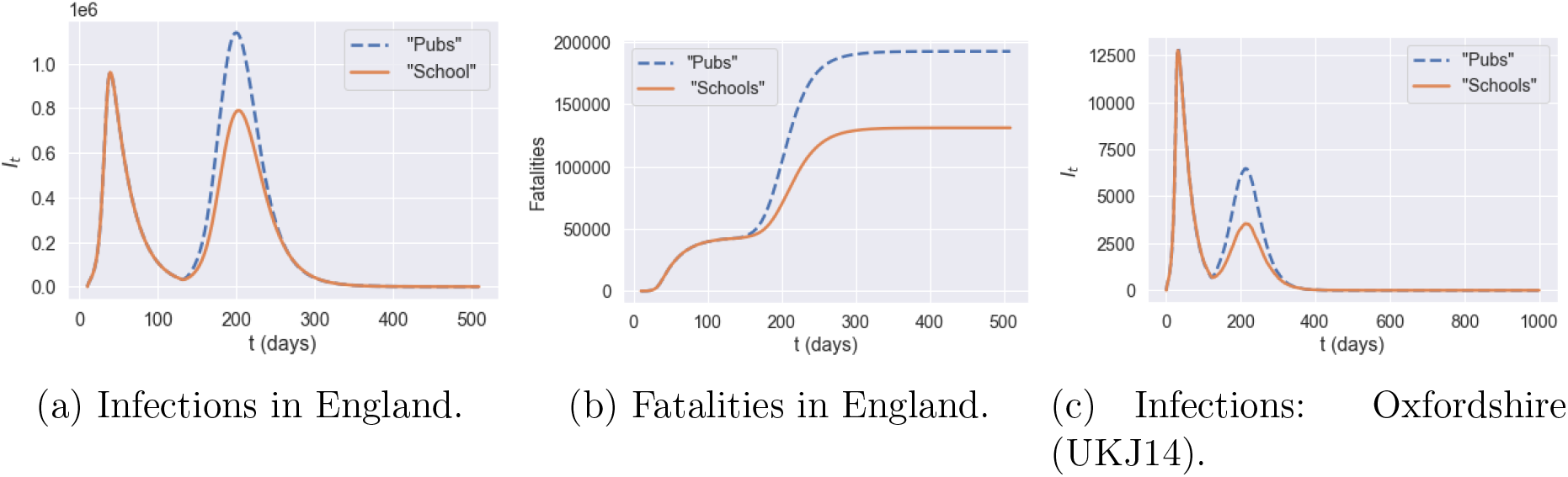
‘Open pubs’ versus ‘open schools’ policy.

#### 6.2.2 Shielding of senior citizens

We have examined the impact of shielding in isolation and also in combination with other measures such as school closure and social distancing.

As shown in Figure 30a, whether applied in isolation or in combination with other measures, shielding of elderly populations is by far the most effective measure for reducing the number of fatalities. As clearly shown in Figure 30a, regardless of the trade-off between social cost and health outcome, a policy which neglects shielding of the elderly is not efficient and its outcomes can always be improved through shielding measures.

#### Impact of shielding on policy efficiency

For policies without shielding, the level of social gatherings, *u^O^*, is the leading factor to determine the efficiency frontier. In Figures 33a, the efficiency frontier contains two classes of policies: ‘School and Work’ and ‘No Pubs’.

- ‘School and Work’ policies, which do not include any restrictions on school or work (*u^S^* = 1, *u^W^* = 1) but varying level of restrictions on social gatherings (0.3 ≤ *u^O^* ≤ 1). Within this class of policies, different level of social gatherings lead to very different outcome of fatalities, as illustrated in Figure 33a.
- ‘No Pubs’ policies, where social gatherings outside school and work are restricted (*u^O^* = 0.3), with different levels of social distancing *u^S^* ∈ {0, 0.5,1} *u^W^* ∈ [0.2,1] at school and work.

**Figure 32:**
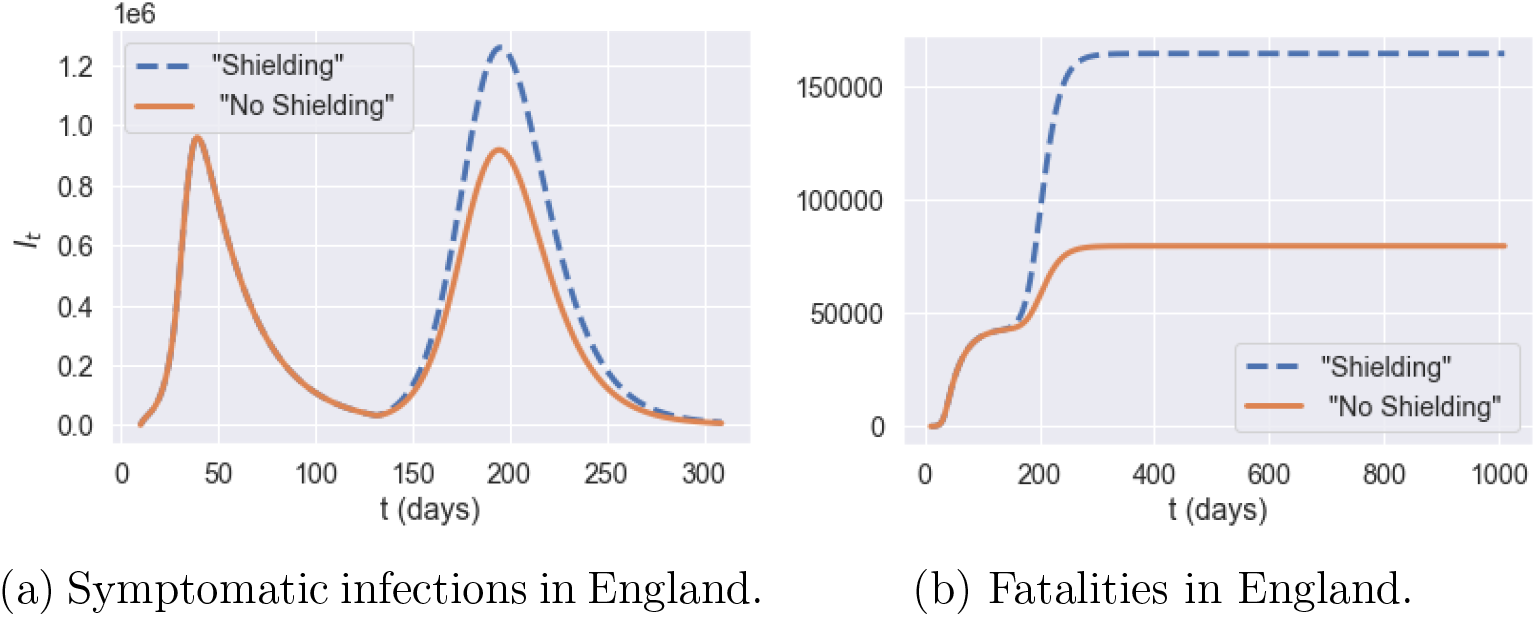
Comparison of policies with and without shielding in place, *u* = (1; 0:0; 1:0; 0:5). **Blue:** no shielding; **orange:** shielding in place.

**Figure 33:**
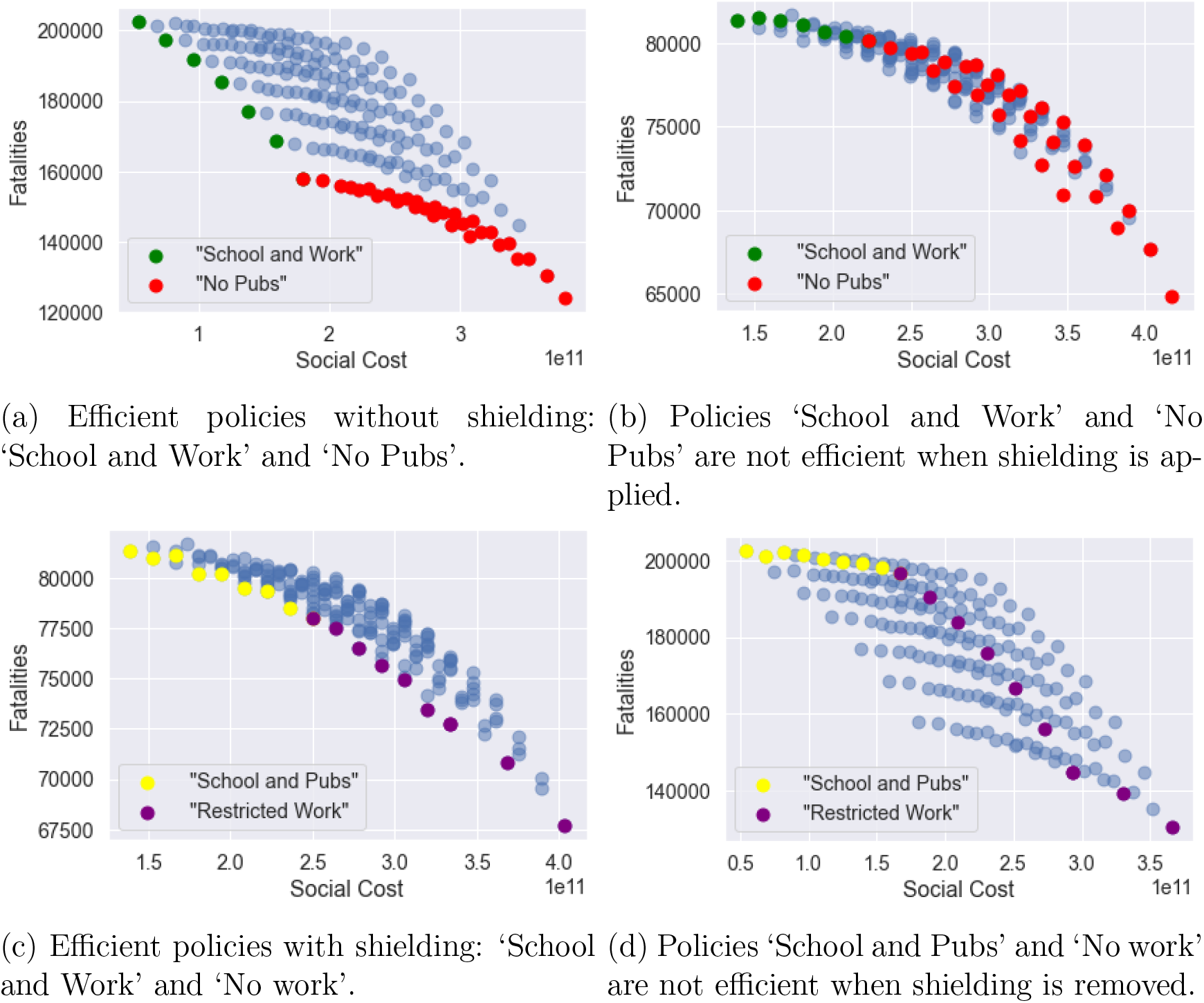
Impact of shielding on the efficiency frontier.

However, as observed in Figure 33b, these policies are not efficient when shielding measures are put in place for the elderly.

Under shielding, the spectrum of efficient policies is parameterised by the fraction *u^W^* of the workforce returning to work. As shown in Figure 33c, we can distinguish two classes of efficient policies under shielding:

- ‘School and Pubs’, consisting of policies without restrictions on schools or social gatherings (*u^S^* = 1, *u^O^* = 1) and different levels *u^W^* of restrictions on workplace gatherings.
- ‘Restricted Work’ policies, under which only ‘essential’ workers are allowed on-site work (*u^W^* = 0.2), with either

i. no school restrictions (*u^S^* = 1) and different levels of restrictions on social gatherings (0.2 ≤ *u^O^* ≤ 1) or
ii. restrictions on social gatherings (*u^O^* = 0.3, that is ‘no pubs’) and different levels of social distancing in school (0 ≤ *u^S^* ≤ 1).

As Figure 33d illustrates, ‘School and Pubs’ and ‘Restricted Work’ policies are not efficient without shielding.

In absence of shielding, social gatherings seem to be the main vector for contagion. When shielding measures are put in place, the social contacts associated with the elderly are reduced to the same level as under lockdown; in this case, contacts at work become the main vector of contagion.

## Adaptive mitigation policies

We now consider *adaptive* mitigation policies, in which the daily number of (national or regional) reported cases is used as a trigger for social distancing measures. Such policies have been recently implemented, in the U.K. and elsewhere, at a local or national level with various degrees of success. We distinguish *centralised* policies, based on monitoring of national case numbers, from *decentralised* policies where monitoring and implementation of measures are done at the level of (NUTS-3) regions.

### Centralised policies

We consider centralised policies which monitor the number of daily reported cases at country level. Whenever the whenever the number of daily reported cases (per 100,000) exceeds a threshold *R*_on_, confinement measures are imposed for a minimum of *L* days, until the number of daily reported cases falls below the threshold *R*_off_ < *R*_on_. Outside these lockdown periods we assume social distancing is in place with a compliance level *m* is the social compliance; we use a default value of *m* = 0.5.

This policy is implemented after the initial lockdown (i.e after July 4, 2020). In terms of the social contact matrix, we have, for *t* > *t*_0_ + *T*,

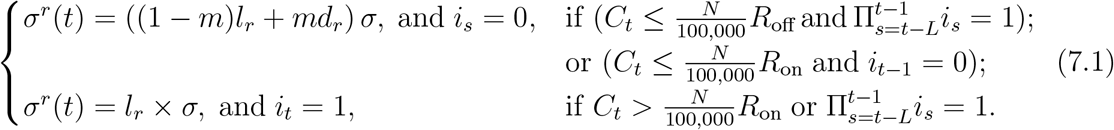

Here *T* = 105, *i_t_* is the indicator of whether lockdown is applied on day *t* and *C_t_* is the daily reported cases in England on day *t*. 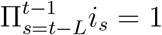 if lockdown has been applied for *L* consecutive days during the period [*t* − *L, t* − 1].

We simulate the dynamics with various choices of *R*_off_ and *R*_on_:

- *R*_on_ ∈ {2,4, 6, 8,10} (daily reported cases per 100,000 inhabitants); and
- *R*_off_ = 0.2 *R*_on_, *R*_off_ = 0.4 *R*_on_ or *R*_off_ = 0.8 *R*_on_.

We assume that once a lockdown is triggered it lasts a minimum of *L* = 7 days and that, once lockdown is removed, individuals continue to observe social distancing as measured by the parameter *m* ∈ [0,1]. Data on real-time mobility monitoring in the UK^11^, indicate mobility to be at 50% of normal level during the post-lockdown period, and thus we use *m* = 0.5 as a default value.

#### Example

Figure 35 shows an example of such an adaptive policy, where lockdown is triggered when daily cases exceeds 2240 nationally, and maintained until the count of new daily cases drops to 896. In the scenario shown in Fig. 35a, this results in four short lockdowns, totaling 34 days in all, which bring under control the national progression of the epidemic and avoid a ’second peak’ at national level. However, as shown in Figure 35b, this policy is less successful at regional level, resulting in a regional outbreak in Leicester.

**Figure 34:**
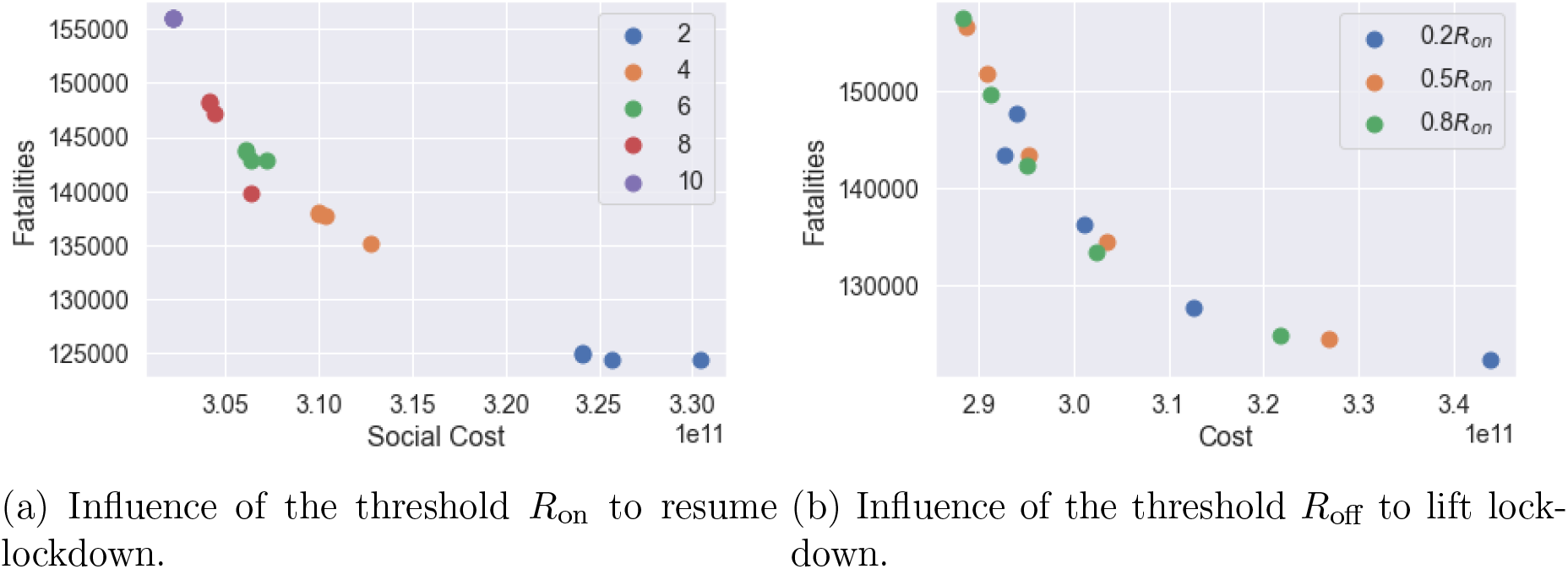
Social cost against fatalities when m = 0:5.

**Figure 35:**
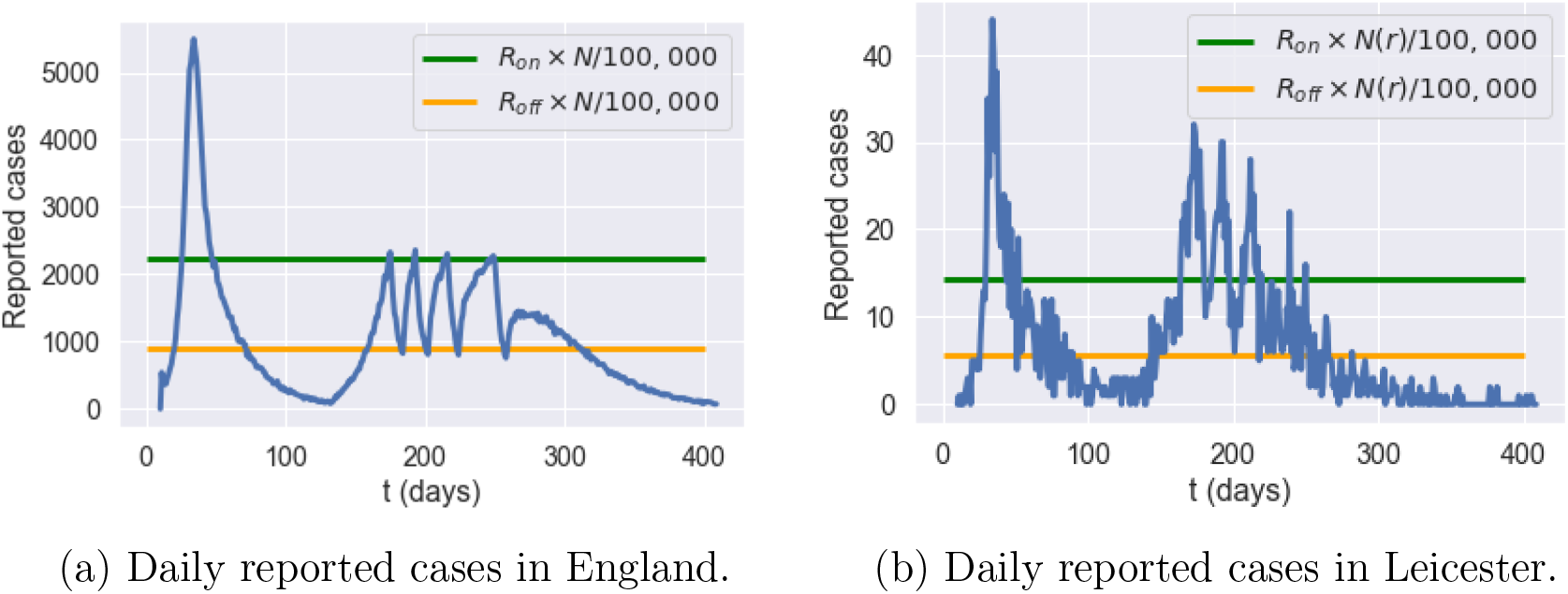
Simulation of reported cases in England and Leicester under a centralized triggering policy with *R*_on_ = 4, *R*_off_ = 0,4 × *R*_on_, *m* = 0:5 and no shielding

#### Impact of the triggering threshold

*R*_on_ The trigger threshold *R*_on_ has a significant impact on the efficiency of the policy. Smaller *R*_on_ values correspond to more frequent lockdowns, leading to a larger social cost and fewer fatalities. Here we compare the impact of the triggering threshold *R*_on_ when *m* = 0.5 and *R*_off_ = 0.4 × *R*_on_.

We observe in our simulations a second peak in *I_t_* for England when *R*_on_ = 10, while we observe no second peak when *R*_on_ = 2. When *R*_on_ = 2, *I_t_* remains at level 2 × 10^5^ with frequent interventions for 200 days and then decreases to zero. The social cost for policy *R*_on_ = 10 and policy *R*_on_ = 2 are 2.9 and 3.2, respectively. Policy *R*_on_ = 2 have 20% fewer fatalities compared to policy *R*_on_ = 10. Oxfordshire exhibits the same profile as England when *R*_on_ = 10. However, the shape of *I_t_* is different for *R*_on_ = 2 where Oxfordshire experiences a small outbreak around day 400.

In summary, smaller *R*_on_ values correspond to more frequent lockdowns and result in damping or elimination of the ’second peak’.

#### Increasing testing capacity

To study the effect of an increased testing capacity, we assume wide testing is adopted such that the reporting probability is increased from 4.5% to a significantly higher level (20%, 50%) on July 4th (see Figure 37).

**Figure 36:**
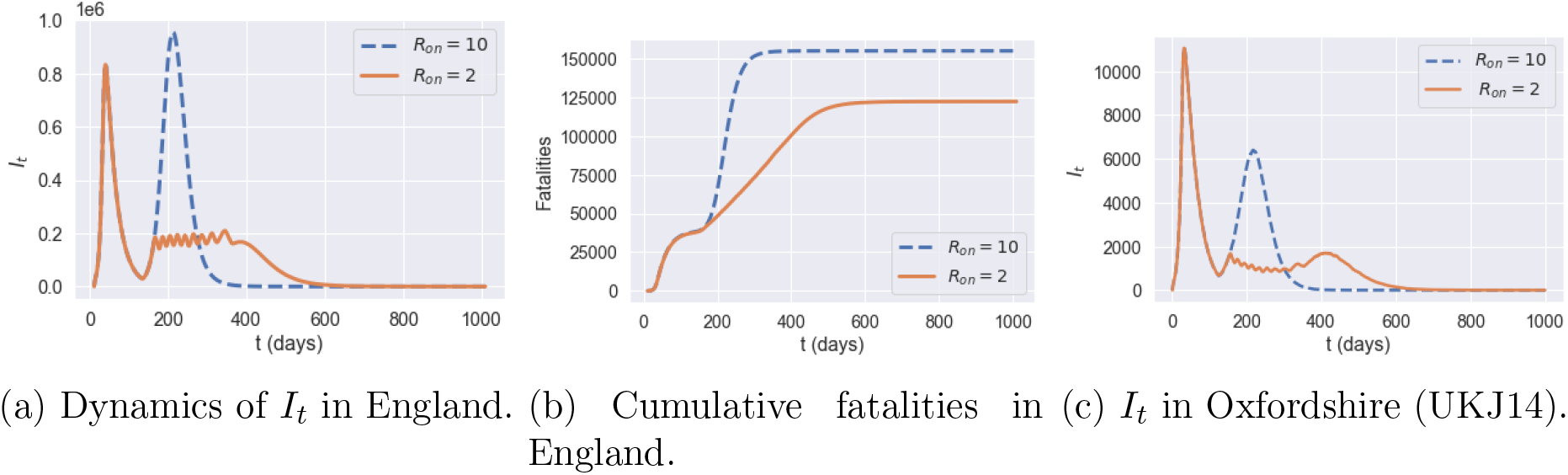
Comparison between triggering thresholds *R*_on_ = 10 and *R*_on_ = 2.

**Figure 37:**
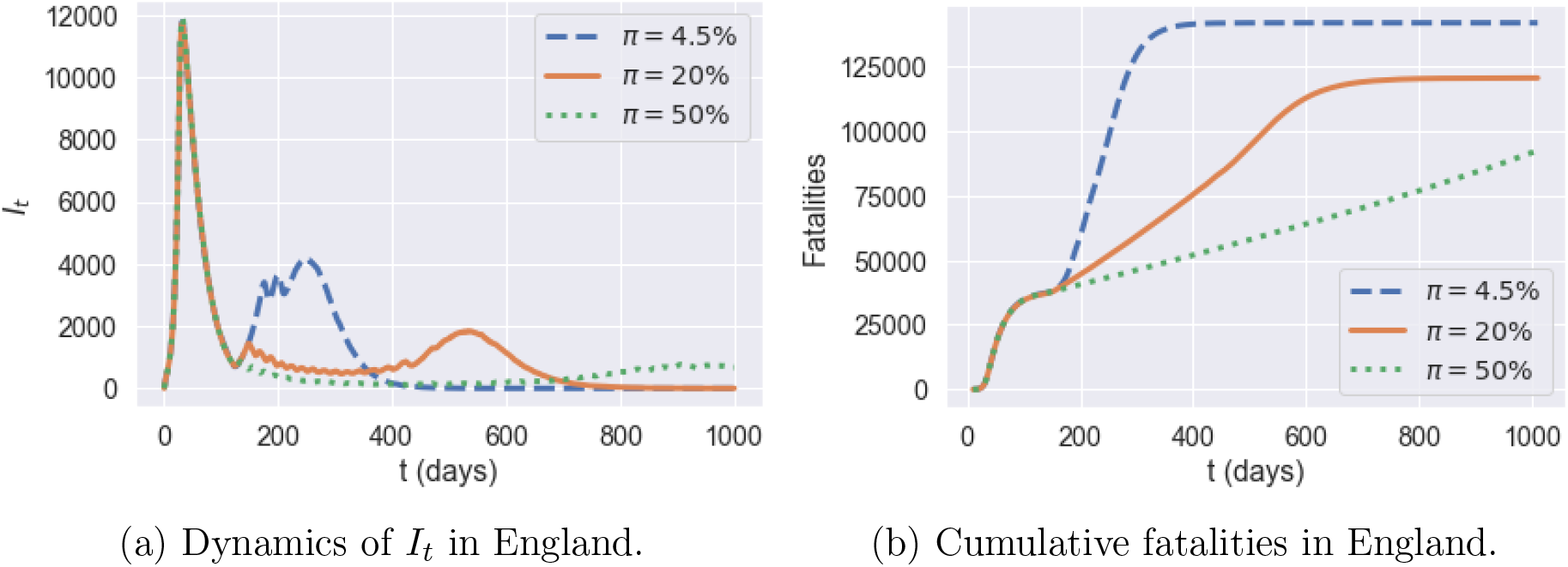
Reporting probabilities *π* = 4.5% (blue line) versus *π* = 20% (orange line) and *π* = 50% (green line). Policy: *R*_on_ = 6, *R*_off_ = 0.2*R*_on_, and *m* = 0.5.

By increasing the testing capacity, the observable quantity of daily reported cases becomes more consistent with the underlying dynamics of *I_t_*. Compared to the policy with a reporting probability *π* = 4.5% throughout the reference period, we see that the dynamics of *I_t_* when *π* = 50% decrease to a small value rapidly. Increasing the testing capacity also implies a more efficient control and as a result leads to fewer fatalities.

### Decentralised policies

We now consider a decentralised version of the above policies, based on monitoring of regional number of cases as triggers for regional confinement measures. In terms of the social contact matrices, we have, for *t* > *t*_0_ + *T*,

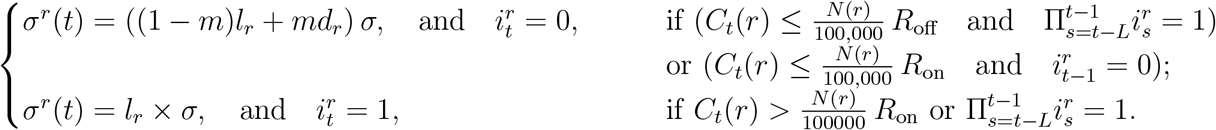

Here 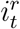 is the indicator of whether lockdown is applied in region *r* on day *t* and *C_t_*(*r*) is the daily number of cases reported in region *r* on day *t*. The term 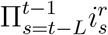 is used to track if lockdown has been applied in region *r* for *L* consecutive days during [*t* − *L, t* − 1]. We use the same values of *R*_on_ and *R*_off_ as in Section 7.1.

Figure 39 compares the outcomes of centralised and decentralised triggering policies. Decentralised policies are observed to always improve over centralised policies.

**Figure 38:**
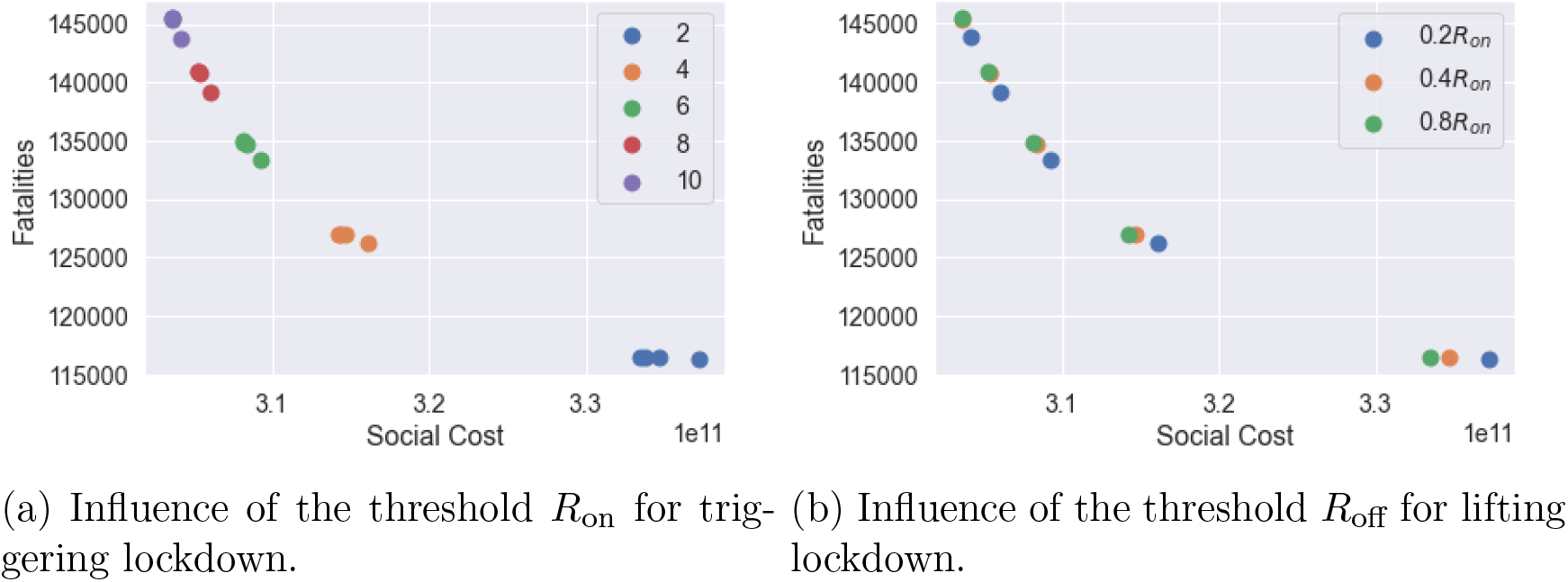
Decentralised confinement triggered by regional daily case numbers: social cost versus fatalities (*m* = 0.5).

**Figure 39:**
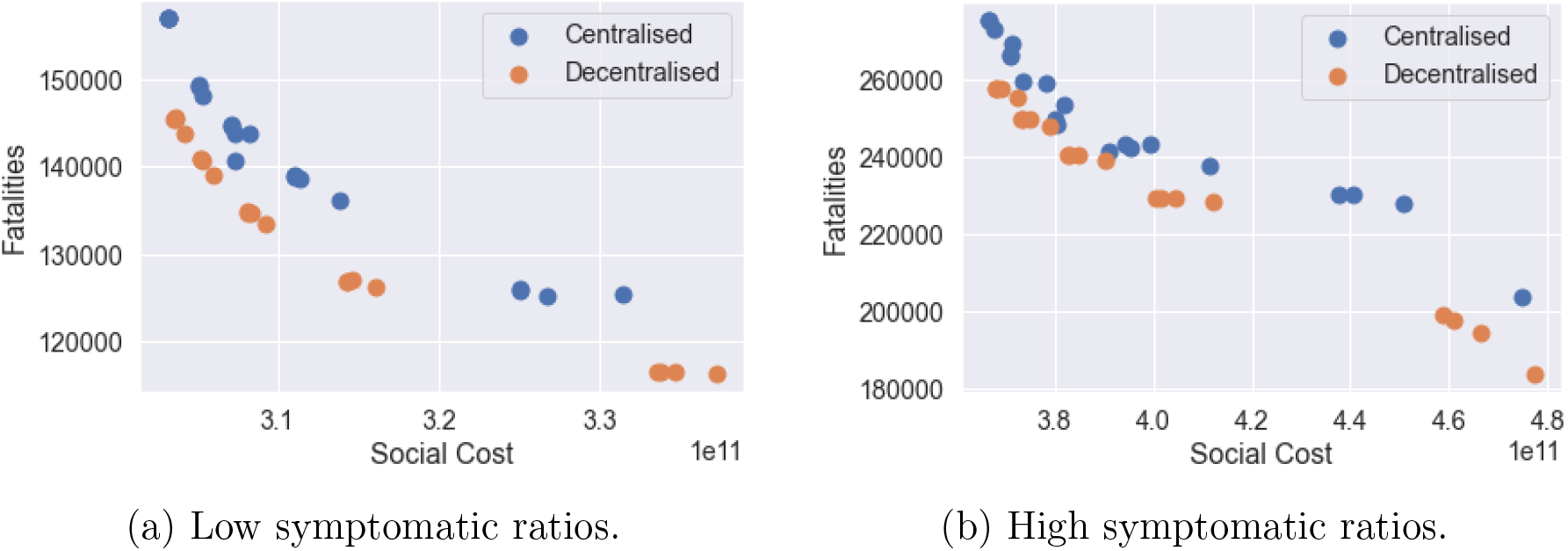
Efficiency analysis for centralised (**blue**) and decentralised (**orange**) adaptive mitigation policies. Outcomes are averaged across 100 simulated scenarios.

As an example, for *R*_on_ = 4 and *R*_off_ = 0.4*R*_on_ fatalities in England are 137, 700 under the centralised policy and 126, 300 under the decentralised policy, that is 8% lower.

Figure 41 compares regional fatalities per 100,000 habitants for these policies. For more than 90% of the regions, decentralised measures lead to fewer fatalities. The most effective reductions are in Dorset, South West England (UKK22) with 23% fewer fatalities and in Portsmouth (UKJ31) with 22% fewer fatalities. There are a few exceptions (see regions in light blue in Figure 41c). These regions are already under control before adaptive policies are applied. Therefore the improvement of moving from centralised policy to decentralised policy is limited.

**Figure 40:**
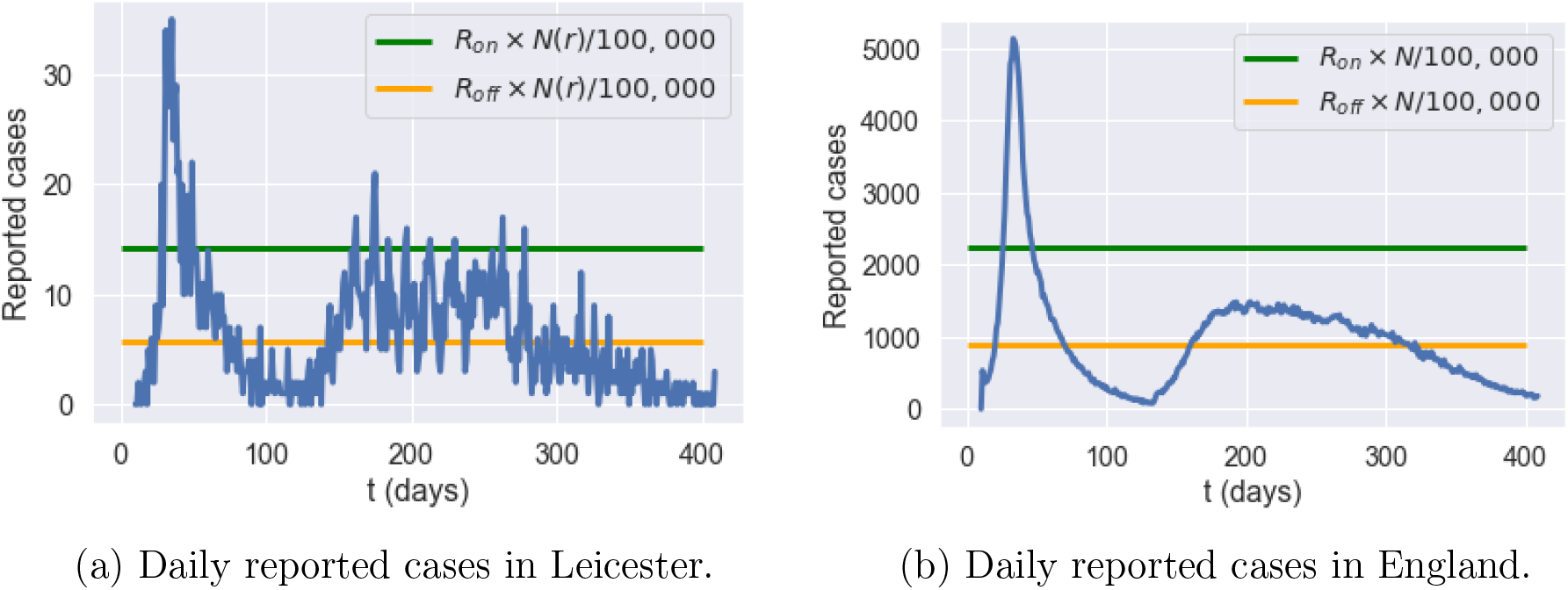
Reported cases in England and Leicester under a decentralised triggering policy: average of 50 simulated scenarios with *R*_on_ = 4, *R*_off_ = 0.4 × *R_on_, m* = 0.5, no shielding.

**Figure 41:**
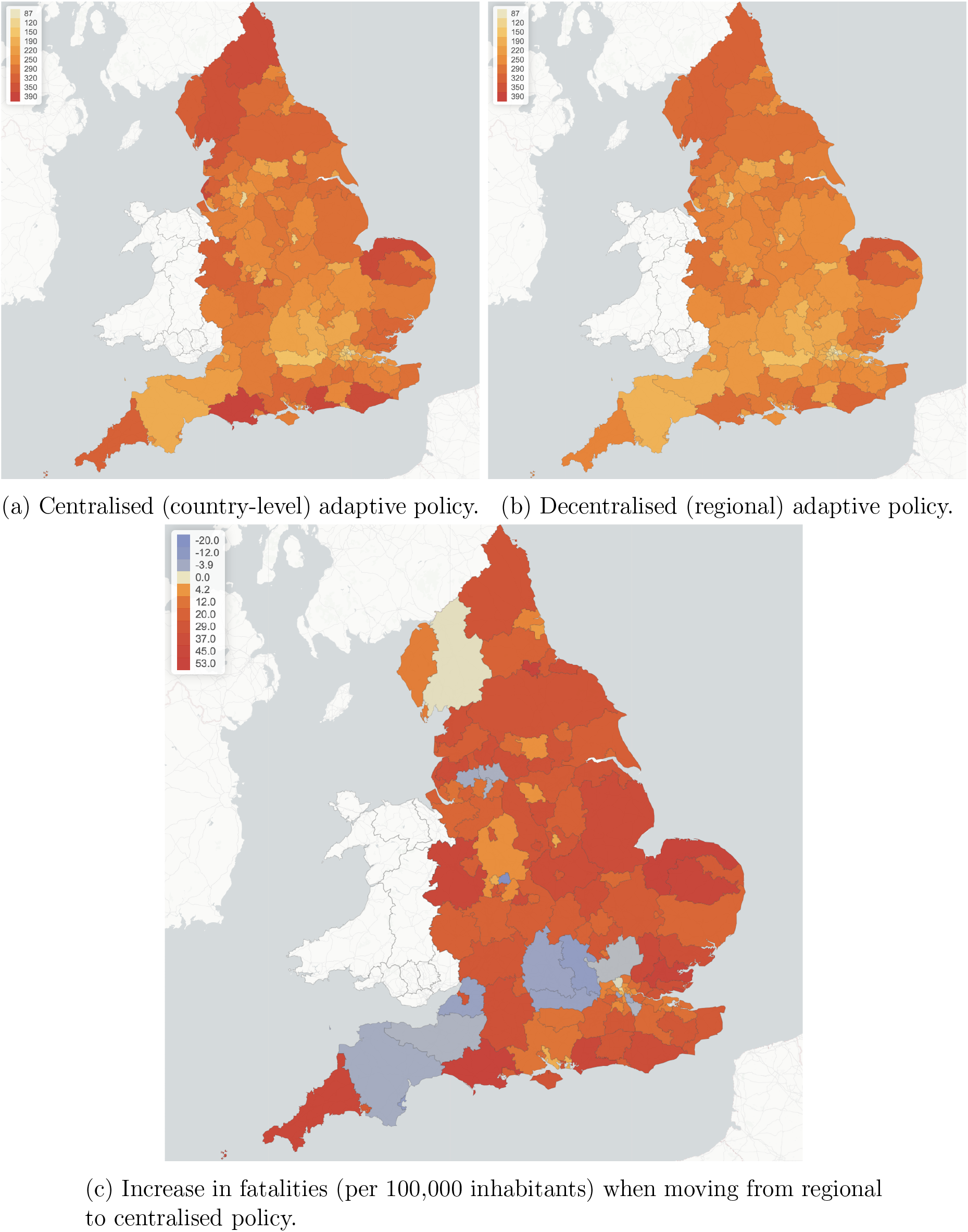
Fatalities per 100,000 inhabitants for centralised (left) verus regional (right) adaptive mitigation policies. Same triggering thresholds are used in both cases: *R*on = 4 and *R*off = 0:4*R*on.

Figure 42a compares the dynamics of symptomatic infections (*I_t_*) for the same example. There is a reduction of 100,000 in the amplitude of the second peak value when moving from the centralised policy to decentralised one. Decentralised policy also damps the second-peak values in most of the regions. Similar effects are observed for York (Figure 42c) and Leicester (Figure 42b).

**Figure 42:**
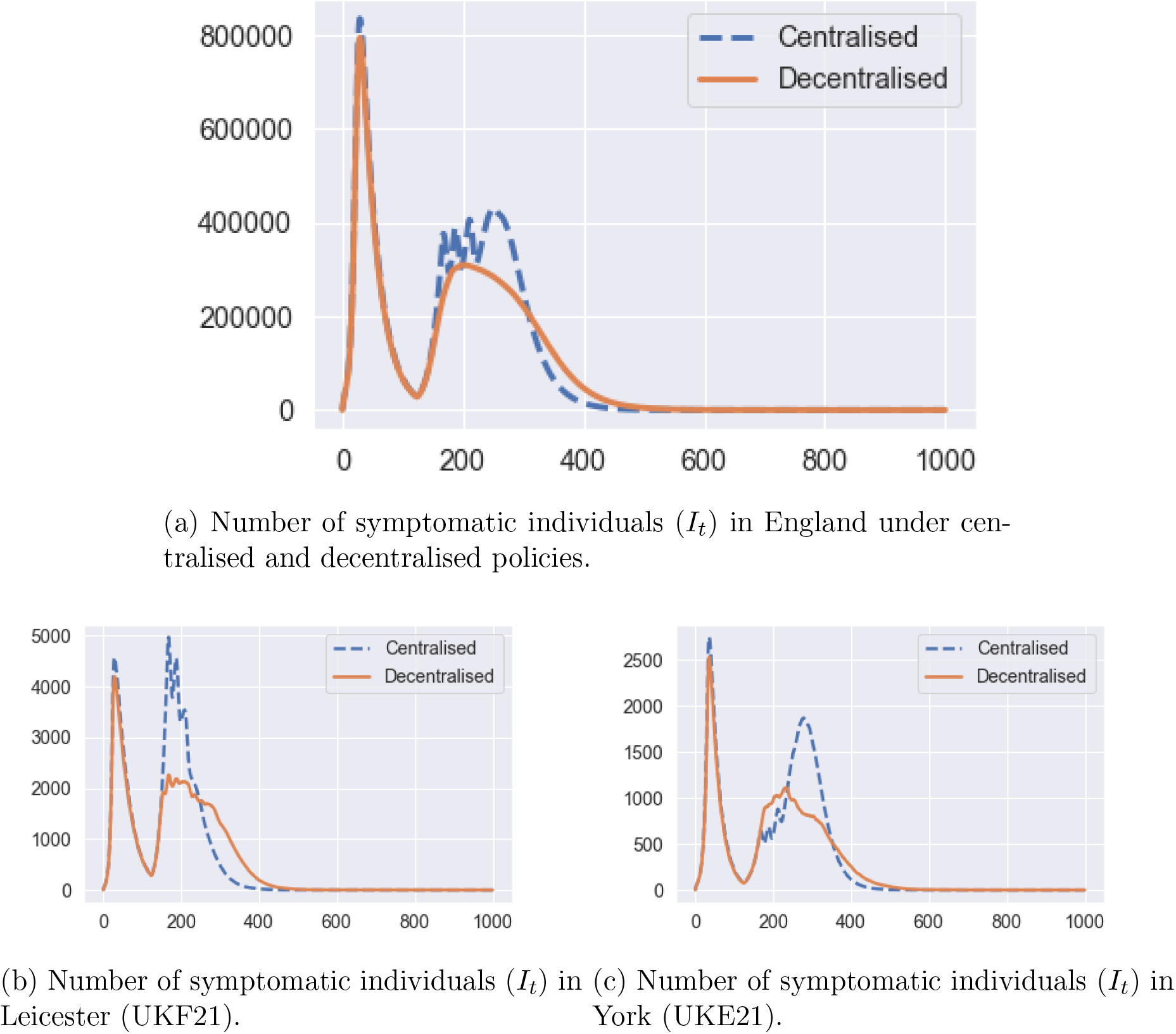
Number of infected individuals under under centralised (**blue dashed line**) and decentralised (**orange solid line**) policies. Same triggering thresholds are used in both cases: *R*_on_ = 4 and *R*_off_ = 0.4*R*_on_.

On June 29, 2020, Leicester became the first city in Britain to be placed in a local lockdown, after public health officials voiced concern at the city’s alarming rise in COVID-19 cases. Earlier in June, the Government announced that parts of the city would be released from lockdown, while a “targeted” approach will see pockets remain under tighter restrictions. Our simulations indicate a 60% reduction of the second-peak value in Leicester when a decentralised policy is implemented (Figure 42b).

#### Example

Figure 40 shows an example of such a decentralised triggering policy, with the same triggering thresholds as in the centralised example in Fig. 35. At regional level, we see in Figure 40a that this policy is more successful than the centralised policy in taming the local outbreaks in Leicester, substantially reducing the second peak through 8 one-week regional lockdowns. At the national level this results in a strong damping of ’second wave’ infections, as shown in Fig. 40b (compare with Fig. 35a).

### Adaptive versus pre-planned policies

Figure 43 compares the health outcome and social cost of the efficient policies considered in Sections 6.2, 7.1 and 7.2. The efficient frontier of pre-planned policies are among policies with *u^S^* ∈ {0, 0.5,1}, 0.2 ≤ *u^W^ ≤* 1.0 and 0.3 ≤ *u^O^* ≤ 1.0. For centralised and decentralised policies, *m* = 0.25, 0.5, 0.75, 1; *R*_on_ = 2, 4, 6, 8, 10; and *R*_off_ = *p* × *R*_on_ with *p* = 0.2, 0.4, 0.8.

**Figure 43:**
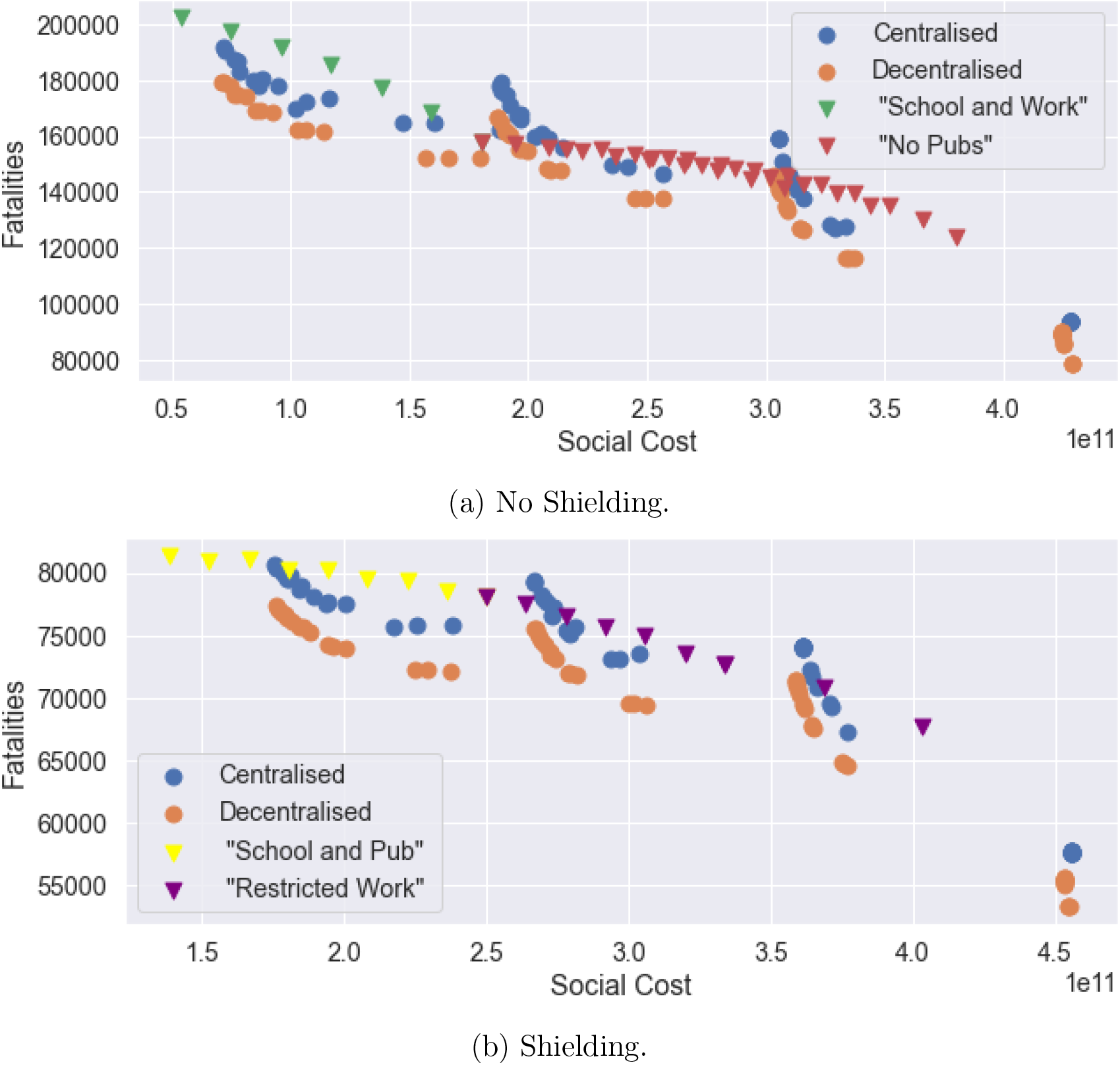
Efficiency plot: pre-planned versus adaptive mitigation policies.

We observe that

- Adaptive policies, in which measures are triggered when the number of daily new cases exceeds a threshold, are more efficient than pre-planned policies; and
- As shown in Figures 43a and 43b, a decentralised policy is more efficient than both centralised policy and pre-planned policy.

In Table 8, we provide a summary of outcomes for five different types of policies.

- Confinement followed by social distancing: *T* = 105 and *m* = 0.5 and no shielding.
- Pre-planned policy: social distancing at work and school (*u^H^* = 1, *u^S^* = 0.5,*u^W^* = 0.5), restrictions on social gatherings (*u^O^* = 0.3) and no shielding.
- Centralised and decentralised triggering policies (Sec. 7.1 and Sec. 7.2) with *m* = 0.5, *R*_on_ = 4, *R*_off_ = 0.4*R*_on_ and no shielding;
- Decentralised triggering combined with shielding of elderly populations: *m* = 0.5, *R*_on_ = 4, *R*_off_ = 0.4*R*_on_;
- ’Protect Lives’ policy: in the range of efficient policies, the one which results in the fewest fatalities is a decentralised triggering policy with *R*_on_ = 2, *R*_off_ = 0.2*R*_on_ (so more frequent triggering of confinement measures than the above), high degree of social distancing (*m* = 0.25) and shielding of elderly populations. This policy corresponds to the point in the lower right corner of Figure 43b. The social cost is 4.52, much higher than the other policies considered.

**Table 7:** Average social cost and fatalities for a given policy with different testing capacities (50 paths). Policy: *R*_on_ = 6, *R*_off_ = 0.4*R*_on_, and *m* = 0.5.

| Reporting prob. $\pi$ | 4.5% | 20% | 50% |
| --- | --- | --- | --- |
| Social Cost ($10^{11}$) | 3.0 | 3.4 | 4.0 |
| Fatalities | 141,700 | 120,400 | 92,300 |

**Table 8:** Summary of outcomes for different policies, starting from the same initial conditions on July 4, 2020.

| Policy | $I_t$<br>(Aug 1) | $A_t$<br>(Aug 1) | Fatalities<br>(Aug 1) | max $I_t$<br>(2nd peak) | Social<br>cost | Total<br>fatalities |
| --- | --- | --- | --- | --- | --- | --- |
| Confinement followed by social distancing | 124,400 | 462,000 | 42,000 | 960,700 | 3.0 | 165,700 |
| Pre-planned | 112,000 | 436,000 | 43,800 | 819,300 | 3.1 | 138,700 |
| Centralised triggering | 102,000 | 378,500 | 39,400 | 430,800 | 3.11 | 137,700 |
| Decentralised triggering | 97,200 | 379,800 | 38,500 | 309,400 | 3.14 | 126,300 |
| Decentralised triggering + shielding | 70,700 | 309,000 | 38,291 | 295,500 | 3.65 | 67,900 |
| 'Protect Lives' | 32,000 | 132,500 | 37,400 | 81,700 | 4.50 | 53,000 |

Outcomes are averaged across 50 scenarios, starting from the same initial conditions on July 4 (end of the national UK lockdown).

#### Regional outcomes

Comparing the regional outcomes of the centralised, decentralised and pre-planned policies displayed in Table 8 shows that the decentralised triggering policies are able in many cases to considerably damp the ‘second wave’ of infections. Figure 44 illustrates this in the case of Leicester and Birmingham: the decentralised triggering policy reduces the second peak amplitude by more than two-thirds compared to the pre-planned policy.

**Figure 44:**
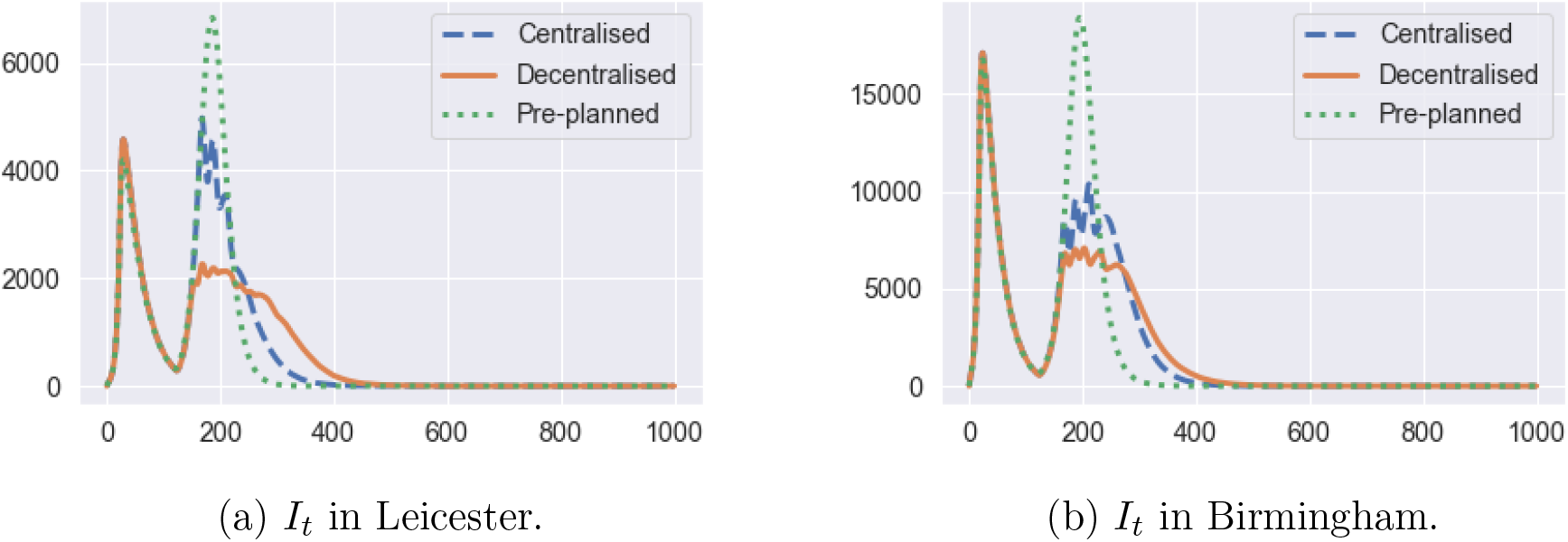
Regional comparison of pre-planned and adaptive mitigation policies.

## Data Availability

All data used in the article is publicly available and the data sources are indicated in the article.

## Demographic regions

Table 9 details the used mapping between Upper Tier Local Authority (UTLA) region codes and the Nomenclature of Territorial Units for Statistics at level 3 (NUTS-3) codes. If a single UTLA region falls within multiple NUTS-3 regions, the data is distributed proportionally to the population size in each of these regions. On the other hand if multiple UTLA regions are contained within a single NUTS-3 region, the data is simply aggregated.

**Table 9:** Mapping between the Upper Tier Local Authority (UTLA) regions and the Nomenclature of Territorial Units for Statistics at level 3 codes (NUTS-3).

| UTLA Code | UTLA Region Name | NUTS-3 Code Mapping |
| --- | --- | --- |
| E06000001 | Hartlepool | UKC11 |
| E06000002 | Middlesbrough | UKC12 |
| E06000003 | Redcar and Cleveland | UKC12 |
| E06000004 | Stockton-on-Tees | UKC11 |
| E06000005 | Darlington | UKC13 |
| E06000006 | Halton | UKD71 |
| E06000007 | Warrington | UKD61 |
| E06000008 | Blackburn with Darwen | UKD41 |
| E06000009 | Blackpool | UKD42 |
| E06000010 | Kingston upon Hull, City of | UKE11 |
| E06000011 | East Riding of Yorkshire | UKE12 |
| E06000012 | North East Lincolnshire | UKE13 |
| E06000013 | North Lincolnshire | UKE13 |
| E06000014 | York | UKE21 |
| E06000015 | Derby | UKF11 |
| E06000016 | Leicester | UKF21 |
| E06000017 | Rutland | UKF22 |
| E06000018 | Nottingham | UKF14 |
| E06000019 | Herefordshire, County of | UKG11 |
| E06000020 | Telford and Wrekin | UKG21 |
| E06000021 | Stoke-on-Trent | UKG23 |
| E06000022 | Bath and North East Somerset | UKK12 |
| E06000023 | Bristol, City of | UKK11 |
| E06000024 | North Somerset | UKK12 |
| E06000025 | South Gloucestershire | UKK12 |
| E06000026 | Plymouth | UKK41 |
| E06000027 | Torbay | UKK42 |
| E06000030 | Swindon | UKK14 |
| E06000031 | Peterborough | UKH11 |
| E06000032 | Luton | UKH21 |
| E06000033 | Southend-on-Sea | UKH31 |
| E06000034 | Thurrock | UKH32 |
| E06000035 | Medway | UKJ41 |
| E06000036 | Bracknell Forest | UKJ11 |
| E06000037 | West Berkshire | UKJ11 |
| E06000038 | Reading | UKJ11 |
| E06000039 | Slough | UKJ11 |
| E06000040 | Windsor and Maidenhead | UKJ11 |
| E06000041 | Wokingham | UKJ11 |
| E06000042 | Milton Keynes | UKJ12 |
| E06000043 | Brighton and Hove | UKJ21 |
| E06000044 | Portsmouth | UKJ31 |
| E06000045 | Southampton | UKJ32 |
| E06000046 | Isle of Wight | UKJ34 |
| E06000047 | County Durham | UKC14 |
| E06000049 | Cheshire East | UKD62 |
| E06000050 | Cheshire West and Chester | UKD63 |
| E06000051 | Shropshire | UKG22 |
| E06000052 | Cornwall and Isles of Scilly | UKK30 |
| E06000054 | Wiltshire | UKK15 |
| E06000055 | Bedford | UKH24 |
| E06000056 | Central Bedfordshire | UKH25 |
| E06000057 | Northumberland | UKC21 |
| E06000058 | Bournemouth and Poole | UKK21 |
| E06000059 | Dorset | UKK22 |
| E08000001 | Bolton | UKD36 |
| E08000002 | Bury | UKD37 |
| E08000003 | Manchester | UKD33 |
| E08000004 | Oldham | UKD37 |
| E08000005 | Rochdale | UKD37 |
| E08000006 | Salford | UKD34 |
| E08000007 | Stockport | UKD35 |
| E08000008 | Tameside | UKD35 |
| E08000009 | Trafford | UKD34 |
| E08000010 | Wigan | UKD36 |
| E08000011 | Knowsley | UKD71 |
| E08000012 | Liverpool | UKD72 |
| E08000013 | St. Helens | UKD71 |
| E08000014 | Sefton | UKD73 |
| E08000015 | Wirral | UKD74 |
| E08000016 | Barnsley | UKE31 |
| E08000017 | Doncaster | UKE31 |
| E08000018 | Rotherham | UKE31 |
| E08000019 | Sheffield | UKE32 |
| E08000021 | Newcastle upon Tyne | UKC22 |
| E08000022 | North Tyneside | UKC22 |
| E08000023 | South Tyneside | UKC22 |
| E08000024 | Sunderland | UKC23 |
| E08000025 | Birmingham | UKG31 |
| E08000026 | Coventry | UKG33 |
| E08000027 | Dudley | UKG36 |
| E08000028 | Sandwell | UKG37 |
| E08000029 | Solihull | UKG32 |
| E08000030 | Walsall | UKG38 |
| E08000031 | Wolverhampton | UKG39 |
| E08000032 | Bradford | UKE41 |
| E08000033 | Calderdale | UKE44 |
| E08000034 | Kirklees | UKE44 |
| E08000035 | Leeds | UKE42 |
| E08000036 | Wakefield | UKE45 |
| E08000037 | Gateshead | UKC22 |
| E09000001 | City of London | UKI31 |
| E09000002 | Barking and Dagenham | UKI52 |
| E09000003 | Barnet | UKI71 |
| E09000004 | Bexley | UKI51 |
| E09000005 | Brent | UKI72 |
| E09000006 | Bromley | UKI61 |
| E09000007 | Camden | UKI31 |
| E09000008 | Croydon | UKI62 |
| E09000009 | Ealing | UKI73 |
| E09000010 | Enfield | UKI54 |
| E09000011 | Greenwich | UKI51 |
| E09000012 | Hackney | UKI41 |
| E09000013 | Hammersmith and Fulham | UKI33 |
| E09000014 | Haringey | UKI43 |
| E09000015 | Harrow | UKI74 |
| E09000016 | Havering | UKI52 |
| E09000017 | Hillingdon | UKI74 |
| E09000018 | Hounslow | UKI75 |
| E09000019 | Islington | UKI43 |
| E09000020 | Kensington and Chelsea | UKI33 |
| E09000021 | Kingston upon Thames | UKI63 |
| E09000022 | Lambeth | UKI45 |
| E09000023 | Lewisham | UKI44 |
| E09000024 | Merton | UKI63 |
| E09000025 | Newham | UKI41 |
| E09000026 | Redbridge | UKI53 |
| E09000027 | Richmond upon Thames | UKI75 |
| E09000028 | Southwark | UKI44 |
| E09000029 | Sutton | UKI63 |
| E09000030 | Tower Hamlets | UKI42 |
| E09000031 | Waltham Forest | UKI53 |
| E09000032 | Wandsworth | UKI34 |
| E09000033 | Westminster | UKI32 |
| E10000002 | Buckinghamshire | UKJ13 |
| E10000003 | Cambridgeshire | UKH12 |
| E10000006 | Cumbria | UKD11, UKD12 |
| E10000007 | Derbyshire | UKF13, UKF12 |
| E10000008 | Devon | UKK43 |
| E10000011 | East Sussex | UKJ22 |
| E10000012 | Essex | UKH37, UKH34, UKH35, UKH36 |
| E10000013 | Gloucestershire | UKK13, UKK12 |
| E10000014 | Hampshire | UKJ36, UKJ37, UKJ35 |
| E10000015 | Hertfordshire | UKH23 |
| E10000016 | Kent | UKJ43, UKJ44, UKJ45, UKJ46 |
| E10000017 | Lancashire | UKD45, UKD46, UKD47, UKD44 |
| E10000018 | Leicestershire | UKF22 |
| E10000019 | Lincolnshire | UKE13, UKF30 |
| E10000020 | Norfolk | UKH15, UKH17, UKH16 |
| E10000021 | Northamptonshire | UKF24, UKF25 |
| E10000023 | North Yorkshire | UKE22 |
| E10000024 | Nottinghamshire | UKF15, UKF16 |
| E10000025 | Oxfordshire | UKJ14 |
| E10000027 | Somerset | UKK12, UKK23 |
| E10000028 | Staffordshire | UKG24 |
| E10000029 | Suffolk | UKH14 |
| E10000030 | Surrey | UKJ25, UKJ26 |
| E10000031 | Warwickshire | UKG13 |
| E10000032 | West Sussex | UKJ28, UKJ27 |
| E10000034 | Worcestershire | UKG12 |

## Baseline parameters for social contact rates

This appendix provides a description of sources used for baseline social contact rate parameters. Two sources have been used: the POLYMOD study [39], processed using PyRoss [52], and the BBC Pandemic study [28]. These parameters are used as a baseline and a further detailed calibration is carried out region by region to account for heterogeneity of social contact patterns across UK regions.

The PyRoss library [2] uses a Bayesian Hierarchical framework to decompose contact rates into ‘work’, ‘home’, ‘school’, and ‘other’ [43]. Using this approach we estimate a contact matrix for four age groups [0, 20), [20, 60), [60, 70) and [70,100) using UK demographic data.^12^ This leads to the following baseline contact matrices, visualized in Figure 45:

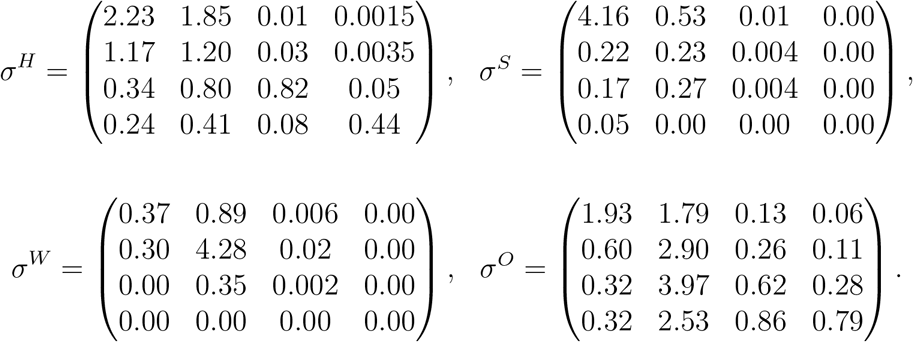

**Figure 45:**
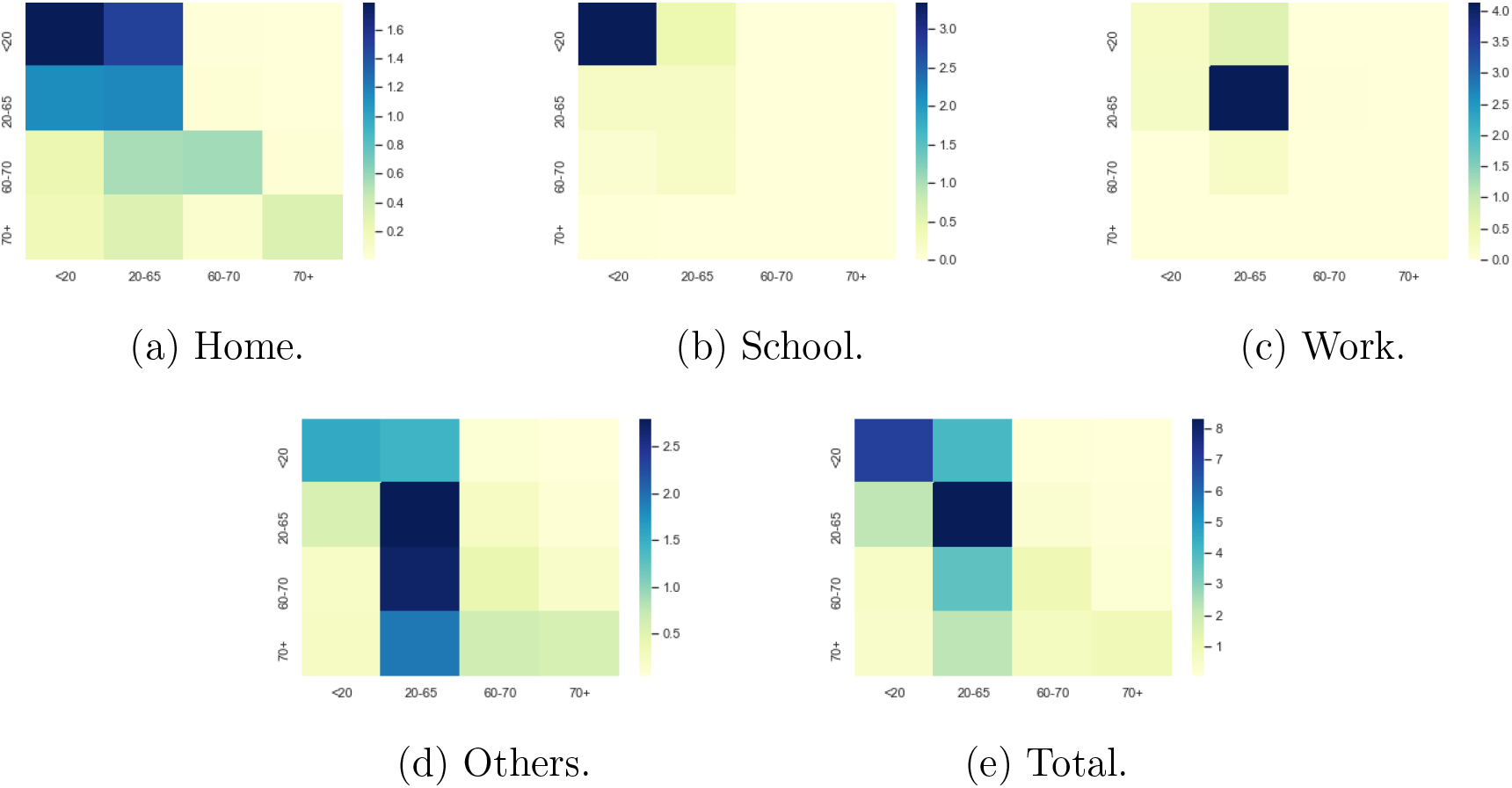
Baseline social contact matrices.

These baseline values are modulated to reflect regional differences, using the approach described in Section 3.

## Footnotes

See https://coronavirus.data.gov.uk/. Data retrieved on June 29, 2020.

See https://ec.europa.eu/eurostat/web/nuts/background.

See https://ec.europa.eu/eurostat/product?code=demo_r_pjangrp3&mode=view

See https://geoportal.statistics.gov.uk/datasets/c893dfece45f465f857ac34641041863_0 for a lookup table used in the process of mapping. If more than one UTLA region falls within the boundary of a single NUTS-3 region, the data is then aggregated. On the other hand, if a single UTLA region lies within more than one NUTS-3 region, the data is distributed among NUTS-3 regions in proportion to the total number of people living in each region.

https://www.ons.gov.uk/peoplepopulationandcommunity/populationandmigration/populationprojections/datasets/tablea21principalprojectionukpopulationinagegroup

See https://www.nomisweb.co.uk/census/2011/wu01ew. Data retrieved on January 2020.

See https://geoportal.statistics.gov.uk/datasets/c893dfece45f465f857ac34641041863_0.

https://www.legislation.gov.uk/ukpga/2020/7/contents/enacted

https://www.oxford-covid-19.com/

https://ourworldindata.org/coronavirus-testing

https://www.oxford-covid-19.com/

https://www.statista.com/statistics/281174/uk-population-by-age/

